# Global distribution of *Leptospira* serovar isolations and detections from animal host species: a systematic review and online database

**DOI:** 10.1101/2023.10.03.23296503

**Authors:** Nienke N. Hagedoorn, Michael J. Maze, Manuela Carugati, Shama Cash-Goldwasser, Kathryn J. Allan, Kevin Chen, Brieuc Cossic, Elena Demeter, Sarah Gallagher, Richard German, Renee L. Galloway, Josipa Habuš, Matthew P. Rubach, Kanae Shiokawa, Nadezhda Sulikhan, John A. Crump

**Author notes:** **Corresponding author:** Michael J. Maze, Address: Department of Medicine, University of Otago Christchurch 2 Riccarton Avenue, Christchurch 8011 New Zealand. Contributed equally (Nienke N. Hagedoorn, Michael J. Maze).

## Abstract

**Objectives:** *Leptospira,* the spirochaete causing leptospirosis, can be classified into >250 antigenically distinct serovars. Although knowledge of the animal host species and geographic distribution of *Leptospira* serovars is critical to understand the human and animal epidemiology of leptospirosis, currently data are fragmented. We aimed to systematically review the literature on animal host species and geographic distribution of *Leptospira* serovars to examine associations between serovars with animal host species and regions, and to identify geographic regions in need of study.

**Methods:** Nine library databases were searched from inception through 9 March 2023 using keywords including *Leptospira*, animal, and a list of serovars. We sought reports of detection of *Leptospira*, from any animal, characterized by cross agglutinin absorption test, monoclonal antibody typing, serum factor analysis, or pulsed-field gel electrophoresis to identify the serovar.

**Results:** We included 409 reports, published from 1927 through 2022, yielding data on 154 *Leptospira* serovars. The reports included data from 66 (26.5%) of 249 countries. Detections were from 144 animal host species including 135 (93.8%) from the class Mammalia, 5 (3.5%) from Amphibia, 3 (2.1%) from Reptilia, and 1 (0.7%) from Arachnida. Across the animal host species, *Leptospira* serovars that were detected in the largest number of animal species included Grippotyphosa (n=39), Icterohaemorrhagiae (n=29), Pomona (n=28), Australis (n=25), and Ballum (n=25). Of serovars, 76 were detected in a single animal host species. We created an online database to identify animal host species for each serovar by country.

**Conclusions:** We found that many countries have few or no *Leptospira* serovars detected from animal host species and that many serovars were detected from a single animal species. Our study highlights the importance of efforts to identify animal host species of leptospirosis, especially in places with a high incidence of human leptospirosis. We provide an updated resource for leptospirosis researchers.

## Introduction

Leptospirosis is a widespread zoonotic disease caused by bacteria from the genus *Leptospira,* which can infect and cause disease in both animals and humans.(1) *Leptospira* can be antigenically subdivided into >250 serovars that can be grouped into 30 serogroups.(2, 3) The antigenic classification into serovars is distinct from genomic classification systems of species or sequence type with imperfect overlap.(4)

Although *Leptospira* detections have been described in a large range of animal host species including mammals, reptiles, and amphibians,(5, 6) *Leptospira* spp. serovars (hereafter shortened as serovars) are thought to vary in their degree of adaptation to animal species. In some animal hosts, certain serovars may cause disease and establish chronic infection in which *Leptospira* persist in the renal and reproductive tracts and produce prolonged genito-urinary shedding. Some host-adapted serovars have their reservoir in specific animal host species, for instance serovar Icterohaemorrhagiae in *Rattus* species, and serovar Hardjo in *Bos taurus* (cattle).(6, 7)

Human *Leptospira* infection occurs through either direct contact with an infected animal or contact with soil or water contaminated by the urine of an infected animal.(8) *Leptospira* infection is responsible for >1 million human illnesses and 59,000 human deaths annually.(9) High incidence of leptospirosis is observed in tropical regions, urban slum environments, or following flooding. Because *Leptospira* are rarely isolated in culture and uncommonly detected using molecular methods from human samples, serology retains a critical role in the diagnosis of human leptospirosis. Serology can identify infecting *Leptospira* to the serogroup level which could aid to identify possible animal species as source of the human infection. Knowledge of local serogroups also informs the serovar composition of microscopic agglutination testing (MAT) antigen panels for locations.(10)

Knowledge of infecting serovar may have clinical implications as, although unconfirmed, there is some evidence that different serovars cause varied disease severity.(11) Understanding the animal host range and geographic distribution of serovars is essential during epidemiologic investigations to identify local animal hosts of serovars infecting people. Such information can subsequently inform control, potentially through diverse applications such as livestock vaccine composition, or rodent control programmes.(12) Conversely, gaps in our understanding of geographic and animal host distribution can impair control efforts when important host species are unrecognized.

To the best of our knowledge, the ‘Leptospiral serotype distribution lists’ in 1966,(13) and the update in 1975 (14) were the last publications that systematically documented the animal host range and geographic distribution of serovars. There has since been considerable research on leptospirosis, with developments in classification, and recognition of new serovars.(4) Further, the development of systematic review methods may identify additional reports prior to 1975 that were not included in the historic distribution lists. Although recent systematic reviews have focused on specific regions or animals,(15–17) a comprehensive and updated resource documenting the animal host range of serovars is lacking. Therefore, we aimed to synthesize the literature on animal host species and geographic distribution of *Leptospira* serovars to (i) examine associations between serovars with animal host species and regions, (ii) identify geographic regions in need of study, and (iii) make data available through a curated online database.

## Methods

We undertook a systematic review to identify all reports of *Leptospira* spp. identified to the serovar level detected in animal hosts. Our review followed Preferred Reporting Items for Systematic Reviews and Meta-Analyses (PRISMA) guidelines.(18) Although the International Prospective Register of Systematic Reviews (PROSPERO)(19) recently has allowed registration of systematic reviews of animal studies, this was not available at the start of our study in 2015. We searched nine databases: BIOSIS; CAB abstracts, including Aquaculture Compendium, CABI Full Text, Global Health and Leisure Tourism; Ovid Medline; PubMed; Proquest Agricultural Science Collection; Scopus; Web of Science; Wildlife and Ecology Studies Worldwide; and Zoological Record. Key search terms included ‘animal’, ‘*Leptospira’*, ‘serovar’, and a list of serovars (Text S1). A clinical librarian (SG) searched each database 28- 31 August 2015. The search was updated on 9 March 2023. Our search was not limited by year, publication type, or language. In addition, we searched the Amsterdam Medical Centre *Leptospira* library for additional articles listed for reference strain provenance.(3)

We included papers that reported *Leptospira* infection in an animal host, detected by culture or molecular testing, in which *Leptospira* was characterized to the serovar level by an acceptable method. Acceptable methods for identifying serovar included cross agglutinin absorption test (CAAT), monoclonal antibody typing, serum factor analysis, and pulsed-field gel electrophoresis (PFGE).(20–22) We included reports where typing methods were not explicitly stated but were performed at a reference laboratory and could be inferred to use the acceptable methods. We included only papers in which the common or scientific name of the animal was provided. We excluded papers in which animals had been experimentally infected and in which the isolate was not characterized to the serovar level.

First, a single author (MJM or NNH) reviewed all titles and abstracts for eligibility. For reports that passed abstract and title review, two independent reviewers (MJM and one of BC, ED, JH, KC, KS, MC, MPR, NNH, NS, SCG) reviewed the full-text. Final decision on eligibility was resolved through consensus or by decision of a third reviewer (JAC).

### Data abstraction

Data were abstracted into the database Airtable (Airtable, San Francisco, CA, United States) by MJM or NNH. We recorded: the period of sample collection; common and scientific name of the infected animal; country, number, age, and health status of the infected animal; the biologic sample from which *Leptospira* was detected; methods used for *Leptospira* detection and serovar determination; and serovar. For detections of serovar Hardjo, we also collected the subtype as Bovis, Prajitno, or not determined.

Publication year was grouped into 25-year periods. Geographic region was determined to the country level as defined by the United Nations (UN) and countries were grouped according to UN region and subregion.(23) We used the Integrated Taxonomic Information System to update and correct animal host scientific names that had changed from historic publications.(24) We inferred animal scientific names to either the genus or species level from common names for livestock species (e.g., *Bos taurus* for cattle), and for animal common names that described a single genus or species. Subsequently, all animals were grouped by animal order and animal class. The serogroup to which each serovar belonged was determined from existing reference lists.(3, 25)

### Data analysis

First, we described the publication year, country, and methods for serovar determination of the included reports. Second, we described the number of reported serovars and the number of unique animal host species with detected serovars. Third, we evaluated the animal host species range of detected serovars. For each serovar, we aggregated the number of detections by animal host identified to genus, species and order level, and visualised the serovar distribution by animal hosts using heat maps.

For the geographical distribution of serovar detections, we described the number of unique serovar detections by UN region and subregion. For each UN region and subregion, we counted the number of reports of animal detections per serovar. In addition, we counted the serovars that were detected in each UN region, and the serovars that were detected in a single region. The analysis of the geographical distribution of serovars were repeated for the three animal host orders with the largest number of reports.

Among countries with an estimated incidence of human leptospirosis >50 cases per 100,000 people,(9) we counted the number of reports of serotyped *Leptospira* detected in animal host species.

We generated summary tables of abstracted data of serovar detections per animal host species and geographic area, grouped by serogroup. In addition to tables, we developed an online searchable database named for Dr. Leopold Kirschner,(26, 27) a prominent leptospirosis researcher who worked at the University of Otago. Data analyses were performed in R version 4.2.

## Results

We identified 11,173 unique reports (Figure 1), with 1,862 (17.7%) included for full-text review after title and abstract screening. We completed full-text review of 1,697 (91.1%) reports, could not locate 79 (4.2%) reports, and were unable to translate 86 (4.6%) (Table S2, Text S3). Analysis included 409 reports: thirteen (3.2%) reports were published from 1927-1950, 158 (38.6%) from 1951-1975, 160 (39.1%) from 1976-2000, and 78 (19.1%) from 2001-2022 (Figure S5). For the reported methods of serovar detection, 265 (64.8%) reports used CAAT, 61 (14.9%) used monoclonal antibodies, 42 (10.3%) used PFGE, and 14 (3.4%) used serum factor analysis. For 66 (16.1%) reports, serovar detection was inferred to be performed by acceptable techniques at a reference laboratory. Multiple methods were used in 34 (8.1%) reports (Table S6).

**Figure 1.**
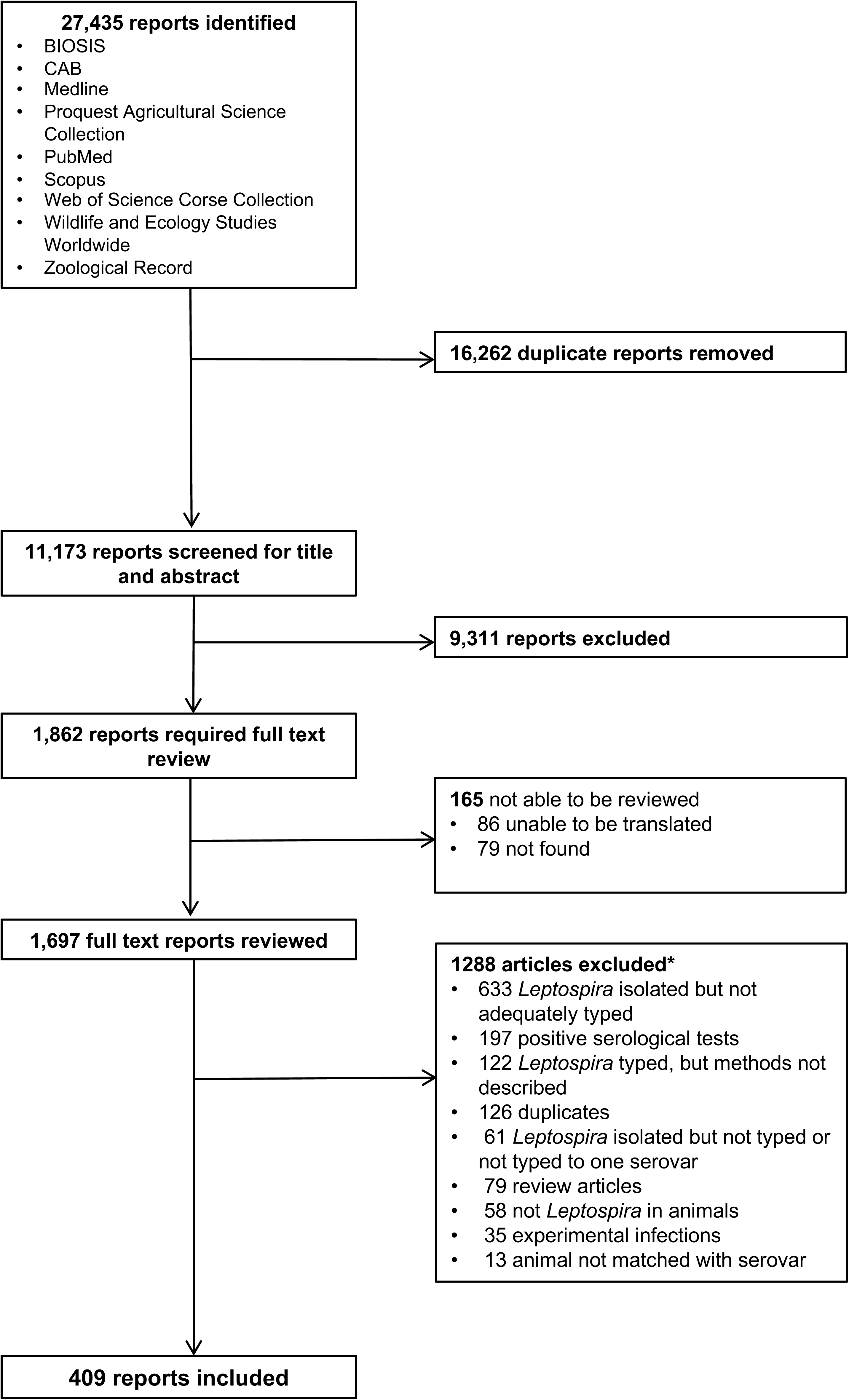
Flow diagram of the article review process for the systematic review of reports on *Leptospira* serovar isolations and detections in animal host species published 1927-2022 Legend: *Multiple reasons for exclusion possible

### Serovar detections across animal host species

The median (IQR) number of individual animal hosts contributing serotyped isolates was 4 (1–13) across 366 reports with available data. Overall, the 409 reports yielded data on 144 different animal hosts for which genus and species were identifiable, and an additional 6 animal hosts for which only the animal order could be identified. Among the 144 reported animal host species, 135 (93.8%) were from the class Mammalia, 5 (3.5%) from Amphibia, 3 (2.1%) from Reptilia, and 1 (0.7%) from Arachnida. Of the 16 identified animal host orders, Rodentia (e.g., rats) were described in 186 (45.5%) reports, Artiodactyla (e.g., cattle) in 148 reports (36.2%), Carnivora (e.g., dog) in 85 (20.8%) reports, Didelphimorphia (e.g., opossum) in 24 (5.9%) reports, and Eulipotyphla (e.g., shrew) in 19 (4.6%) reports. Other animal host orders are shown in Table S7. Animal age and animal health status were not provided, in 293 (71.6%) and 295 (72.1%) reports, respectively (Table S8).

Among the 409 reports, the median (IQR) number of detected serovars was 1 (1–2). Overall, 154 different serovars were reported from 25 serogroups. Five serovars could not be classified to the serogroup level. Serovars that were most frequently reported included Icterohaemorrhagiae (n=63), Pomona (n=61), Hardjo (n=54), Canicola (n=51), Grippotyphosa (n=48), and Ballum (n=40) (Figure 2A). Among 53 reports that detected serovar Hardjo, 11 included serovar Hardjo subtype Bovis, 6 included serovar Hardjo subtype Prajitno, and in 42 Hardjo subtype was not further determined. Across 144 animal host species with an identifiable genus and species, the serovars detected in the largest number of different animal species included Grippotyphosa (n=39), Icterohaemorrhagiae (n=29), Pomona (n=28), Australis (n=25), and Ballum (n=25) (Figure 2B). Of serovars, 76 were detected in one animal host species, and 32 serovars in two animal host species. Among animal host species, the largest number of serovars were detected in *Bos taurus* (n=43), *Rattus rattus* (n=30), *Mus musculus* (n=23), *Sus scrofa* (n=23), *Canis lupus* (n=21), and *Rattus norvegicus* (n=20) (Figure S8).

**Figure 2.**
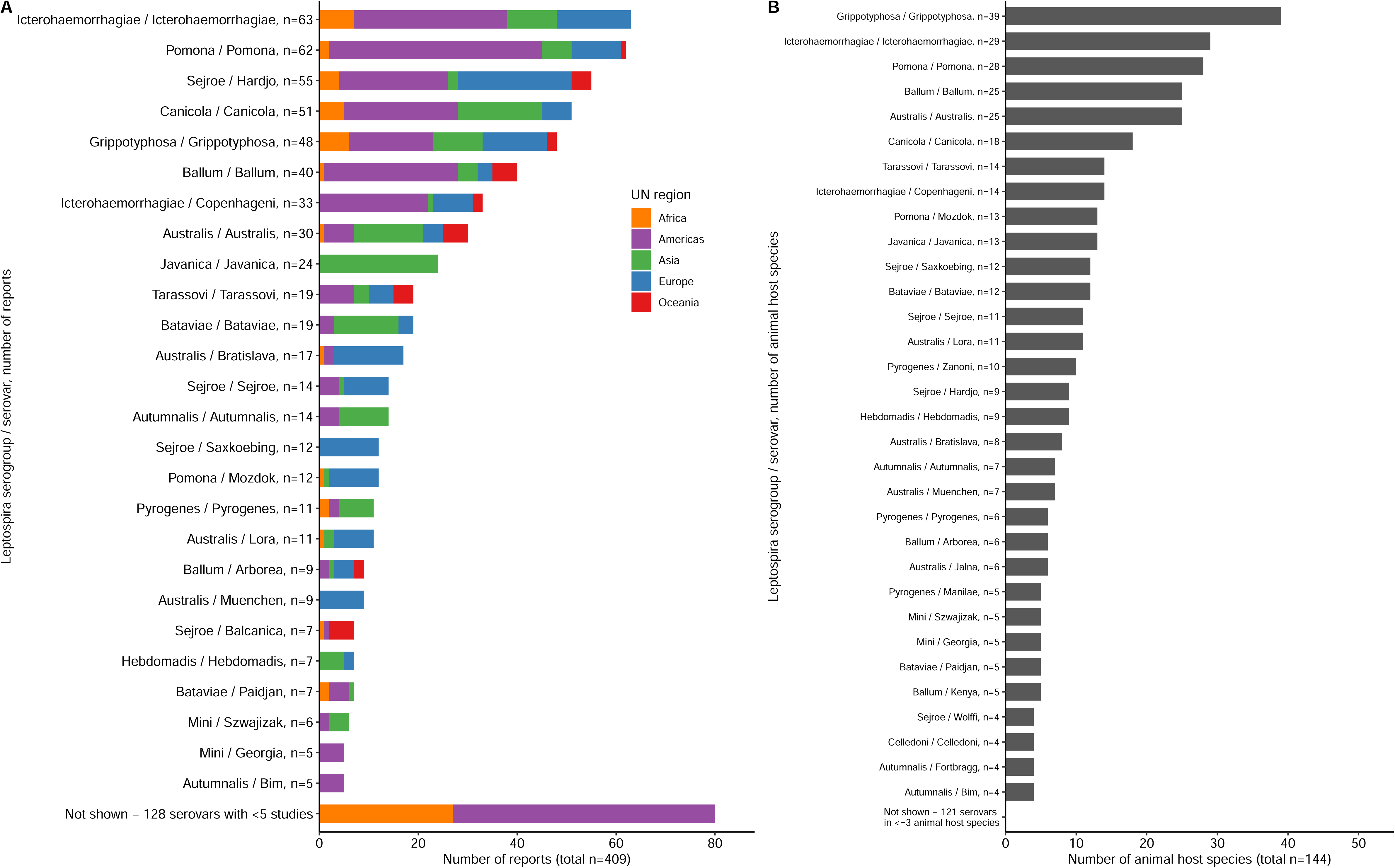
Frequency of number of reports per *Leptospira* serovar detected from animal hosts, per UN region (A), and frequency of unique animal host species in which each *Leptospira* serovar was detected (B) in the systematic review of reports on *Leptospira* serovar isolations and detections in animal host species published 1927-2022 Legend: A: Reports that described different regions are counted multiple times. Serovar Pomona was described in 62 reports; serovar Hardjo was described in 53 reports.

The number of detected serovars ranged from 1 through 90 across the 16 animal host orders: 90 serovars in Rodentia, 58 in Artiodactyla, 34 in Carnivora, 31 in Didelphimorphia, 17 in Eulipotyphla, and 10 or less serovars in the other 11 animal host orders (Figure 3). Serovar Grippotyphosa was detected in 10 animal host orders. 95 serovars were detected in a single animal host order, and 33 serovars in two animal host orders. The median (IQR) number of animal host orders in which the remaining 25 serovars were detected was 4 (3–5).

**Figure 3.**
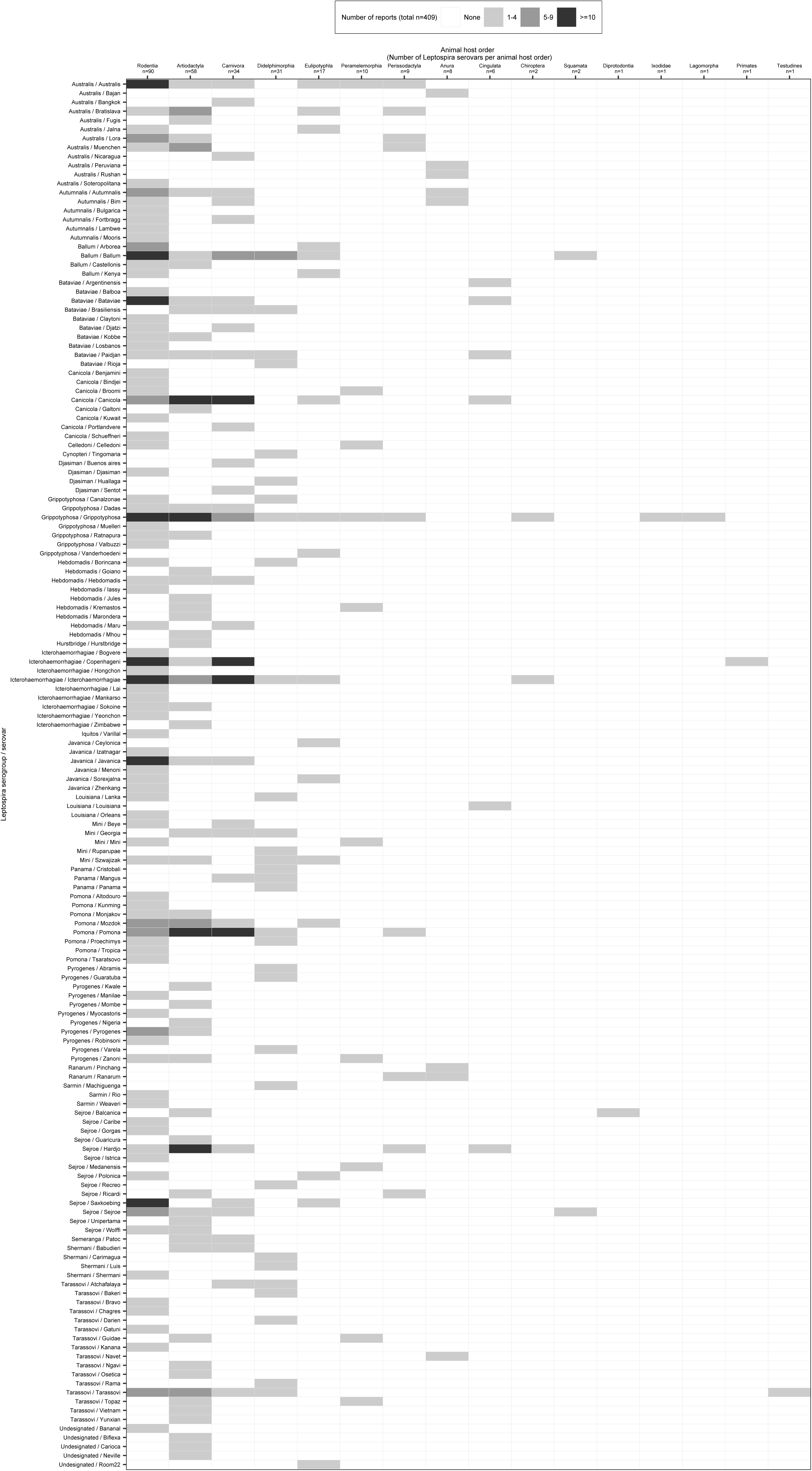
Heat map of detected *Leptospira* serovars in 16 animal host orders in the systematic review of reports on *Leptospira* serovar isolations and detections in animal host species published 1927-2022

### Serovar geographical distribution

Of 409 included reports, 28 (6.9%) were from Africa, 169 (41.3%) from the Americas, 90 (22.0%) from Asia, 103 (25.2%) from Europe, 23 (5.6%) from Oceania, and none from Antarctica (Table 1, Figure S4). UN subregions lacking reported data were Middle Africa and Micronesia. The reports included data from 66 (26.5%) of 249 countries, including 9 (15.0%) of 60 countries in Africa, 17 (29.8%) of 57 countries in the Americas, 15 (30.0%) of 50 countries in Asia, 21 (40.4%) of 52 countries in Europe, and 4 (13.8%) of 29 countries in Oceania (Table 1). The number of detected serovars varied by region: 29 serovars were detected in Africa, 86 serovars in the Americas, 54 serovars in Asia, 36 serovars in Europe, and 20 serovars in Oceania (Table 1). Serovars detected at least once from each of the five UN regions included serovars Australis, Ballum, Grippotyphosa, Hardjo, and Pomona. Of 117 serovars that were detected in a single UN region, 13 (11.1%) were detected in Africa, 60 (51.3%) in the Americas, 27 (23.1%) in Asia, 10 (8.5%) in Europe, and 7 (5.9%) in Oceania.

**Table 1.** Geographical distribution and animal host range of reports included in the systematic review for the isolations and detections of *Leptospira* serovars, published 1927-2022.

| UN region and subregions<sup>^</sup> | Number of reports, % of total reports* |  | Number of countries/total number of countries, % ± |  | Number of unique serovars | Serovars with rank order 1, 2 or 3 for number of reports, ordered by frequency<br>(number of reports) | Number of unique animal orders | Animal host orders with rank order 1, 2 or 3 for number of reports, ordered by frequency<br>(number of reports) |
| --- | --- | --- | --- | --- | --- | --- | --- | --- |
|  | n (%) |  | n/n (%) |  | n |  | n |  |
| **Overall** | **409** | **(100)** | **66/249** | **(26.5)** | **154** | **Icterohaemorrhagiae (63); Pomona (61); Hardjo (54)** | **16** | **Rodentia (186); Artiodactyla (148); Carnivora (85)** |
| **Africa<sup>^</sup>** | **28** | **(6.9)** | **9/60** | **(15.0)** | **29** | **Icterohaemorrhagiae (7); Grippotyphosa (6); Canicola (5)** | **4** | **Artiodactyla (17); Rodentia (12); Carnivora (3)** |
| Eastern Africa | 14 | (3.4) | 4/22 | (18.2) | 22 | Kenya (3); [11 serovars] (2)# | 3 | Artiodactyla (10); Rodentia (5); Eulipotyphla (1) |
| Northern Africa | 6 | (1.5) | 3/7 | (42.9) | 6 | Icterohaemorrhagiae (4); Grippotyphosa (3); Canicola (1); Patoc (1); Polonica (1); Pyrogenes (1) | 3 | Rodentia (5); Carnivora (2); Artiodactyla (1) |
| Southern Africa | 5 | (1.2) | 1/5 | (20.0) | 4 | Canicola (2); Hardjo (1); Icterohaemorrhagiae (1); Pomona (1) | 3 | Artiodactyla (3); Carnivora (1); Rodentia (1) |
| Western Africa | 3 | (0.7) | 1/17 | (5.9) | 10 | [10 serovars] (1) | 2 | Artiodactyla (3); Rodentia (1) |
| **The Americas** | **169** | **(41.3)** | **17/57** | **(29.8)** | **86** | **Pomona (43); Icterohaemorrhagiae (31); Ballum (27)** | **13** | **Rodentia (66); Artiodactyla (60); Carnivora (52)** |
| Caribbean | 18 | (4.4) | 6/28 | (21.4) | 26 | Copenhageni (8); Bim (5); Ballum (4); Icterohaemorrhagiae (4) | 5 | Rodentia (11); Carnivora (10); Anura (5) |
| Central America | 4 | (1.0) | 3/8 | (37.5) | 24 | [24 serovars] (1)# | 3 | Didelphimorphia (3); Rodentia (3); Carnivora (2) |
| Northern America | 82 | (20.0) | 2/5 | (40.0) | 23 | Pomona (29); Ballum (18); Hardjo (15); Icterohaemorrhagiae (15) | 11 | Artiodactyla (31); Carnivora (31); Rodentia (23) |
| Southern America | 65 | (15.9) | 6/16 | (37.5) | 48 | Pomona (14); Copenhageni (12); Icterohaemorrhagiae (12) | 8 | Rodentia (29); Artiodactyla (27); Carnivora (9) |
| **Asia** | **90** | **(22.0)** | **15/50** | **(30.0)** | **54** | **Javanica (24); Canicola (17); Australis (14)** | **7** | **Rodentia (50); Artiodactyla (24); Carnivora (19)** |
| Central Asia | 1 | (0.2) | 1/5 | (20.0) | 1 | Grippotyphosa (1) | 1 | Ixodidae (1) |
| Eastern Asia | 25 | (6.1) | 3/7 | (42.9) | 23 | Australis (5); Canicola (4); Hebdomadis (4); Icterohaemorrhagiae (4); Javanica (4); Lai (4) | 4 | Rodentia (16); Carnivora (6); Artiodactyla (4) |
| Southern Asia | 22 | (5.4) | 5/11 | (45.5) | 14 | Javanica (7); Canicola (4); Grippotyphosa (3) | 4 | Rodentia (13); Artiodactyla (7); Carnivora (4) |
| South-Eastern Asia | 31 | (7.6) | 3/9 | (33.3) | 29 | Javanica (13); Bataviae (12); Australis (8) | 4 | Rodentia (15); Artiodactyla (11); Carnivora (8) |
| Western Asia | 11 | (2.7) | 3/18 | (16.7) | 11 | Szwajizak (4); Ballum (3); Canicola (3); Grippotyphosa (3) | 4 | Rodentia (6); Eulipotyphla (4); Artiodactyla (2) |
| **Europe** | **103** | **(25.2)** | **21/52** | **(40.4)** | **36** | **Hardjo (23); Icterohaemorrhagiae (15); Bratislava (14)** | **6** | **Rodentia (48); Artiodactyla (42); Carnivora (10); Eulipotyphla** |
| Eastern Europe | 29 | (7.1) | 7/10 | (70.0) | 25 | Grippotyphosa (6); Pomona (5); Sejroe (5); Tarassovi (5) | 5 | Rodentia (20); Artiodactyla (7); Eulipotyphla (4) |
| Northern Europe | 35 | (8.6) | 3/17 | (17.6) | 14 | Hardjo (18); Bratislava (8); Muenchen (8) | 5 | Artiodactyla (26); Rodentia (9); Perissodactyla (5) |
| Southern Europe | 20 | (4.9) | 4/16 | (25.0) | 22 | Icterohaemorrhagiae (7); Mozdok (5); Arborea (4) | 5 | Rodentia (11); Carnivora (4); Artiodactyla (3) |
| Western Europe | 21 | (5.1) | 7/9 | (77.8) | 15 | Grippytyphosa (5); Copenhageni (4); Lora (4) | 5 | Rodentia (8); Artiodactyla (6); Carnivora (3);<br>Perissodactyla (3) |
| **Oceania^** | **23** | **(5.6)** | **4/29** | **(13.8)** | **20** | **Australis (5); Balcanica (5); Ballum (5)** | **6** | **Rodentia (10); Artiodactyla (9); Diprotodontia (5)** |
| Australia and<br>New Zealand | 21 | (5.1) | 2/6 | (33.3) | 19 | Balcanica (5); Ballum (5); Australis (4); Hardjo (4);<br>Tarassovi (4); Zanoni (4) | 6 | Artiodactyla (9); Rodentia (9); Diprotodontia (5) |
| Melanesia | 1 | (0.2) | 1/5 | (20.0) | 1 | Guidae (1) | 1 | Peramelemorphia (1) |
| Polynesia | 1 | (0.2) | 1/10 | (10.0) | 1 | Australis (1) | 1 | Rodentia (1) |

^No data reported from Middle Africa (9 countries), Micronesia (8 countries), or Antarctica (1 country). \*The number of reports exceeds the total number of reports (n=409) since some reported data from multiple regions or subregions. ±Some reports included data from multiple countries. #Eastern Africa: 11 serovars that had rank order 2; Central America: 24 serovars that had rank order 1

Of 27 countries with an estimated annual incidence of leptospirosis >50 per 100,000 people,(9) 6 (22.2%) countries had available reports of *Leptospira* detection from an animal host species and 21 (77.8%) had no report. Across the six countries with reports, the median (IQR) number of reports was 2 (1–4) (Table S9). Of the 21 countries with no reports, five (Comoros, Mauritius, Rwanda, Sao Tome and Principe, Seychelles) were in Africa, five (Antigua and Barbuda, Jamaica, Saint Kitts and Nevis, Saint Lucia, Saint Vincent, and the Grenadines) in the Americas, two (Maldives, Timor-Leste) in Asia, and nine (Fiji, Kiribati, Federal states of Micronesia, Palau, Samoa, Solomon Islands, Tonga, Tuvalu, Vanuatu) in Oceania.

The largest number of reports were on animal host order Rodentia, Artiodactyla, and Carnivora. Of 90 serovars detected in hosts of the order Rodentia (Figure 4), serovars detected at least once from each UN region were serovars Australis, Ballum, and Grippotyphosa. Of 58 serovars detected in hosts of order Artiodactyla, serovars detected at least once from each UN region included serovars Grippotyphosa, Hardjo, and Pomona. The distribution of serovar Hardjo subtypes among Artiodactyla order hosts was subtype Bovis in four regions (Africa, the Americas, Europe, and Oceania), subtype Prajitno in two regions (Europe, the Americas), and subtype not determined in all five regions. Of 34 serovars detected in hosts of order Carnivora, no serovars were detected in all five UN regions.

**Figure 4.**
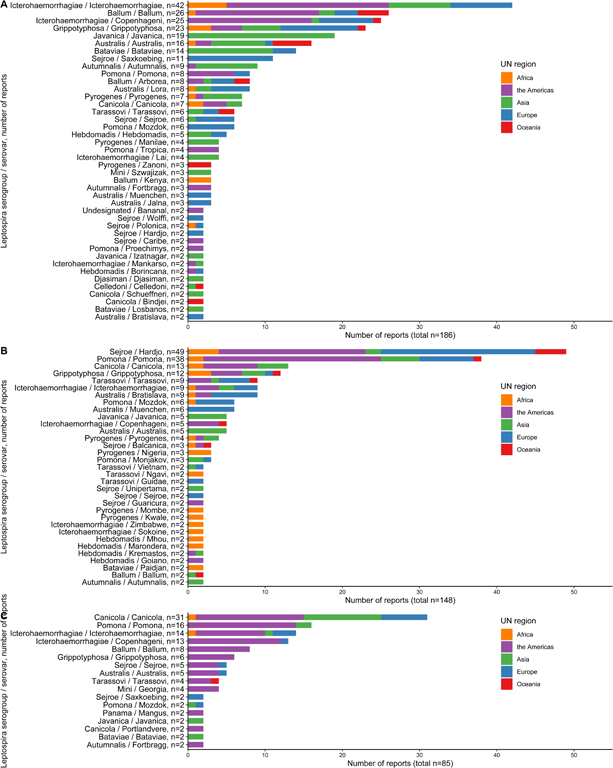
Frequency of number of reports that detected *Leptospira* serovars per UN region, stratified for Rodentia (A), Artiodactyla (B), and Carnivora (C) in the systematic review of reports on *Leptospira* serovar isolations and detections in animal host species published 1927-2022 Legend: *Serovars that were detected in only one report are not shown.

Serogroups from which all serovars were isolated in animal hosts from a single UN subregion included serogroups Cynopteri and Iquitos reported only from South America (Peru), and serogroup Hurstbridge reported only from Australia and New Zealand (Australia).

### Online database

The animal host species and country of origin for serovars are shown in Table S10 and the curated online database can be found at https://www.otago.ac.nz/internationalhealth/research/leptospirosis/[this is a pilot version, the final version will be available upon publication]. The database is searchable by serovar, serogroup, country, animal scientific name, and animal common name.

## Discussion

With this systematic review, we synthesize the literature on *Leptospira* serovar isolations and detections in animal host species and make these data available through a curated online database. This updated *Leptospira* serovar database on serovar animal host range could aid investigations of and response to leptospirosis outbreaks,(10) and inform leptospirosis control programs by suggesting which animal host species might be the source of infection. For example, in a human population in rural Rwanda, Ntabanganhiman*et al.* found a high prevalence of *Leptspira* in human samples.(28) Of these, MAT results suggested that mosts infections were caused by serogroup *Icterohaemorrhagiae*, but the potential animal source of infection was unclear and there are not typed*Leptospira* isolates from Rwanda. Our systematic review found that serovars from the Icterohaemorrhagiae serogroup have been isolated from pouched rats and cattle in countries neigbouring Rwanda, providing hypotheses regarding possible host animal sources. (29–32) In addition, our database may facilitate detection of cases of human leptospirosis by informing serovar composition of MAT antigen panels.

Our systematic review used a rigorous process and included published reports that were indexed in the searched databases and used acceptable methods for serovar determination. We identified serovar detections in animal hosts in 66 countries. Previous reviews in 1966 and the update in 1975 reported detections in animals in 71 countries respectively, but were based on unpublished data and personal communications.(13, 14) Since country borders and country names have changed over the period in which included reports were published, a comparison of number of countries was not possible.

Overall, *Leptospira* detections were reported from a small proportion of all known animal species. Serotyped *Leptospira* detections were predominantly from mammals, but among mammals, we found *Leptospira* detections from <2% of the approximately 6,000 currently described mammalian species.(24) There were also reports of detection from reptiles and amphibians, and one report of detection from arachnids. We did not identify reports of detection of adequately typed *Leptospira* from birds or fish.(33, 34) Some *Leptospira* serovars were detected from a limited number of animal species whereas others were detected from a diversity of animal species. For instance, serovar Grippotyphosa showed the largest host range with detections across 39 different animal host species from 10 animal host orders. On the other hand, some frequently reported serovars were detected in one or two animal species. For example, serovar Hardjo was mainly detected in cattle, and serovar Canicola in dogs. More than 100 serovars were detected in only one or two animal species. It is possible that human *Leptospira* infection with a serovar that has a narrow host range implicates the likely source of infection. However, variation in the number ofserovar detections by animal species was potentially influenced by differences in host susceptibility, and lack of research on many animal species. Our results should be interpreted with caution and the lack of identification of a serovar from any particular animal species should not be interpreted as an inability to infect that species.

We have confirmed that *Leptospira* are geographically widespread, with typed isolates detected from animal hosts in every continent except Antarctica. Despite this, there are many countries with few or no serovar detections, including countries in Africa and Oceania thought to have a high incidence of human leptospirosis.(9) It is likely that the absence of information reflects a lack of research rather than an absence of animal *Leptospira* infection in those countries. We note that many countries with a high incidence of human leptospirosis that lacked data on *Leptospira* detection from animal hosts are island nations.(9, 15) Importantly, each island might have local variation in factors influencing leptospirosis epidemiology including biodiversity, ecosystem, and human-animal interactions.

There were several serovars and serogroups reported from a limited geographic range, while others had a broader geographic distribution. An example of a serovar with a limited geographic range is serovar Saxkoebing, detected exclusively in Europe. However, as it was detected in mice, voles and dogs, which are widespread in other regions, it is possible that the geographic range of serovar Saxkoebing is wider than what is currently reported. On the other hand, serovars Australis, Ballum, Grippotyphosa, Hardjo, and Pomona were identified in all five UN regions, suggesting wider geographic distribution.

Of the >250 serovars of *Leptospira* recorded,(20) we identified reports of detection of 154 serovars from animal hosts. Data on detection from an animal species are lacking for many serovars. For many others, we found just a single report of detection in an animal species. Potential explanations include highly localized epidemiology of particular serovars, occasional exposure of animals to serovars for which the environment acts as a reservoir, variation in host range for some serovars, or insufficient research and reporting of animal *Leptospira* infection in many areas.

Our study has some limitations. We cannot establish if the absence of a serovar from a location or animal is a result of a report being missed by our systematic review process; or not being sought, detected, or reported by researchers; or not being present. *Leptospira* culture is insensitive and shedding of bacteria by host animals can be intermittent.(35, 36) Loss of provenance of isolates is also a limitation of our study, as detailed typing of isolates by reference laboratories may be performed without citing the origin of the isolate. It is also likely that there has been misclassification of reported isolates, particularly as serovar classification has undergone several major changes, with the alteration in the definition of serovar, and renaming of serovars.(4, 20, 37) Early serotype determinations were not standardized and the reference standard for most of the 20^th^ century, CAAT, is a complex test to perform. As evidence of these difficulties, we identified published reports in which authors identified errors in their own earlier classification of isolates.(38, 39) Lastly, we did not perform a bias assessment of the included reports as we focused solely on serovar detections of animal hosts and only selected reports that used appropriate methods of serovar detections.

## Conclusion

In conclusion, we have performed a systematic review on *Leptospira* serovar isolations and detections from animal host species worldwide using contemporary systematic review methods and made them available in a curated publicly accessible database. In so doing, we have identified that there are few published data on *Leptospira* serovars from animal host species from multiple countries considered to have a high incidence of human leptospirosis, especially those in Africa and Oceania. We anticipate that our updated review and database will provide a useful resource for public health practitioners and leptospirosis researchers, and inform efforts to control and prevent leptospirosis.

## Data Availability

All data produced in the present study are available upon reasonable request to the authors. Upon publication the data will be available in an online database.

## Acknowledgments

The authors would like to thank University of Otago librarians, particularly Jacinda Boivin, Jessie Neilson, Susan Pennock, Dierdre O’Neill, Jan Wyllie and Tanya Low; as well as colleagues at libraries around the world who helped collect the full-text articles for review. The authors would also like to thank Gary Niemi, Geoffrey Hughes, Louis Prior, and Kevin Maeer, Applications Development Team, Information Systems Group, ITS, University of Otago, for development of the online database.

## Disclaimer

The findings and conclusions in this report are those of the authors and do not necessarily represent the official position of the US Centers for Disease Control and Prevention. Use of trade names and commercial sources is for identification only and does not imply endorsement by the US Department of Health and Human Services or the Centers for Disease Control and Prevention.

## Data availability statement

Data underling the study can be accessed in the curated online database (https://www.otago.ac.nz/internationalhealth/research/leptospirosis/). For other data requests or questions contact the corresponding author.

## Supporting information captions

**Text S1.** Searches performed during the systematic review of reports on *Leptospira* serovar isolations and detections in animal host species, 9 March 2023

**Table S2.** Publication language of reports selected for full-text review during the systematic review of reports on Leptospira serovar isolations and detections in animal **host species**

**Text S3.** Reports identified until 9 March 2023 which met criteria for full-text review but could not be reviewed in the systematic review of reports on *Leptospira* serovar isolations and detections in animal host species

**Figure S4.** Global frequency of number of reports per country identified in the systematic review of reports on *Leptospira* serovar isolations and detections in animal host species published 1927-2022

**Figure S5.** Histogram of publication years of reports in the systematic review of reports on Leptospira serovar isolations and detections in animal host species published 1927-2022

**Table S6.** Characteristics of reports included in the systematic review of reports on *Leptospira* serovar isolations and detections in animal host species published 1927-2022

**Table S7.** Number of reports of Leptospira serovar detected per animal host order in the systematic review of reports on Leptospira serovar isolations and detections in animal host species published 1927-2022

**Figure S8.** Heat map of detected *Leptospira* serovar in 16 animal host orders per animal host genus in the systematic review of reports on *Leptospira* serovar isolations and detections in animal host species published 1927-2022 (409 reports) (A, Rodentia; B, Artiodactyla; C, Carnivora; D, Didelphimorphia; E, Eulipotyphla; F, Perissodactyla; G, Anura; H, Peramelemorphia; I, Cingulata; J, Squamata; K, Chiroptera; L, 5 animal orders with 1 serovar (Diprotodontia, Ixodidae, Lagomorpha, Primates, Testudines)

**Table S9.** Table of estimated human leptospirosis morbidity and mortality by Costa *et al.* and number of reports, number of serovars, and number of animal hosts identified in the systematic review of reports on *Leptospira* serovar isolations and detections in animal host species published 1927-2022

**Table S10.** Summary tables of *Leptospira* serovar isolations and detections in animal host species published 1927-2022, sorted by serogroup, serovar, and animal genus and species

**Table S11.** PRISMA checklist

