## Supplemental file for "Global distribution of *Leptospira* serovar isolations and detections from animal host species: a systematic review and online database"

#### Supporting Information

#### Text S1. Searches performed during the systematic review of reports on *Leptospira* serovar isolations and detections in animal host species, 9 March 2023

Databases:

Medline, Scopus, Web of Science, BIOSIS, CAB abstracts, Wildlife and ecology studies worldwide, Proquest Agricultural Science Collection, Zoological record, PubMed

Search as performed in Web of science: BIOSIS, Medline, Web of Science Core collection, Zoological record

| Search | Terms Used |
|---|---|
| 1 | TS=((Abramis) OR (Agogo) OR (Aguaruna) OR (Alexi) OR (Alice) OR (Altodouro) OR (Andamana) OR (Anhoa) OR (Arborea) OR (Arenal) OR (Argentinensis) OR (Atchafalaya) OR (Atlantae) OR (Australis) OR (Autumnalis) OR (Babudieri) OR (Bafani) OR (Bajan) OR (Bakeri) OR (Balboa) OR (Balcanica) OR (Ballum) OR (Bangkinang) OR (Banna) OR (Bataviae) OR (Benjamini) OR (Beye) OR (Biggis) OR (Bim) OR (Bindjei) OR (Birkini) OR (Bogvere) OR (Borincana) OR (Brasiliensis) OR (Bratislava) OR (Bravo) OR (Broomi) OR (Buenos Aires) OR (Bulgarica) OR (Butembo) OR (Camlo) OR (Canalzonae) OR (Canicola) OR (Caribe) OR (Carimagua) OR (Carlos) OR (Castellonis) OR (Celledoni) OR (Ceylonica) OR (Chagres) OR (Claytoni) OR (Copenhageni) OR (Copenhageni) OR (Corredores) OR (Costa Rica) OR (Coxi) OR (Cristobali) OR (Cuica) OR (Cynopteri) OR (Dadas) OR (Dakota) OR (Darien) OR (Dehong) OR (Dikkeni) OR (Djasiman) OR (Djatzi) OR (Erinaceiauriti) OR (Evansi) OR (Fluminense) OR (Fortbragg) OR (Fugis) OR (Galtoni) OR (Gatuni) OR (Gem) OR (Gengma) OR (Georgia) OR (Geyaweera) OR (Goiano) OR (Gorgas) OR (Grippotyphosa) OR (Guangdong) OR (Guaratuba) OR (Guaricura) OR (Guidae) OR (Gurungi) OR (Haemolytica) OR (Hainan) OR (Hamptoni) OR (Hardjo) OR (Hawain) OR (Hebdomadis) OR (Hekou) OR (Holland) OR (Hongchon) OR (Hualin) OR (Huallaga) OR (Huanuco) OR (Hurstbridge) OR (Icterohaemorrhagiae) OR (Istrica) OR (Jalna) OR (Javanica) OR (Jonsis) OR (Jules) OR (Kabura) OR (Kambale) OR (Kamituga) OR (Kanana) OR (Kaup) OR (Kenya) OR (Khorat) OR (Kisuba) OR (Kobbe) OR (Kremastos) OR (Kunming) OR (Kuwait) OR (Kwale) OR (Lai) OR (Lambwe) OR (Langati) OR (Lanka) OR (Lichuan) OR (Lincang) OR (Lora) OR (Losbanos) OR (Louisiana) OR (Luis) OR (Machiguenga) OR (Malaya) OR (Malaysia) OR (Mangus) OR (Manhao) OR (Manilae) OR (Mankarso) OR (Manzhuang) OR (Maru) OR (Medanensis) OR (Mengding) OR (Mengla) OR (Menglian) OR (Mengma) OR (Mengpeng) OR (Mengrun) OR (Menoni) OR (Mini) OR (Mogden) OR (Mooris) OR (Mozdok) OR (Muelleri) OR (Muenchen) OR (Mujunkumi) OR (Mwogolo) OR (Myocastoris) OR (Naam) OR (Nanla) OR (Navet) OR (Ndahambukuje) OR (Ndambari) OR (Nicaragua) OR (Nigeria) OR (Nona) OR (Nyanza) OR (Orleans) OR (Paidjan) OR (Panama) OR (Patoc) OR (Perameles) OR (Peru) OR (Peruviana) OR (Pina) OR (Pinchang) OR (Poi) OR (Polonica) OR (Pomona) OR (Portblairi) OR (Portlandvere) OR |

| | |
|---|---|
| | (Princestown) OR (Proechimys) OR (Pyrogenes) OR (Qingshui) OR (Rachmati) OR (Rama) OR (Ramisi) OR (Ranarum) OR (Ratnapura) OR (Recreo) OR (Ricardi) OR (Rio) OR (Rioja) OR (Robinsoni) OR (Roumanica) OR (Ruparupae) OR (Rushan) OR (Sanmartini) OR (Santarosa) OR (Saopaulo) OR (Sarmin) OR (Saxkoebing) OR (Schueffneri) OR (Sejroe) OR (Semaranga) OR (Sentot) OR (Shermani) OR (Sichuan) OR (Smithi) OR (Sofia) OR (Sokoine) OR (Sorexjalna) OR (Soteropolitana) OR (Srebarna) OR (Sulzerae) OR (Sumneri) OR (Szwajizak) OR (Tabaquite) OR (Tarassovi) OR (Tingomaria) OR (Tonkini) OR (Topaz) OR (Trinidad) OR (Tropica) OR (Tsaratsovo) OR (Tunis) OR (Valbuzzi) OR (Vanderhoedeni) OR (Varela) OR (Vargonicas) OR (Varillal) OR (Vughia) OR (Waskurin) OR (Weaveri) OR (Weerasinghe) OR (Whitcombi) OR (Wolffi) OR (Worsfoldi) OR (Yaan) OR (Yeonchon) OR (Yunnan) OR (Yunxian) OR (Zanoni) OR (Zhenkang) OR (Serovar) OR (Serogroup) OR (Serotype)) |
| 2 | TS=((Animal*) OR (mammal) OR (reptile) OR (bird) OR (avian) OR (amphibian))<br><br>Timespan=All years |
| 3 | TS=Leptospir*<br>Timespan=All years |
| 1 | 1 AND 2 AND 3 |

Search as performed in CAB abstracts

| Search | Search terms |
|---|---|
| 1 | Leptospir* AND ((Animal*) OR (mammal) OR (reptile) OR (bird) OR (avian) OR (amphibian)) AND ((Abramis) OR (Agogo) OR (Aguaruna) OR (Alexi) OR (Alice) OR (Altodouro) OR (Andamana) OR (Anhoa) OR (Arborea) OR (Arenal) OR (Argentinensis) OR (Atchafalaya) OR (Atlantae) OR (Australis) OR (Autumnalis) OR (Babudieri) OR (Bafani) OR (Bajan) OR (Bakeri) OR (Balboa) OR (Balcanica) OR (Ballum) OR (Bangkinang) OR (Banna) OR (Bataviae) OR (Benjamini) OR (Beye) OR (Biggis) OR (Bim) OR (Bindjei) OR (Birkini) OR (Bogvere) OR (Borincana) OR (Brasiliensis) OR (Bratislava) OR (Bravo) OR (Broomi) OR (Buenos Aires) OR (Bulgarica) OR (Butembo) OR (Camlo) OR (Canalzonae) OR (Canicola) OR (Caribe) OR (Carimagua) OR (Carlos) OR (Castellonis) OR (Celledoni) OR (Ceylonica) OR (Chagres) OR (Claytoni) OR (Copenhageni) OR (Copenhageni) OR (Corredores) OR (Costa Rica) OR (Coxi) OR (Cristobali) OR (Cuica) OR (Cynopteri) OR (Dadas) OR (Dakota) OR (Darien) OR (Dehong) OR (Dikkeni) OR (Djasiman) OR (Djatzi) OR (Erinaceiauriti) OR (Evansi) OR (Fluminense) OR (Fortbragg) OR (Fugis) OR (Galtoni) OR (Gatuni) OR (Gem) OR (Gengma) OR (Georgia) OR (Geyaweera) OR (Goiano) OR (Gorgas) OR (Grippotyphosa) OR (Guangdong) OR (Guaratuba) OR (Guaricura) OR (Guidae) OR (Gurungi) OR (Haemolytica) OR (Hainan) OR (Hamptoni) OR (Hardjo) OR (Hawain) OR (Hebdomadis) OR (Hekou) OR (Holland) OR (Hongchon) OR (Hualin) OR (Huallaga) OR (Huanuco) OR (Hurstbridge) OR (Icterohaemorrhagiae) OR (Istrica) OR (Jalna) OR (Javanica) OR (Jonsis) OR (Jules) OR (Kabura) OR (Kambale) OR (Kamituga) OR (Kanana) OR (Kaup) OR (Kenya) OR (Khorat) OR (Kisuba) OR (Kobbe) OR (Kremastos) OR (Kunming) OR (Kuwait) OR (Kwale) OR (Lai) OR (Lambwe) OR (Langati) OR (Lanka) OR (Lichuan) OR (Lincang) OR (Lora) OR (Losbanos) OR (Louisiana) OR (Luis) OR (Machiguenga) OR (Malaya) OR (Malaysia) OR (Mangus) OR (Manhao) OR (Manilae) OR (Mankarso) OR (Manzhuang) OR (Maru) OR (Medanensis) OR (Mengding) OR (Mengla) OR (Menglian) OR (Mengma) OR (Mengpeng) OR (Mengrun) OR (Menoni) OR (Mini) OR (Mogden) OR (Mooris) OR (Mozdok) OR (Muelleri) OR (Muenchen) OR (Mujunkumi) OR (Mwogolo) OR (Myocastoris) OR (Naam) OR (Nanla) OR (Navet) OR (Ndahambukuje) OR (Ndambari) OR (Nicaragua) OR (Nigeria) OR (Nona) OR (Nyanza) OR (Orleans) OR (Paidjan) OR (Panama) OR (Patoc) OR (Perameles) OR (Peru) OR (Peruviana) OR (Pina) OR (Pinchang) OR (Poi) OR (Polonica) OR (Pomona) OR (Portblairi) OR (Portlandvere) OR (Princestown) OR (Proechimys) OR (Pyrogenes) OR (Qingshui) OR (Rachmati) OR (Rama) OR (Ramisi) OR (Ranarum) OR (Ratnapura) OR (Recreo) OR (Ricardi) OR (Rio) OR (Rioja) OR (Robinsoni) OR (Roumanica) OR (Ruparupae) OR (Rushan) OR (Sanmartini) OR (Santarosa) OR (Saopaulo) OR (Sarmin) OR (Saxkoebing) OR (Schueffneri) OR (Sejroe) OR (Semaranga) OR (Sentot) OR (Shermani) OR (Sichuan) OR (Smithi) OR (Sofia) OR (Sokoine) OR (Sorexjalna) OR (Soteropolitana) OR (Srebarna) OR (Sulzerae) OR (Sumneri) OR (Szwajizak) OR (Tabaquite) OR (Tarassovi) OR (Tingomaria) OR (Tonkini) OR |

| | |
|---|---|
| | (Topaz) OR (Trinidad) OR (Tropica) OR (Tsaratsovo) OR (Tunis) OR (Valbuzzi) OR (Vanderhoedeni) OR (Varela) OR (Vargonicas) OR (Varillal) OR (Vughia) OR (Waskurin) OR (Weaveri) OR (Weerasinghe) OR (Whitcombi) OR (Wolffi) OR (Worsfoldi) OR (Yaan) OR (Yeonchon) OR (Yunnan) OR (Yunxian) OR (Zanoni) OR (Zhenkang) OR (Serovar) OR (Serogroup) OR (Serotype)) |

Search performed in Proquest Agricultural Science Collection

| Search | Search Terms |
|---|---|
| 1 | ((Abramis) OR (Agogo) OR (Aguaruna) OR (Alexi) OR (Alice) OR (Altodouro) OR (Andamana) OR (Anhoa) OR (Arborea) OR (Arenal) OR (Argentinensis) OR (Atchafalaya) OR (Atlantae) OR (Australis) OR (Autumnalis) OR (Babudieri) OR (Bafani) OR (Bajan) OR (Bakeri) OR (Balboa) OR (Balcanica) OR (Ballum) OR (Bangkinang) OR (Banna) OR (Bataviae) OR (Benjamini) OR (Beye) OR (Biggis) OR (Bim) OR (Bindjei) OR (Birkini) OR (Bogvere) OR (Borincana) OR (Brasiliensis) OR (Bratislava) OR (Bravo) OR (Broomi) OR (Buenos Aires) OR (Bulgarica) OR (Butembo) OR (Camlo) OR (Canalzonae) OR (Canicola) OR (Caribe) OR (Carimagua) OR (Carlos) OR (Castellonis) OR (Celledoni) OR (Ceylonica) OR (Chagres) OR (Claytoni) OR (Copenhageni) OR (Copenhageni) OR (Corredores) OR (Costa Rica) OR (Coxi) OR (Cristobali) OR (Cuica) OR (Cynopteri) OR (Dadas) OR (Dakota) OR (Darien) OR (Dehong) OR (Dikkeni) OR (Djasiman) OR (Djatzi) OR (Erinaceiauriti) OR (Evansi) OR (Fluminense) OR (Fortbragg) OR (Fugis) OR (Galtoni) OR (Gatuni) OR (Gem) OR (Gengma) OR (Georgia) OR (Geyaweera) OR (Goiano) OR (Gorgas) OR (Grippotyphosa) OR (Guangdong) OR (Guaratuba) OR (Guaricura) OR (Guidae) OR (Gurungi) OR (Haemolytica) OR (Hainan) OR (Hamptoni) OR (Hardjo) OR (Hawain) OR (Hebdomadis) OR (Hekou) OR (Holland) OR (Hongchon) OR (Hualin) OR (Huallaga) OR (Huanuco) OR (Hurstbridge) OR (Icterohaemorrhagiae) OR (Istrica) OR (Jalna) OR (Javanica) OR (Jonsis) OR (Jules) OR (Kabura) OR (Kambale) OR (Kamituga) OR (Kanana) OR (Kaup) OR (Kenya) OR (Khorat) OR (Kisuba) OR (Kobbe) OR (Kremastos) OR (Kunming) OR (Kuwait) OR (Kwale) OR (Lai) OR (Lambwe) OR (Langati) OR (Lanka) OR (Lichuan) OR (Lincang) OR (Lora) OR (Losbanos) OR (Louisiana) OR (Luis) OR (Machiguenga) OR (Malaya) OR (Malaysia) OR (Mangus) OR (Manhao) OR (Manilae) OR (Mankarso) OR (Manzhuang) OR (Maru) OR (Medanensis) OR (Mengding) OR (Mengla) OR (Menglian) OR (Mengma) OR (Mengpeng) OR (Mengrun) OR (Menoni) OR (Mini) OR (Mogden) OR (Mooris) OR (Mozdok) OR (Muelleri) OR (Muenchen) OR (Mujunkumi) OR (Mwogolo) OR (Myocastoris) OR (Naam) OR (Nanla) OR (Navet) OR (Ndahambukuje) OR (Ndambari) OR (Nicaragua) OR (Nigeria) OR (Nona) OR (Nyanza) OR (Orleans) OR (Paidjan) OR (Panama) OR (Patoc) OR (Perameles) OR (Peru) OR (Peruviana) OR (Pina) OR (Pinchang) OR (Poi) OR (Polonica) OR (Pomona) OR (Portblairi) OR (Portlandvere) OR (Princestown) OR (Proechimys) OR (Pyrogenes) OR (Qingshui) OR (Rachmati) OR (Rama) OR (Ramisi) OR (Ranarum) OR (Ratnapura) OR (Recreo) OR (Ricardi) OR (Rio) OR (Rioja) OR (Robinsoni) OR (Roumanica) OR (Ruparupae) OR (Rushan) OR (Sanmartini) OR (Santarosa) OR (Saopaulo) OR (Sarmin) OR (Saxkoebing) OR (Schueffneri) OR (Sejroe) OR (Semaranga) OR (Sentot) OR (Shermani) OR (Sichuan) OR (Smithi) OR (Sofia) OR (Sokoine) OR (Sorexjalna) OR (Soteropolitana) OR (Srebarna) OR (Sulzerae) OR (Sumneri) OR (Szwajizak) OR (Tabaquite) OR (Tarassovi) OR (Tingomaria) OR (Tonkini) OR (Topaz) OR (Trinidad) OR (Tropica) OR (Tsaratsovo) OR (Tunis) OR (Valbuzzi) OR (Vanderhoedeni) OR (Varela) OR (Vargonicas) OR (Varillal) OR (Vughia) OR (Waskurin) OR (Weaveri) OR |

| | |
|---|---|
| | (Weerasinghe) OR (Whitcombi) OR (Wolffi) OR (Worsfoldi) OR (Yaan) OR (Yeonchon) OR (Yunnan) OR (Yunxian) OR (Zanoni) OR (Zhenkang) OR (Serovar) OR (Serogroup) OR (Serotype)) |
| 2 | (Animal*) OR (mammal) OR (reptile) OR (bird) OR (avian) OR (amphibian) |
| 3 | Leptospir* |
| 4 | 1 AND 2 AND 3 (**limited to scholarly articles**) |

Search in PubMed

| Search | Search Terms |
|---|---|
| 1 | *((leptospir*) AND (animal* OR mammal* OR reptil* OR bird* OR avian OR amphibian*)) AND (abramis OR agogo OR aguaruna OR alexi OR alice OR altodouro OR andaman OR anova OR arborea OR adrenal OR argentinensis OR atchafalaya OR atlanta OR australia OR autumnalis OR babudieri OR befani OR bajan OR bakeri OR balboa OR balcanica OR ballum OR bangkinang OR banna OR bataviae OR benjamini OR beye OR biggs OR Bim OR bindjei OR bikini OR bogvere OR borincana OR brasiliensis OR bratislava OR bravo OR broom OR buenos aires OR bulgaria OR butembo OR carlo OR canal zone OR canicola OR caribe OR carimagua OR carlos OR castellani OR celledoni OR zeylanica OR charges OR clayton OR copenhageni OR copenhageni OR corredores OR costa rica OR coxi OR cristobal OR ciuca OR cynopteri OR dadas OR dakota OR darien OR dehong OR dikken OR djasiman OR djauzi OR erinacei auriti OR evansi OR fluminense OR fort bragg OR fugas OR galton OR gatuni OR Gem OR gengma OR georgia OR geyaweera OR goiano OR gorgas OR grippotyphosa OR guangdong OR guaratuba OR guaricura OR guide OR gurung OR haemolytica OR hainan OR hampton OR hardjo OR hawtin OR hebdomadis OR helou OR holland OR hongchon OR hualien OR huallaga OR huanuco OR hurstbridge OR icterohaemorrhagiae OR istria OR jalan OR javanica OR jonxis OR jules OR kabara OR kamble OR kamiyoga OR kanaya OR kaup OR kenya OR khor t OR kaszuba OR kobbe OR kremastinos OR kunming OR kuwait OR kwale OR Lai OR lambwe OR langat OR lanka OR sichuan OR lincang OR lora OR los banos OR louisiana OR luis OR machiguenga OR malaya OR malaysia OR mangus OR sanhao OR manila OR mankarso OR manzhuang OR maru OR medinensis OR mending OR mengla OR mendelian OR meng a OR mengpeng OR menglun OR menoni OR mini OR ogden OR moores OR mozdok OR muelleri OR muenchen OR mujunkums OR mwogolo OR myocastor OR naam OR nanlu OR navet OR ndahambukuje OR ndambari OR nicaragua OR nigeria OR nona OR nyanza OR orleans OR paidjan OR panama OR patoc OR perameles OR peru OR peruvian OR pina OR pinching OR Poi OR polonica OR pomona OR portblairi OR portlandvere OR princeton OR proechimys OR pyrogens OR qingshui OR rahmati OR rama OR raeisi OR ranarum OR ratnapura OR recreio OR ricardi OR Rio OR rioja OR robinson OR roumania OR rupar pa OR rusjan OR sanmartin OR santarosa OR sao paulo OR sharmin OR saxkoebing OR schueffneri OR sejroe OR semarang OR sentot OR shermani OR sichuan OR smithi OR sofia OR sokoine OR sorexjalna OR sotero politano OR srebarna OR sulzer d OR sumner OR szwajizak OR tabaquismo OR tarassovi OR tingomaria OR tonkin OR topaz OR trinidad OR tropical OR tsaratsovo OR tunis OR valbuzzi OR vanderhoeden OR varela OR vargovics OR varillal OR vugia OR wakuri OR weaver OR weerasinghe OR whitcomb OR wolff OR worsfold OR yaan OR yeoncheon OR yunnan OR yunxian OR zanoni OR zhejiang OR serovar* OR serogroup* OR serotype*).* |

Search performed in Scopus

| Search | Search Terms |
|---|---|
| 1 | ( ( TITLE-ABS-KEY ( ( ( abramis ) OR ( agogo ) OR ( aguaruna ) OR ( alexi ) OR ( alice ) OR ( altodouro ) OR ( andamana ) OR ( anhoa ) OR ( arborea ) OR ( arenal ) OR ( argentinensis ) OR ( atchafalaya ) OR ( atlantae ) OR ( australis ) OR ( autumnalis ) ) ) ) OR ( TITLE-ABS-KEY ( ( babudieri ) OR ( bafani ) OR ( bajan ) OR ( bakeri ) OR ( balboa ) OR ( balcanica ) OR ( ballum ) OR ( bangkinang ) OR ( banna ) OR ( bataviae ) OR ( benjamini ) OR ( beye ) OR ( biggis ) OR ( bim ) OR ( bindjei ) OR ( birkini ) OR ( bogvere ) OR ( borincana ) OR ( brasiliensis ) ) ) OR ( TITLE-ABS-KEY ( ( bratislava ) OR ( bravo ) OR ( broomi ) OR ( buenos aires ) OR ( bulgarica ) OR ( butembo ) OR ( camlo ) OR ( canalzonae ) OR ( canicola ) OR ( caribe ) OR ( carimagua ) ) ) OR ( ( TITLE-ABS-KEY ( ( carlos ) OR ( castellonis ) OR ( celledoni ) OR ( ceylonica ) OR ( chagres ) OR ( claytoni ) OR ( copenhageni ) OR ( copenhageni ) OR ( corredores ) OR ( costa rica ) OR ( coxi ) OR ( cristobali ) OR ( cuica ) OR ( cynopteri ) ) OR TITLE-ABS-KEY ( ( dadas ) OR ( dakota ) OR ( darien ) OR ( dehong ) OR ( dikkeni ) OR ( djasiman ) OR ( djatzi ) OR ( erinaceiauriti ) OR ( evansi ) OR ( fluminense ) OR ( fortbragg ) ) OR TITLE-ABS-KEY ( ( fugis ) OR ( galtoni ) OR ( gatuni ) OR ( gem ) OR ( gengma ) OR ( georgia ) OR ( geyaweera ) OR ( goiano ) OR ( gorgas ) OR ( grippotyphosa ) OR ( guangdong ) OR ( guaratuba ) OR ( guaricura ) OR ( guidae ) ) OR TITLE-ABS-KEY ( ( gurungi ) OR ( haemolytica ) OR ( hainan ) OR ( hamptoni ) OR ( hardjo ) OR ( hawain ) OR ( hebdomadis ) OR ( hekou ) OR ( holland ) OR ( hongchon ) OR ( hualin ) OR ( huallaga ) OR ( huanuco ) OR ( hurstbridge ) ) ) ) OR ( ( TITLE-ABS-KEY ( ( icterohaemorrhagiae ) OR ( istrica ) OR ( jalna ) OR ( javanica ) OR ( jonsis ) OR ( jules ) OR ( kabura ) OR ( kambale ) OR ( kamituga ) OR ( kanana ) OR ( kaup ) OR ( kenya ) OR ( khorat ) OR ( kisuba ) OR ( kobbe ) OR ( kremastos ) OR ( kunming ) ) OR TITLE-ABS-KEY ( ( kuwait ) OR ( kwale ) OR ( lai ) OR ( lambwe ) OR ( langati ) OR ( lanka ) OR ( lichuan ) OR ( lincang ) OR ( lora ) OR ( losbanos ) OR ( louisiana ) OR ( luis ) OR ( machiguenga ) ) OR TITLE-ABS-KEY ( ( malaya ) OR ( malaysia ) OR ( mangus ) OR ( manhao ) OR ( manilae ) OR ( mankarso ) OR ( manzhuang ) OR ( maru ) OR ( medanensis ) OR ( mengding ) OR ( mengla ) OR ( menglian ) OR ( mengma ) OR ( mengpeng ) ) OR TITLE-ABS-KEY ( ( mengrun ) OR ( menoni ) OR ( mini ) OR ( mogden ) OR ( mooris ) OR ( mozdok ) OR ( muelleri ) OR ( muenchen ) OR ( mujunkumi ) OR ( mwogolo ) OR ( myocastoris ) OR ( naam ) ) ) ) OR ( ( TITLE-ABS-KEY ( ( nanla ) OR ( navet ) OR ( ndahambukuje ) OR ( ndambari ) OR ( nicaragua ) OR ( nigeria ) OR ( nona ) OR ( nyanza ) OR ( orleans ) OR ( paidjan ) OR ( panama ) OR ( patoc ) OR ( perameles ) OR ( peru ) OR ( peruviana ) OR ( pina ) ) OR TITLE-ABS-KEY ( ( pinchang ) OR ( poi ) OR ( polonica ) OR ( pomona ) OR ( portblairi ) OR ( portlandvere ) OR ( princestown ) OR ( proechimys ) OR ( pyrogenes ) OR ( qingshui ) OR ( rachmati ) OR ( rama ) ) OR TITLE-ABS-KEY ( ( ramisi ) OR ( ranarum ) OR ( ratnapura ) OR ( recreo ) OR ( ricardi ) OR ( rio ) |

| | |
|---|---|
| | OR ( rioja ) OR ( robinsoni ) OR ( roumanica ) OR ( ruparupae ) OR ( rushan ) OR ( sanmartini ) OR ( santarosa ) OR ( saopaulo ) OR ( sarmin ) ) OR TITLE-ABS-KEY ( ( saxkoebing ) OR ( schueffneri ) OR ( sejroe ) OR ( semaranga ) OR ( sentot ) OR ( shermani ) OR ( sichuan ) OR ( smithi ) OR ( sofia ) OR ( sokoine ) OR ( sorexjalna ) OR ( soteropolitana ) OR ( srebarna ) OR ( sulzerae ) OR ( sumneri ) ) ) ) OR ( ( TITLE-ABS-KEY ( ( szwajizak ) OR ( tabaquite ) OR ( tarassovi ) OR ( tingomaria ) OR ( tonkini ) OR ( topaz ) OR ( trinidad ) OR ( tropica ) OR ( tsaratsovo ) OR ( tunis ) OR ( valbuzzi ) OR ( vanderhoedeni ) ) OR TITLE-ABS-KEY ( ( varela ) OR ( vargonicas ) OR ( varillal ) OR ( vughia ) OR ( waskurin ) OR ( weaveri ) OR ( weerasinghe ) OR ( whitcombi ) OR ( wolffi ) OR ( worsfoldi ) OR ( yaan ) ) OR TITLE-ABS-KEY ( ( yeonchon ) OR ( yunnan ) OR ( yunxian ) OR ( zanoni ) OR ( zhenkang ) OR ( serovar ) OR ( serogroup ) OR ( serotype ) ) |
| 2 | ( TITLE-ABS-KEY ( animal* ) OR TITLE-ABS-KEY ( mammal* ) OR TITLE-ABS-KEY ( reptil* ) OR TITLE-ABS-KEY ( bird* ) OR TITLE-ABS-KEY ( avian ) OR TITLE-ABS-KEY ( amphibian ) ) ) |
| 3 | ( TITLE-ABS-KEY ( leptospir* ) ) |
| 4 | #1 AND #2 AND #3 |

**Search performed in Wildlife and Ecology Worldwide, 18 August 2015**

This database was not supported anymore by the University of Otago during the updated search in March 2023. Therefore, this database was not used in the update on March 2023.

| Search | Search Terms |
|---|---|
| 1 | ((Abramis) OR (Agogo) OR (Aguaruna) OR (Alexi) OR (Alice) OR (Altodouro) OR (Andamana) OR (Anhoa) OR (Arborea) OR (Arenal) OR (Argentinensis) OR (Atchafalaya) OR (Atlantae) OR (Australis) OR (Autumnalis) OR (Babudieri) OR (Bafani) OR (Bajan) OR (Bakeri) OR (Balboa) OR (Balcanica) OR (Ballum) OR (Bangkinang) OR (Banna) OR (Bataviae) OR (Benjamini) OR (Beye) OR (Biggis) OR (Bim) OR (Bindjei) OR (Birkini) OR (Bogvere) OR (Borincana) OR (Brasiliensis) OR (Bratislava) OR (Bravo) OR (Broomi) OR (Buenos Aires) OR (Bulgarica) OR (Butembo) OR (Camlo) OR (Canalzonae) OR (Canicola) OR (Caribe) OR (Carimagua) OR (Carlos) OR (Castellonis) OR (Celledoni) OR (Ceylonica) OR (Chagres) OR (Claytoni) OR (Copenhageni) OR (Copenhageni) OR (Corredores) OR (Costa Rica) OR (Coxi) OR (Cristobali) OR (Cuica) OR (Cynopteri) OR (Dadas) OR (Dakota) OR (Darien) OR (Dehong) OR (Dikkeni) OR (Djasiman) OR (Djatzi) OR (Erinaceiauriti) OR (Evansi) OR (Fluminense) OR (Fortbragg) OR (Fugis) OR (Galtoni) OR (Gatuni) OR (Gem) OR (Gengma) OR (Georgia) OR (Geyaweera) OR (Goiano) OR (Gorgas) OR (Grippotyphosa) OR (Guangdong) OR (Guaratuba) OR (Guaricura) OR (Guidae) OR (Gurungi) OR (Haemolytica) OR (Hainan) OR (Hamptoni) OR (Hardjo) OR (Hawain) OR (Hebdomadis) OR (Hekou) OR (Holland) OR (Hongchon) OR (Hualin) OR (Huallaga) OR (Huanuco) OR (Hurstbridge) OR (Icterohaemorrhagiae) OR (Istrica) OR (Jalna) OR (Javanica) OR (Jonsis) OR (Jules) OR (Kabura) OR (Kambale) OR (Kamituga) OR (Kanana) OR (Kaup) OR (Kenya) OR (Khorat) OR (Kisuba) OR (Kobbe) OR (Kremastos) OR (Kunming) OR (Kuwait) OR (Kwale) OR (Lai) OR (Lambwe) OR (Langati) OR (Lanka) OR (Lichuan) OR (Lincang) OR (Lora) OR (Losbanos) OR (Louisiana) OR (Luis) OR (Machiguenga) OR (Malaya) OR (Malaysia) OR (Mangus) OR (Manhao) OR (Manilae) OR (Mankarso) OR (Manzhuang) OR (Maru) OR (Medanensis) OR (Mengding) OR (Mengla) OR (Menglian) OR (Mengma) OR (Mengpeng) OR (Mengrun) OR (Menoni) OR (Mini) OR (Mogden) OR (Mooris) OR (Mozdok) OR (Muelleri) OR (Muenchen) OR (Mujunkumi) OR (Mwogolo) OR (Myocastoris) OR (Naam) OR (Nanla) OR (Navet) OR (Ndahambukuje) OR (Ndambari) OR (Nicaragua) OR (Nigeria) OR (Nona) OR (Nyanza) OR (Orleans) OR (Paidjan) OR (Panama) OR (Patoc) OR (Perameles) OR (Peru) OR (Peruviana) OR (Pina) OR (Pinchang) OR (Poi) OR (Polonica) OR (Pomona) OR (Portblairi) OR (Portlandvere) OR (Princestown) OR (Proechimys) OR (Pyrogenes) OR (Qingshui) OR (Rachmati) OR (Rama) OR (Ramisi) OR (Ranarum) OR (Ratnapura) OR (Recreo) OR (Ricardi) OR (Rio) OR (Rioja) OR (Robinsoni) OR (Roumanica) OR (Ruparupae) OR (Rushan) OR (Sanmartini) OR (Santarosa) OR (Saopaulo) OR (Sarmin) OR (Saxkoebing) OR (Schueffneri) OR (Sejroe) OR (Semaranga) OR (Sentot) OR (Shermani) OR (Sichuan) OR (Smithi) OR (Sofia) OR (Sokoine) OR (Sorexjalna) OR (Soteropolitana) OR (Srebarna) OR (Sulzerae) OR (Sumneri) OR |

| | |
|---|---|
| | (Szwajizak) OR (Tabaquite) OR (Tarassovi) OR (Tingomaria) OR (Tonkini) OR (Topaz) OR (Trinidad) OR (Tropica) OR (Tsaratsovo) OR (Tunis) OR (Valbuzzi) OR (Vanderhoedeni) OR (Varela) OR (Vargonicas) OR (Varillal) OR (Vughia) OR (Waskurin) OR (Weaveri) OR (Weerasinghe) OR (Whitcombi) OR (Wolffi) OR (Worsfoldi) OR (Yaan) OR (Yeonchon) OR (Yunnan) OR (Yunxian) OR (Zanoni) OR (Zhenkang) OR (Serovar) OR (Serogroup) OR (Serotype)) |
| 2 | ((Animal*) OR (mammal) OR (reptile) OR (bird) OR (avian) OR (amphibian)) |
| 3 | Leptospir* |
| 4 | 1 AND 2 AND 3 |

#### Table S2. Publication language of reports selected for full-text review during the systematic review of reports on *Leptospira* serovar isolations and detections in animal host species

| Language | Number of reports for full-text review N=1862 | | Number of reports unable to be reviewed N=172 | |
|---|---|---|---|---|
| | n | % | n | % |
| Afrikaans | 1 | 0.1 | 0 | 0.0 |
| Bulgarian | 7 | 0.4 | 4 | 2.3 |
| Chinese | 30 | 1.5 | 0 | 0.0 |
| Croatian | 9 | 0.4 | 0 | 0.0 |
| Czech | 7 | 0.4 | 7 | 4.1 |
| Dutch | 10 | 0.5 | 9 | 5.2 |
| English | 1376 | 55.7 | 36 | 20.9 |
| French | 55 | 3.0 | 3 | 1.7 |
| German | 49 | 2.5 | 3 | 1.2 |
| Hebrew | 1 | 0.1 | 1 | 0.6 |
| Hungarian | 4 | 0.2 | 3 | 1.2 |
| Italian | 17 | 0.8 | 1 | 0.6 |
| Japanese | 3 | 0.2 | 3 | 1.7 |
| Korean | 2 | 0.1 | 2 | 1.2 |
| Malay | 1 | 0.0 | 0 | 0.0 |
| Norwegian | 1 | 0.1 | 0 | 0.0 |
| Polish | 8 | 0.4 | 7 | 4.1 |
| Portuguese | 43 | 2.1 | 7 | 4.1 |
| Romanian | 13 | 0.7 | 3 | 1.7 |
| Russian | 91 | 4.7 | 18 | 9.9 |
| Serbian | 4 | 0.2 | 1 | 0.6 |
| Slovakian | 2 | 0.1 | 2 | 1.2 |
| Spanish | 74 | 3.4 | 15 | 8.7 |
| Swedish | 2 | 0.1 | 1 | 0.6 |
| Turkish | 1 | 0.1 | 0 | 0.0 |
| Vietnamese | 1 | 0.1 | 1 | 0.6 |
| Unknown | 50 | 2.3 | 45 | 23.8 |

#### Text S3. Reports identified until 9 March 2023 which met criteria for full-text review but could not be reviewed in the systematic review of reports on *Leptospira* serovar isolations and detections in animal host species


1. Imamura S, Yukawa M. Recent trends in leptospirosis in Japan and the use of Mongolian gerbil for test. Unknown journal.333-5.
2. Nasibullina FK. A study of sheep horses and camels for leptospirosis in the Betpak-Dala USSR Kazakhstan serotypes. 60- p.
3. Shapiro DM. Leptospirosis of silver fox and muskrat at the Balkhash fur farm USSR *Leptospira Pomona Leptospira Grippotyphosa Leptospira Kazachstanica* II. 61- p.
4. Walch EW, Soesilo R. Serological comparison of *Leptospira* strains isolated in Batavia and elsewhere. Geneesk Tijdschr Nederl Indie. 1927;67(1):84-98.
5. Chiodi E. *L. icterohemorrhagiae* in rats of Argentina. Rev Inst Bact Dept Nation Hig [Buenos Aires]. 1934;6(3):342-6.
6. Klarenbeek A. *Leptospirosis icterohaemorrhagiae* (Weilsche Krankheit) beim Hunde. Twelfth Internal Vet Congress N Y. 1934;3:349-57.
7. Walch-Sorgradger B, Bohlander L, Schuffner WAP. Leptospirosis in Australia and some notes on species determination of isolated strains. Geneesk Tijdschr Nederl Indie. 1938;78(38):2299-305.
8. Borg Petersen C, Christensen HI. Leptosplrosis sejroe. Ugeskrift for Laeger. 1939;101(23):697-700.
9. Collier WA, Mochtar A. A serologically deviating *Leptospira* strain from the kidney of a bat. Geneesk Tijdschr Nederl Indie. 1939;79(4):226-31.
10. Mochtar A. Occurrence of *Leptospira* in pigs in Batavia. Geneesk Tijdschr Nederland Indie. 1940;80(40):2334-45.
11. Ottosen HE. Om Leptospirainfektioner hos Rotter *Leptospira* infections in rats. Maanedsskr Dyr Laeger. 1941;53(7):173-81.
12. Bernkopf H. Experimental work on leptospirosis in cattle and men in Palestine. Harefuah [Jour Palestine Jewish Med Assoc]. 1946;30:109-10.
13. Holm H. Clinical aspects of leptospirosis. Maanedsskr Dyrlaeger. 1946;58(10):189-214.
14. Wolff JW, Bohlander H, Ruys AC. Researches on Leptospirosis ballum. The detection of urinary carriers in laboratory mice. Antonie van Leeuwenhoek. 1949;15(1):1-13.
15. Popp L. Endemic occurrence of field-fever in Hanover. Naturwiss. 1950;37(1):22-.
16. Bordjoski M. Leptospirosis sejroe in Serbia. Srpski Arhiv. 1952;80(2/3):129-37.
17. Angelov S, Kuiumdzhiev I. Animal and human leptospirosis in Bulgaria with special reference to causative species of *Leptospira*. Izvestiia na Mikrobiologicheskiia institut. 1955;6:3-10.
18. Sebek Z. Spontaneous appearance of *Leptospira sejroe* in white mice. Ceskoslovenska epidemiologie, mikrobiologie, imunologie. 1957;6(5):325-6.
19. Amosenkova N. Data from a study of additional reservoirs of *Leptospirae* in large cities. I. Infestation of dogs with icterohemorrhagic *Leptospira*. Tr Inst Epidemiol Mikrobiol I Gig Im Pastera. 1958;18:158-66.
20. Angelov S, Kuiumdzhiev I. Human and animal leptospirosis in Bulgaria and survey of causative strains of

*Leptospira*. IV. Izvestiia na Mikrobiologicheskiia institut. 1958;9:3-8.
21. Murphy LC, Cardeilhac PT, Alexander AD, Evans LB, Marchwicki RH. Prevalence of agglutinations in canine serums to serotypes other than *Leptospira canicola* and *Leptospira icterohaemorrhagiae*. Report of isolation of *Leptospira pomona* from a dog. Amer Jour Vet Res. 1958;19(70):145-51.
22. Oka T. Researches for leptospirosis of the stray dogs in Sakai, Kdchi and Fukui cities. Bull Univ Osaka Prefect Ser B Agric and Biol. 1958;8:33-7.
23. Parnas J. Results of the general investigation of the scientific expeditions on the leptospiroses for the years 1954, 1955, 1956 and 1957. Arch Inst Pasteur Tunis. 1958;35(3/4):275-300.
24. Roth EE, Knieriem BB. The natural occurrence of *Leptospira pomona* in an opossum - a preliminary report. Jour Amer Vet Med Assoc. 1958;132(3):97-8.
25. Soloshenko IZ. The part played by blood-sucking Arthropoda in the transmission and harbouring of pathogenic leptospires I. The part played by blood-sucking Arthropoda in the transmission and harbouring of the organism of Weil's disease. Zhur Mikrobiol Epidemiol I Immunobiol 1958;29(1/2):19-23.
26. Vysotskii BV, Malykh FS, Kuznetsov AP. Fur-bearing animals as additional reservoirs of pathogenic leptospires in the wild. Zh Mikrobiol Epidemiol i Immunobiol. 1958;29(8):1227-9.
27. Zwierz J, Durlakowa I, Karmanska K, Zwierzchowski JAN, Lazuga K, Korczynska A. Investigations of the fauna in the foci of the Tomashow Lubelski County epidemic of leptospirosis. Ann Univ Mariae Curie Sklodowska Sect D Med. 1958;13(38):421-37.
28. Zwierz J, Durlakowa J, Karmaska K, Zwierzchowski J, Lazuga K, Korczyska A. Research on fauna in focal spots of the leptospirosis epidemic in the Tomaszow Lubelski District. Medycyna Weterynaryjna. 1958;14(11):647-57.
29. Hou TC, Cheng LT, Yiu CC, Chung HL, Hsu PN. Epidemiological survey of leptospirosis in Yunnan. Chinese Jour People S Health. 1959;1(6):533-7.
30. Laboratory Of Sino-Soviet Friendship Hospital PFAF, People'S H. Further studies of animal hosts of leptospirosis in Kwangtung. Chinese Jour People S Health. 1959;1(8):733-4.
31. Lacerda PMG, de Freitas DC, Lacerda JPG. Notes on bovine leptospirosis Arq Inst Biol [Sao Paulo]. 1960;27:87-91.
32. Parnas J, Koslak A, Krukowska M. Investigations of *Leptospira bataviae*. Zentralbl Bakt Parasitenk Infektionskrankh U Hyg. 1960;180(3):379-86.
33. Babudieri B, Mateew D. Serologic studies on some Bulgarian strains of *Leptospira*. Rend Ist Super Sanita. 1961;24(9):614-22.
34. Pestana de Castro AF, Rosa CAS, Troise C. Cavies as reservoirs of *Leptospira* in Sao Paulo. Isolation of *Leptospira icterohaemorrhagiae* Biologico. 1961;27(9):207-9.
35. Santa Rosa CA, de Castro AFP, Troise C. Isolation of *Leptospira pomona* from swine in the state of Sao Paulo (Brazil). Arq Inst Biol [Sao Paulo]. 1962;29:165-74.
36. Asmera J. Results of research of leptospiroses in the district of Ostrava. Ceskoslov Parasitol. 1963;10:13-22.
37. Kir'Yanov EA. Abortions of leptospirosis etiology in pigs. Veterinariya. 1963;39(1):31-.
38. Burov AI, Kritskaya ZF. *Leptospira* carrier state in gray rats in Odessa. Zh Mikrobiol Epidemiol Immunobiol. 1964;41(10):131-5.
39. Byalik ZM. Serological investigations of fowl in the foci of leptospirosis Immunology, natural focality, and enteric

infections From: Ref Zh Biol, 1966, No. 4B817. (Translation)1965. 73-7 p.
40. Truszczynski M, Closek D, Tereszczuk S. Serotypes of Leptospira isolated from cases of colibaclllosis and hog cholena in Poland. Med Weter. 1965;21(10):584-9.
41. Anan'In VV, Smirin VM, Khalimov MK, Kokovin IL, Panova VV, Sakhartseva TF. Natural foci of leptospirosis in south-west Tajikistan. Zool Zh. 1966;45(7):1090-2.
42. Anokhof II. No english title available. Zh Mikrobiol Epidemiol Immunobiol. 1966;43(8):149-50.
43. Grannkova SA, Smirin VM, Vorob'Eva RN. A description of the natural leptospirosis foci in the Central Amur River region. Zh Mikrobiol Epidemiol Immunobiol. 1966;43(9):75-7.
44. Popova EM, Sil'ianova VI, Iachmenev NI. Canicola leptospirosis in the northwestern USSR. Trudy Leningradskogo nauchno-issledovatel'skogo instituta epidemiologii i mikrobiologii imeni Pastera. 1966;29:36-43.
45. Tagi-Zade TA, Borisova LP. *Leptospira* carrier state among nutrias (*Myocastor coypus*). Zh Mikrobiol Epidemiol i Immunobiol. 1966;43(8):146.
46. Cacchione RA, Bulgini MJD, Cascelli ES, Martinez ES. Leptospirosis in wild animals. Recent study on its investigations: Isolation and classification from Argentinian sources. Rev Fac C Vet 1967;20 37-54.
47. Grabinski J, Karmanska K. A case fo canine leptospirosis caused by the Ballum serotype. Medycyna Weterynaryjna. 1967;23(8):487-8.
48. Bravo C, Restrepo M, Robledo M, Perez J. Leptospirosis in Antioquia, Colombia I: isolation of *Leptospira pomona* in pigs. Antioquia Medica. 1968;18(6):475-81.
49. Kir'ianov EA. Leptospirosis in cattle caused by *Leptospira bataviae*. Veterinariia. 1968;45(4):36.
50. Aguirre WG, Silva I. Isolation in Argentina of *Leptospira pyrogenes*. Annali dell'Istituto Superiore di Sanita. 1969;5(3-4):195-6.
51. Kadlcik K, Kramar R, Polednikova I, Tondl F, Vychodil J. Occurrence of etiologic agents of some anthropozoonoses in small mammals and domestic animals. Ceskoslovenska Epidemiologie Mikrobiologie Imunologie. 1969;18(4):199-204.
52. Tagi-Zade TA, Alekperov FP. Water fowl of the Kyzylagach preserve and their possible role in the epidemiology of leptospirosis. Izvestiya Akademii Nauk Azerbaidzhanskoi SSR Seriya Biologicheskikh Nauk. 1969(4):127-31.
53. Liceras de Hidalgo J, Hidalgo R. Leptopirosis in cattle and slaughtermen of Tumbes, Peru. Boletin de la Oficina Sanitaria Panamericana Pan American Sanitary Bureau. 1970;68(4):297-306.
54. Rosa CAS, Campedelli Filho O, Castro AFP. Isolation of the leptospiral serotypes Pomona and Guidae from apparently normal pigs1970. 413- p.
55. Rosa CAS, Silva ASD, Giorgi W. Abortion in pigs isolation of the leptospiral serotype Pomona and Brucella suis1970. 412- p.
56. Teodorovici G, Mardari A, Ivan A, Nastase A, Buzdugan I, Oana C, et al. Contributions to the study of leptospiroses on the territory of Moldavia in the 1963-1969 period. Institutul Agronomic "Ion Ionescu De La Brad" Iasi Lucrari Stiintifice II Zootehnie-Medicina Veterinara. 1970:379-85.
57. Arzumanian G, Espino R, Rodriguez S, Ramirez W, Lorenzo J, Monet VM. Some studies of leptospirosis in dogs in Havana. Academia de Ciencias de Cuba Serie Biologica. 1971(36):1-8.
58. Carlos ER, Kundin WD, Watten RH, Tsai CC, Irving GS, Carlos ET, et al. Leptospirosis in teh Philippines feline

studies. Am J Vet Res. 1971;32(9):1455-6.

77. Ellis WA, O'Brien JJ, Neill S, Hanna J, Bryson DG. The isolation of a leptospire from an aborted bovine fetus. Vet

Rec. 1976;99(23):458-9.
78. Ellis WA, O'Brien JJ, Pearson JK, Collins DS. Bovine leptospirosis: infection by the Hebdomadis serogroup and mastitis. Vet Rec. 1976;99(19):368-70.
79. Hykutake S, de Biasi P, Santa Rosa CA, Belluomni HE. Epidemiological study on leptospirosis in Brazilian snakes. Revista do Instituto de Medicina Tropical de Sao Paulo. 1976;18(1):10-6.
80. Kitaoka M, Otsuka S. *Leptospira canicola* isolated from *Apodemus speciosus speciosus* in Kyushu Japan. Jpn J Med Sci Biol. 1976;29(1):45-7.
81. Manev K, Yanakieva M. Typing of Bulgarian strains of *Leptospira pomona* isolated from man, domestic animals and rodents, using specific sera. Epidemiologiya, Mikrobiologiya i Infektsiozni Bolesti. 1976;13(2):164-7.
82. Carpio M, Wobeser G, Iversen J. *Leptospira interrogans* serotype Pomona in Saskatchewan: isolation from a naturally infected striped skunk. Canadian Journal of Microbiology. 1977;23(12):1654-6.
83. Crowell WA, Stuart BP, Adams WV. Renal lesions in striped skunks (*Mephitis mephitis*) from Louisiana. J Wildl Dis. 1977;13(3):300-3.
84. Kmety E, Bakoss P, Manicova E, Stupalova S, Hrabinsky M, Peci J. First observations of leptospirosis in Slovakia. Bratislavske Lekarske Listy. 1977;68(1):37-41.
85. Straton C, Bercovici C, Straton A, Beldiman N, Stefanescu E, Anton N, et al. Study of the natural reservoir of *Leptospira* in the Danube delta. I. Chilia branch region. Revista medico-chirurgicala a Societatii de Medici si Naturalisti din Iasi. 1977;81(2):233-7.
86. Vosta J, Hanak P, Vychodil J. Effect of population dynamics of small mammals on spread and survival of *Leptospira grippotyphosa* in foci. Bratislavske Lekarske Listy. 1977;68(1):50-6.
87. Corrigall W. Naturally occurring leptospirosis (*Leptospira ballum*) in a red deer (*Cervus elaphus*). Vet Rec. 1978;103(4):75-6.
88. Straton A, Straton C, Bercovici C, Beldiman N, Stefanescu E, Anton N, et al. Study of the natural reservoir of *Leptospira* in the Danube Delta. III. Region of the Sfintu Gheorghe branch. Revista medico-chirurgicala a Societatii de Medici si Naturalisti din Iasi. 1978;82(1):69-72.
89. Sebek Z, Hodkova Z, Palicka P, Zitek K, Valova M. First demonstration of natural foci of L*eptospira pomona* in the Czech Socialist Republic. Ceskoslovenska epidemiologie, mikrobiologie, imunologie. 1979;28(3):155-62.
90. Cacchione RA, Cascelli ES, Saravi MA, Martinez ES. Distribution and importance of leptospirosis among animals and human beings in Argentina. Revista de Medicina Veterinaria, Argentina. 1980;61(3):236-42, 44-47.
91. Espino R, Cabrera L, Cornide RI. Isolation of *Leptospira pomona* from *Mus musculus* obtained from a rice growing zone of Havana province, Cuba. Academia de Ciencias de Cuba Informe Cientifico-Tecnico. 1981(164):1-7.
92. Liceras de Hidalgo J. Leptospirosis in Tingo María, Huánuco Department, Peru. II. Study in wild animals. Bull Pan Am Health Organ. 1981;91(1):47-55.
93. Liceras de Hidalgo J, Hidalgo R, Flores M. Leptospirosis in Tingo María, Department of Huánuco, Peru. I. Study on man and domestic animals. Bull Pan Am Health Organ. 1981;90(5):430-8.
94. Saltaren Cobas A, Cornide Gonzalez RI. Leptospirosis in swine of the city and provinces of Havana, Cuba. Academia de Ciencias de Cuba Informe Cientifico-Tecnico. 1981(170):1-17.
95. Zoonotic disease surveillance. Weekly Epidemiological Record. 1982;57(42):321-4.

114. Cinco M, Panfili E, Banfi E, Schonberg A, Everard COR. Remarks on *Leptospira* identification at species level additional tests to better define their saprophytic or parasitic nature. Israel J Vet Med. 1988;44(1):31-6.
115. Kocik T. Actual problems of leptospirosis in animals in Poland. Przeglad epidemiologiczny. 1988;42(4):364-9.
116. Kondratenko VF, Bunin IK, Rodionova NS, Danilkin AP, Yagovkin EA. Wildlife focus of *Leptospira icterohaemorrhagiae* in the Lower Don region of the USSR. Zh Mikrobiol Epidemiol Immunobiol. 1988(5):38-41.
117. Mani R. Bovine leptospirosis: a case report. Indian Vet J. 1988;65(10):924-.
118. Desmecht M, Smeets L. Ondatra one of the main sources of leptospirosis of cattle. Annales de Medecine Veterinaire. 1989;133(5):413-9.
119. Espino R, Malajov Yu A, Cornide RI, Suplico AN. Taxonomic position of Leptospira strains isolated from cattle, swine and synanthropic rodents from the Republic of Cuba. Revista Cubana de Ciencias Veterinarias. 1989;20(1):89-94.
120. Gonzalez Gallo JA, Jimenez R, Martinez A. Leptospirosis in pigs in the Villa Clara Province, Cuba. Revista Cubana de Ciencias Veterinarias. 1989;20(4):251-6.
121. Jarekova J, Kmety E. Following the evolution of the natural foci of leptospirosis in chosen localities of Zahorska lowlands. Entomologicke Problemy. 1989;19:347-55.
122. Kocik T. Isolation of *Leptospira interrogans* serovar Mozdok from an outbreak of leptospirosis in pigs. Medycyna Weterynaryjna. 1989;45(7):409-11.
123. Nagy G. Isolation of leptospires in a semi-solid medium. Magyar Allatorvosok Lapja. 1989;44(6):349-52.
124. Agaev IA. The self-maintenance of natural foci of leptospirosis. Zh Mikrobiol Epidemiol i Immunobiol. 1990(12):40-4.
125. Batra HV, Chandiramani NK, Mandokhot UV. Prevalence of leptospirosis in farm animals in Haryana, India. Indian J Anim Sci. 1990;60(7):755-60.
126. Bolin CA, Cassells JA. Isolation of *Leptospira interrogans* serovar Bratislava from stillborn and weak pigs in Iowa. J Am Vet Med Assoc. 1990;196(10):1601-4.
127. Makeev SM, Maramovich AS, Iaroshenko VA. Immunologic monitoring of leptospirosis in the Maritime Territory. Zh Mikrobiol Epidemiol i Immunobiol. 1990(9):31-6.
128. Makeev SM, Maramovich AS, Yaroshenko VA. Immunological monitoring of Leptospira infections in Primorski Krai Russian SFSR USSR. Zh Mikrobiol Epidemiol i Immunobiol. 1990(9):31-6.
129. Murray RD. A field investigation of causes of abortion in dairy cattle. Vet Rec. 1990;127(22):543-7.
130. Nagy G. Isolation of Leptospira interrogans serovar Hardjo from lactating cows. Magyar Allatorvosok Lapja. 1990;45(8):485-8.
131. Rocha T. Isolation of *Leptospira interrogans* serovar Mozdok from aborted swine fetuses in Portugal. Vet Rec. 1990;126(24):602.
132. Desmecht M, Korver H, Terpstra WJ. Isolation of *Leptospira interrogans* serovars Saxkoebing, Grippotyphosa and Copenhageni from muskrats in Belgium. Vlaams Diergeneeskundig Tijdschrift. 1991;60(2):59-63.
133. Jung SC, Kim JM, Park JM, Choi WP. Biochemical characterization of *Leptospira interrogans* isolated from animals in Korea. Research Reports of the Rural Development Administration (Suweon). 1991;33(3 VET):1-10.
134. Vosta J, Suchy P. Hedgehog western variety *Erinaceus europaeus* the reservoir of *Leptospira bratislava* in South

Bohemia. Sbornik Agronomicka Fakulta v Ceskych Budejovicich Zootechnicka Rada. 1991;8(1):3-12.

153. Suepaul S, Borde G, Carrington C, Campbell M, Chadee D, Adesiyun AA. Serovars of *Leptospira* isolates recovered from suspected cases of canine leptospirosis, apparently healthy stray dogs and rodents in Trinidad. Bangalore: Commonwealth Veterinary Association; 2007. p. 100.
154. Langoni H, Souza LCd, Silva AVd, Cunha ELP, Silva RCd. Epidemiological aspects in leptospirosis. Research of anti-*Leptospira* spp. antibodies, isolation and biomolecular research in bovines, rodents and workers in rural properties from Botucatu, SP, Brazil. Braz J Vet Res An Sci. 2008;45(3):190-9.
155. Clark DV, Kuchuloria T, Akhvlediani T, Hepburn MJ, Pimentel G, Chokheli M, et al. Leptospirosis in the Republic of Georgia. Am J Trop Med Hyg. 2009;81(5, Suppl. S):223-4.
156. Proceedings of the IV Symposium of the Brazilian Association of Equine veterinarians - Abraveq - Nordeste, Praia de Portos de Galinhas, Ipojuca, Pernambuco, Brazil, 24-26 September 20102010; Zumbi: CRMV-PE.
157. Isambert M, Bellina A. The coypu (*Myocastor coypus*): pest to man - species monitoring. Bourgogne Nature. 2011;14:177-82.
158. Rodrigues RO, Silva JA, Alves TM, Dorneles EMS, Minharro S, Lage AP, et al. Characterization of outer membrane proteins of serovar Hardjo isolated from cattle in Minas Gerais, Brazil. Pesquisa Veterinaria Brasileira. 2011;31(7):555-60.
159. Yoshiki A, Matoba Y, Asakawa M, Takahashi T, Nakano Y, Kikuchi N. Isolation of Leptospira from raccoons and serological survey of leptospirosis in Hokkaido, Japan. J Vet Epidemiol. 2011;15(2):100-5.
160. First National Veterinary Epidemiology Meeting, Brazil, 20122012; Porto Alegre: Universidade Federal do Rio Grande do Sul, Faculdade de Veterinária.
161. Panin AN, Viktorova EV. Animal leptospirosis in the Russian Federation. 2012:22-3.
162. Ponti MN, Canu M, Carboni GA, Noworol M, Palmas B, Picardeau M, et al. Canine leptospirosis: report of an outbreak. 2012:86-7.
163. New serovars of *Leptospira* isolated in INCIENSA. Boletín INCIENSA. 2013;25(1):11.
164. Maletskaia OV, Beliaeva AI, Taran TV, Agapitov DS, Kulichenko AN. Epidemiologic situation on dangerous infectious diseases on the territory of Republic of Abkhazia. Zh Mikrobiol Epidemiol i Immunobiol. 2013(5):43-7.
165. Douadi B, Nursheena MS, Lin TK. Antimicrobial Susceptibiliy of *Leptospira* spp. Isolated from Environmental, Human and Animal Sources in Malaysia. Abstr Gen Meet Am Soc Microbiol. 2014;114:19-.

Figure S4. Global frequency of number of reports per country identified in the systematic review of reports on *Leptospira* serovar isolations and detections in animal host species published 1927-2022

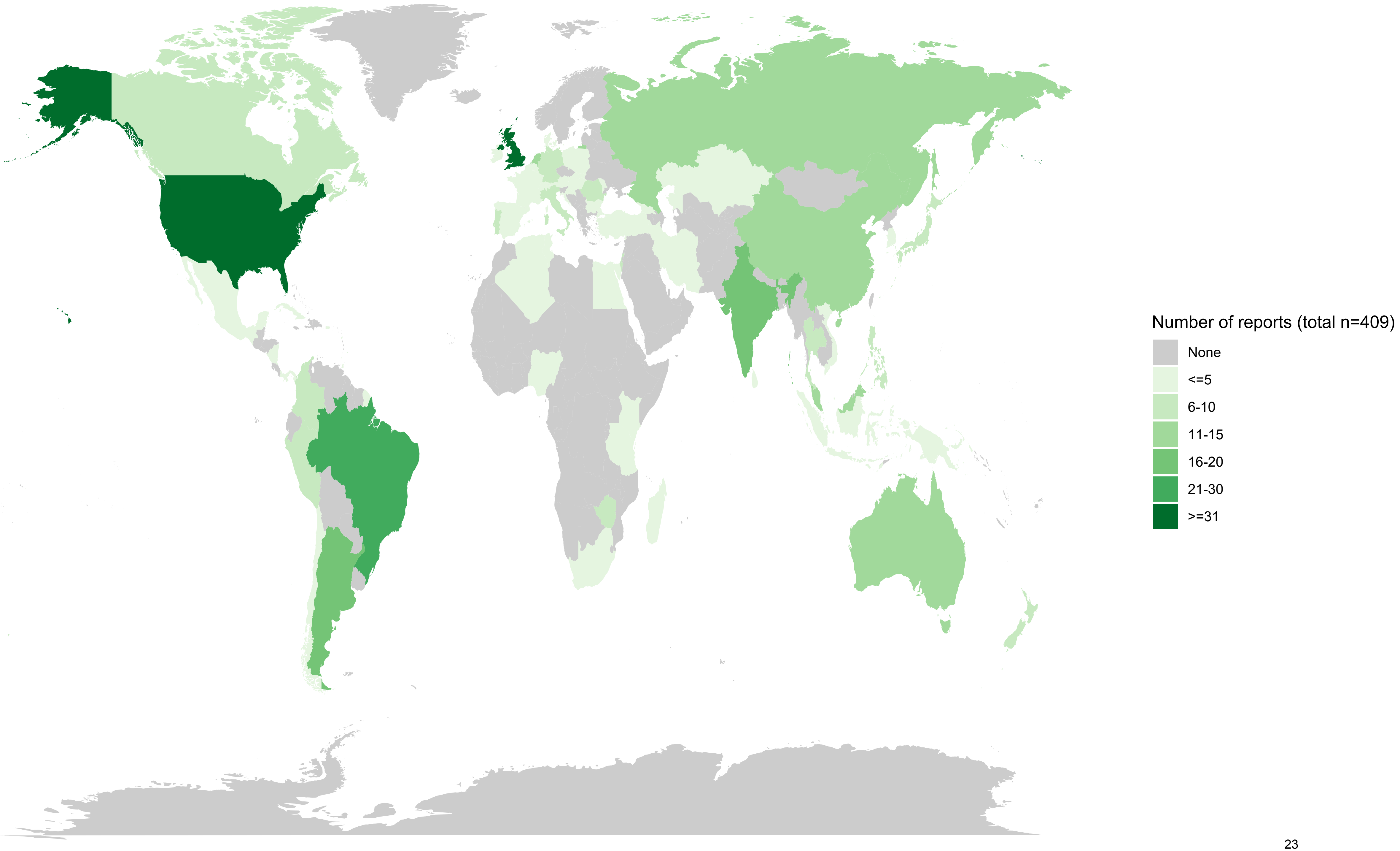
Number of reports (total n=409)
None
<=5
6-10
11-15
16-20
21-30
>=31

Figure S5. Histogram of publication years of reports in the systematic review of reports on Leptospira serovar isolations and detections in animal host species published 1927-2022

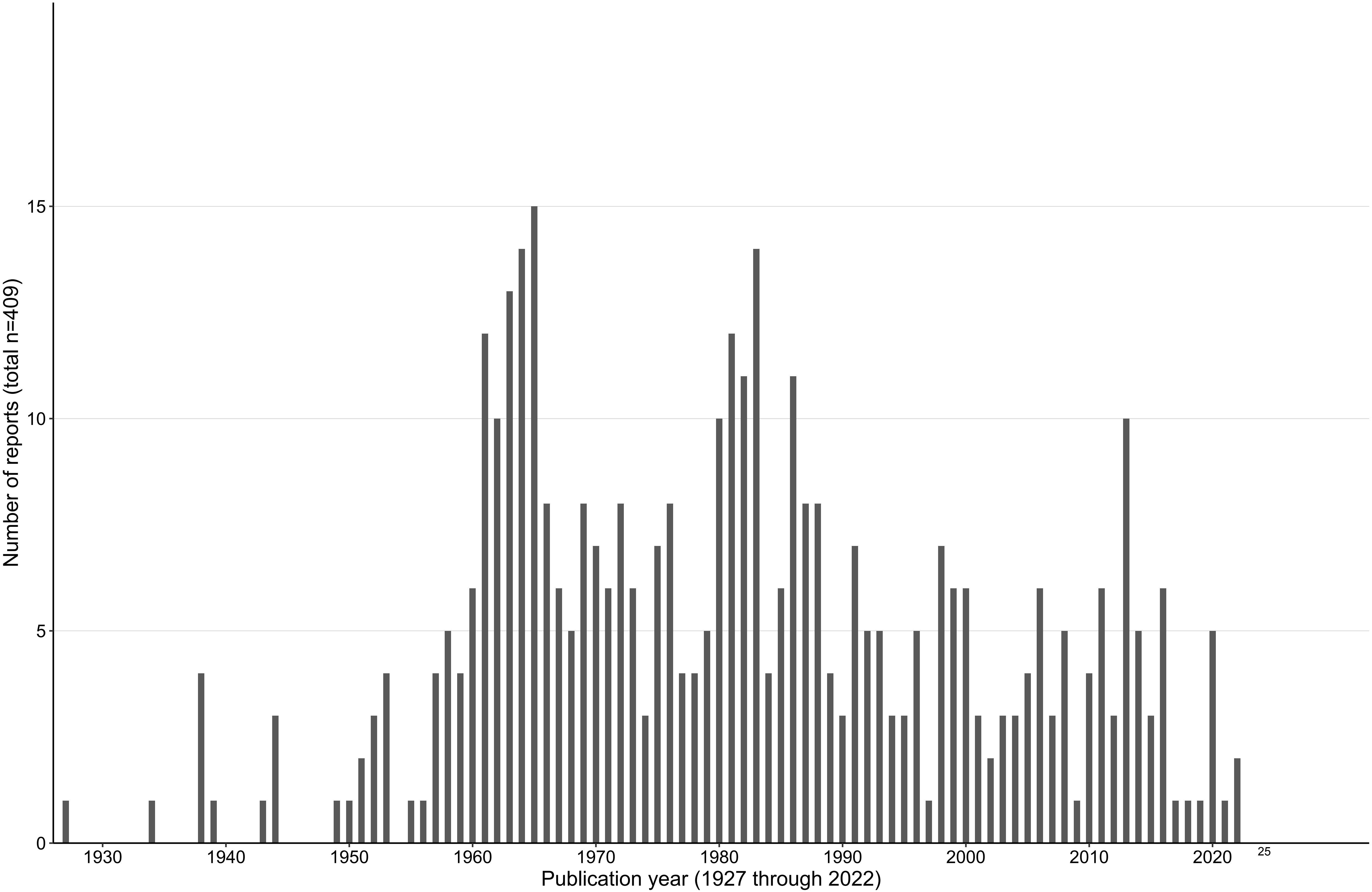

Number of reports (total n=409)
0
5
10
15
1930
1940
1950
1960
1970
1980
1990
2000
2010
2020
Publication year (1927 through 2022)

#### Table S6. Characteristics of reports included in the systematic review of reports on Leptospira serovar isolations and detections in animal host species published 1927-2022

| | Number of reports, n=409 | |
|---|---|---|
| | n | (%) |
| Publication year | | |
| 1927 through 1950 | 13 | (3.2) |
| 1951 through 1957 | 158 | (38.6) |
| 1976 through 2000 | 160 | (39.1) |
| 2001 through 2022 | 78 | (19.1) |
| Period of median year of data collection | | |
| 1922 through 1940 | 6 | (1.4) |
| 1941 through 1960 | 49 | (11.8) |
| 1961 through 1980 | 121 | (29) |
| 1981 through 2000 | 69 | (16.5) |
| 2001 through 2021 | 47 | (11.3) |
| Not stated | 125 | (30) |
| Method of *Leptospira* detection | | |
| Culture | 402 | (98.2) |
| Culture and PCR | 7 | (1.7) |
| Methods of *Leptospira* serovar identification* | | |
| Cross agglutination absorption test | 265 | (64.8) |
| Reference laboratory only | 66 | (16.1) |
| Monoclonal antibodies | 61 | (14.9) |
| Pulsed field gel electrophoresis | 42 | (10.3) |
| Serum factor | 14 | (3.4) |
| Number of appropriate methods of *Leptospira* serovar identification | | |
| 1 | 309 | (75.6) |
| 2 | 29 | (7.1) |
| 3 | 5 | (1.2) |
| Reference laboratory (no listed methods) | 66 | (16.1) |
| Specimen type* | | |
| Blood | 34 | (8.3) |
| Urine | 111 | (27.1) |
| Kidney | 262 | (64.1) |
| Other | 55 | (13.4) |
| Not stated | 61 | (14.9) |
| Number of reported *Leptospira* serovars, median (IQR) | 1 (1-2) | |
| Number of reported animal hosts, median (IQR) | 4 (1-13) | |
| Animal age class* | | |
| Adult | 100 | (24.4) |

| | | |
|---|---|---|
| Juvenile | 48 | (11.7) |
| Foetus/still birth | 18 | (4.4) |
| Not stated | 293 | (71.6) |
| Animal health* | | |
| Healthy/asymptomatic | 47 | (11.5) |
| Diseased | 72 | (17.6) |
| Died due to Leptospirosis | 17 | (4.2) |
| fetes aborted/still birth | 23 | (5.6) |
| Not stated | 295 | (72.1) |

*Multiple categories possible per report

#### Table S7. Number of reports of *Leptospira* serovar detected per animal host order in the systematic review of reports on *Leptospira* serovar isolations and detections in animal host species published 1927-2022

| Animal host class | Animal host order, examples of animal | Number of reports (proportion of total, n=409)* | | Number of unique animal hosts defined by genus and species (proportion of all animal hosts, n=144)^ | |
|---|---|---|---|---|---|
| | | N | (%) | N | (%) |
| Mammalia | Rodentia, e.g. rats, mice | 194 | (45.7) | 82 | (56.9) |
| Mammalia | Artiodactyla, e.g. cattle, pig | 159 | (36.2) | 6 | (4.2) |
| Mammalia | Carnivora, e.g. dog, skunk | 93 | (20.8) | 19 | (13.2) |
| Mammalia | Didelphimorphia, e.g. opossum | 24 | (5.9) | 9 | (6.3) |
| Mammalia | Eulipotyphla, e.g. shrew, hedgehog | 20 | (4.6) | 8 | (5.6) |
| Mammalia | Perissodactyla, e.g. horse | 14 | (3.2) | 1 | (0.7) |
| Amphibia | Anura, e.g. frog | 9 | (2) | 5 | (3.5) |
| Mammalia | Diprotodontia, e.g. brushtailed possum | 5 | (1.2) | 1 | (0.7) |
| Mammalia | Cingulata, e.g. armadillo | 4 | (1) | 2 | (1.4) |
| Mammalia | Peramelemorphia, e.g. bandicoot | 4 | (1) | 3 | (2.1) |
| Mammalia | Primates, e.g. monkey | 2 | (0.5) | 2 | (1.4) |
| Reptilia | Squamata, e.g. snake | 2 | (0.5) | 2 | (1.4) |
| Mammalia | Chiroptera, e.g. bat | 1 | (0.2) | - # | - # |
| Arachnida | Ixodidae, e.g. tick | 1 | (0.2) | 1 | (0.7) |
| Mammalia | Lagomorpha, e.g. rabbit | 1 | (0.2) | 2 | (1.4) |
| Reptilia | Testudines, e.g. turtle | 1 | (0.2) | 1 | (0.7) |

*Some reports included multiple animal host orders;
^Six animals which were only identified to animal host order, and could not be classified to animal genus/species were excluded;
### Chiroptera had one animal host which was only identified to the animal order (Fruit bat)

Figure S8. Heat map of detected *Leptospira* serovar in 16 animal host orders per animal host genus in the systematic review of reports on *Leptospira* serovar isolations and detections in animal host species published 1927-2022 (409 reports) (A, Rodentia; B, Artiodactyla; C, Carnivora; D, Didelphimorphia; E, Eulipotyphla; F, Perissodactyla; G, Anura; H, Peramelemorphia; I, Cingulata; J, Squamata; K, Chiroptera; L, 5 animal orders with 1 serovar (Diprotodontia, Ixodidae, Lagomorpha, Primates, Testudines)

**Leptospira* serovars which were not detected in the specific animal order are not shown

A, Rodentia, 90 serovars

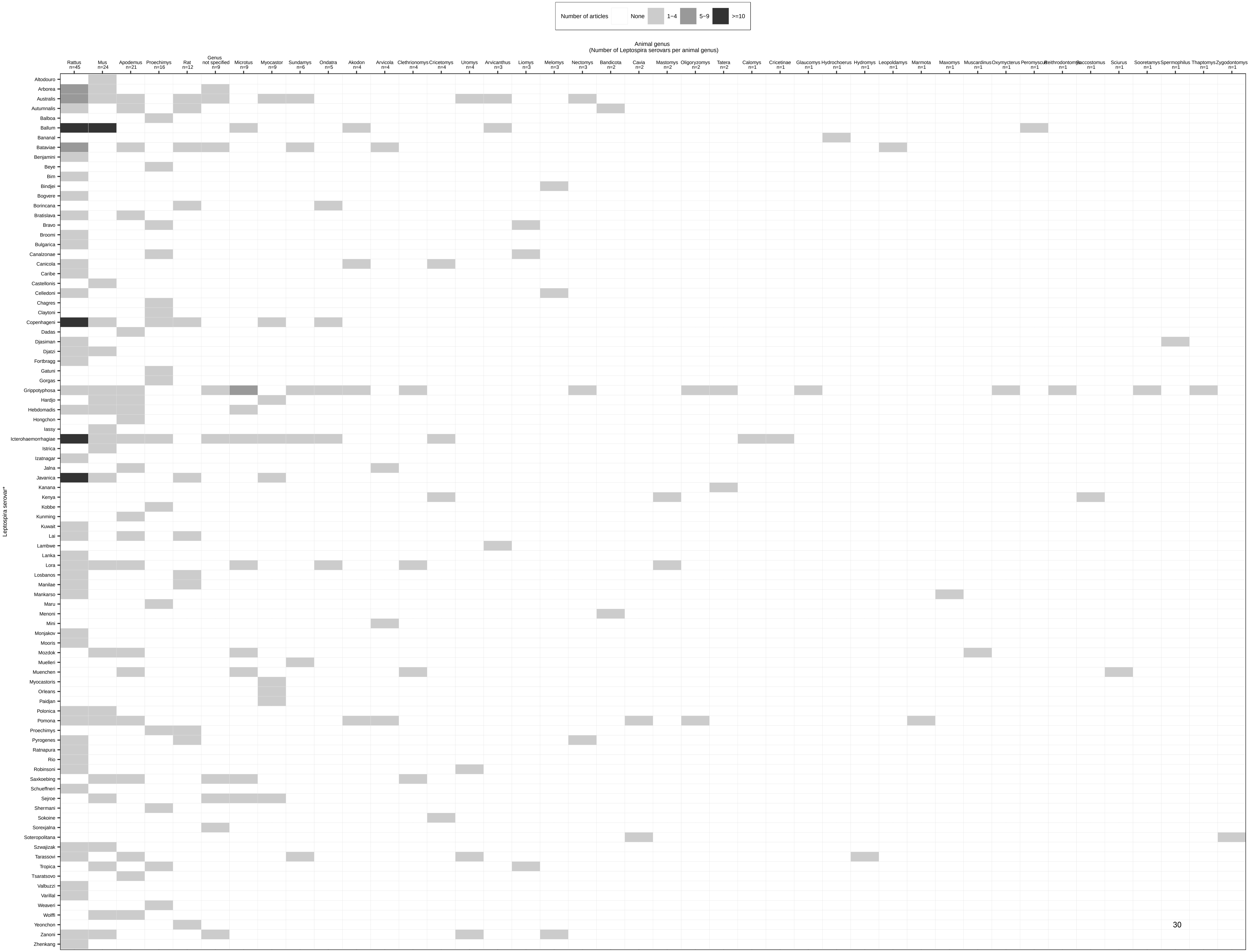

B, Artiodactyla, 58 serovars

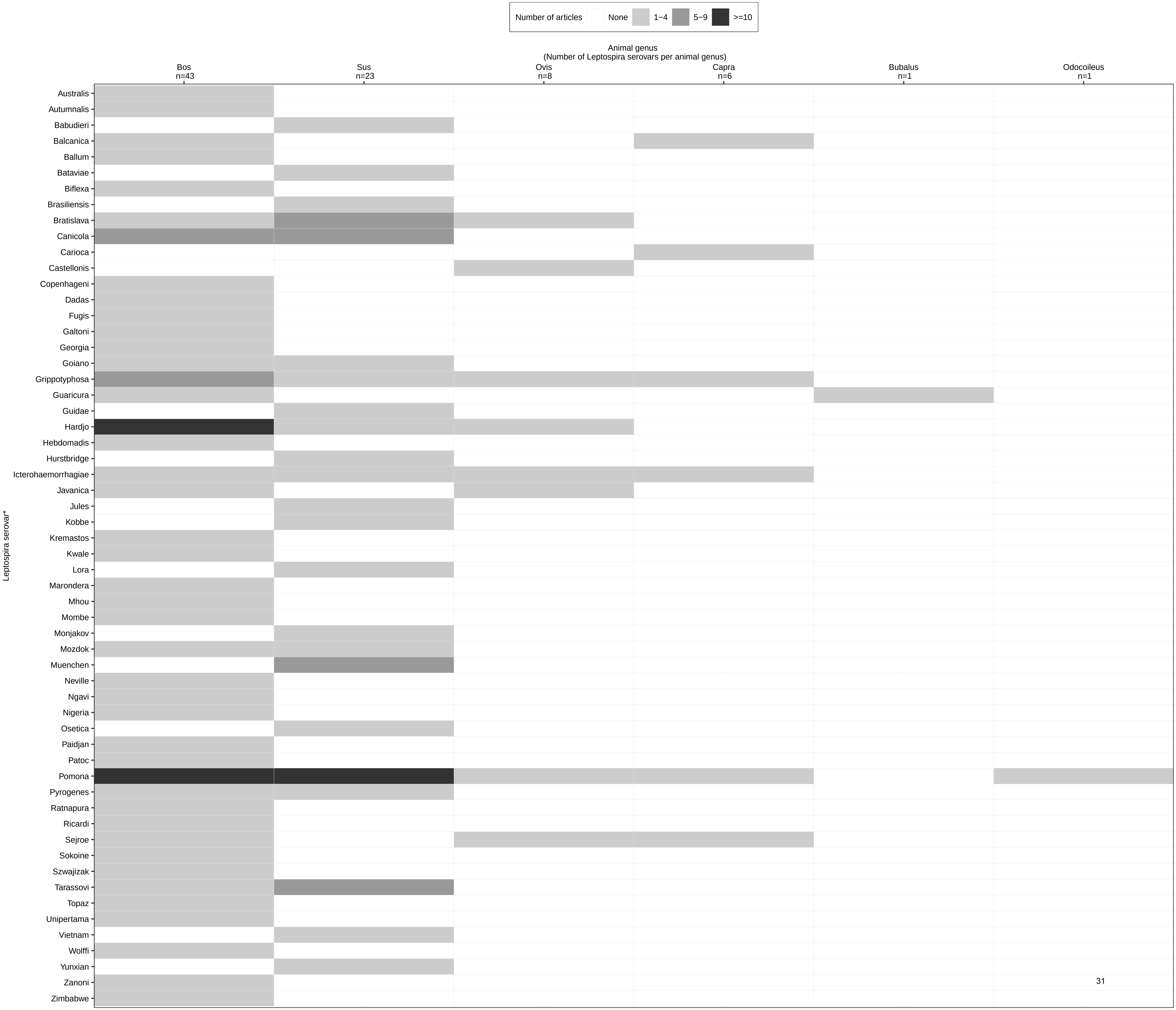

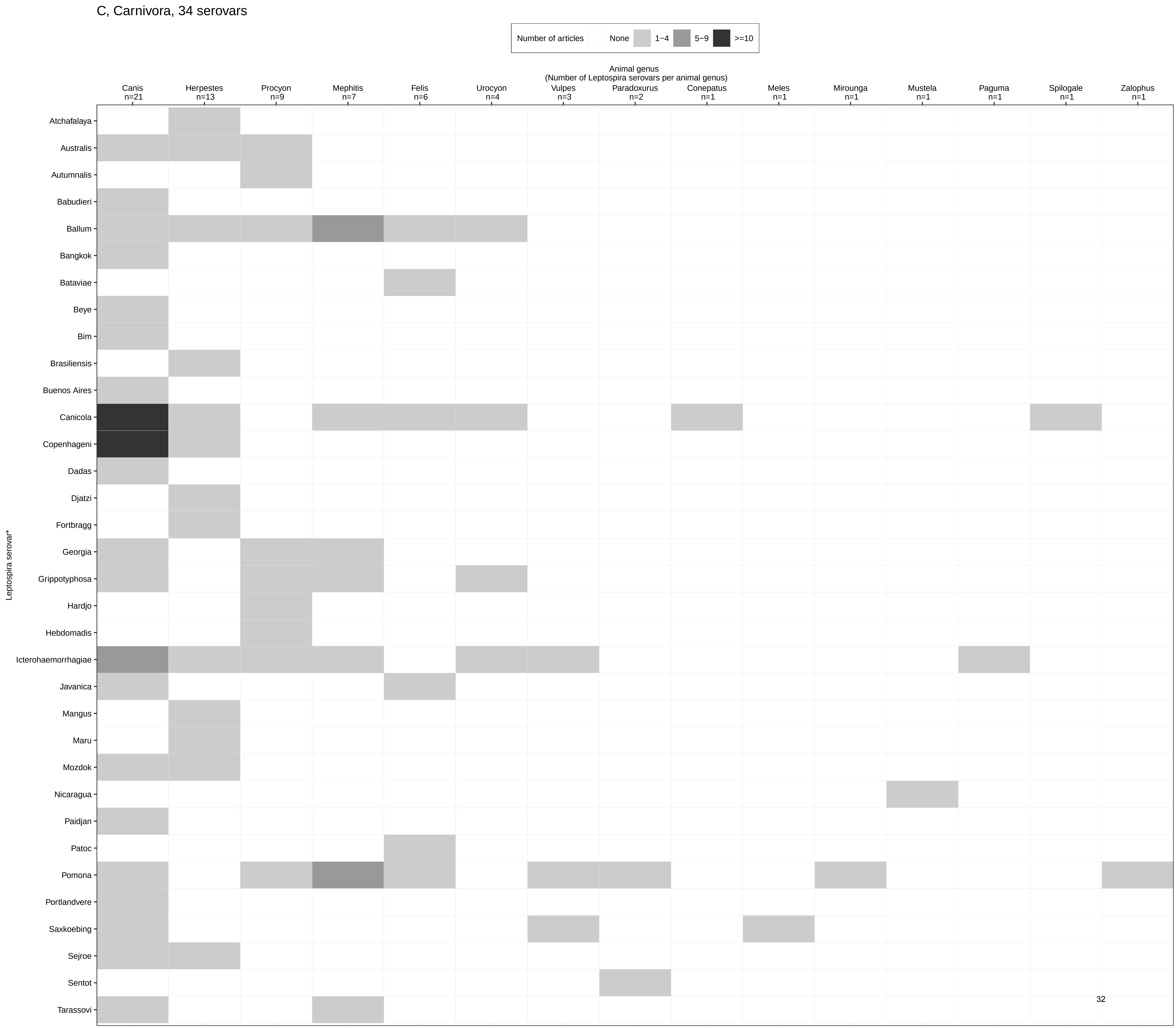

C, Carnivora, 34 serovars
Number of articles
None
1−4
5−9
>=10
Animal genus
(Number of Leptospira serovars per animal genus)
Canis n=21
Herpestes n=13
Procyon n=9
Mephitis n=7
Felis n=6
Urocyon n=4
Vulpes n=3
Paradoxurus n=2
Conepatus n=1
Meles n=1
Mirounga n=1
Mustela n=1
Paguma n=1
Spilogale n=1
Zalophus n=1
Leptospira serovar*
Atchafalaya
Australis
Autumnalis
Babudieri
Ballum
Bangkok
Bataviae
Beye
Bim
Brasiliensis
Buenos Aires
Canicola
Copenhageni
Dadas
Djatzi
Fortbragg
Georgia
Grippotyphosa
Hardjo
Hebdomadis
Icterohaemorrhagiae
Javanica
Mangus
Maru
Mozdok
Nicaragua
Paidjan
Patoc
Pomona
Portlandvere
Saxkoebing
Sejroe
Sentot
Tarassovi

D, Didelphimorphia, 31 serovars

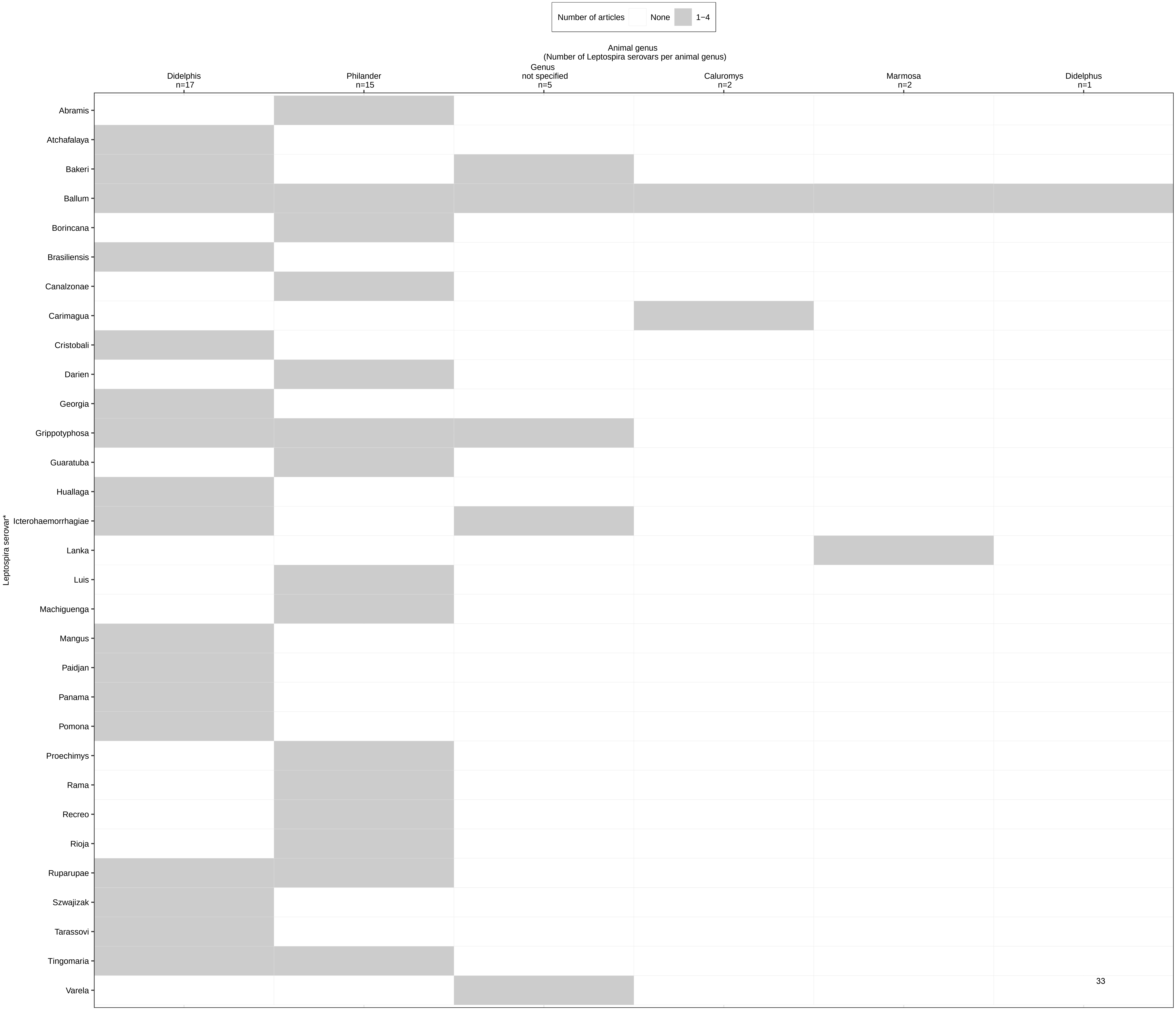

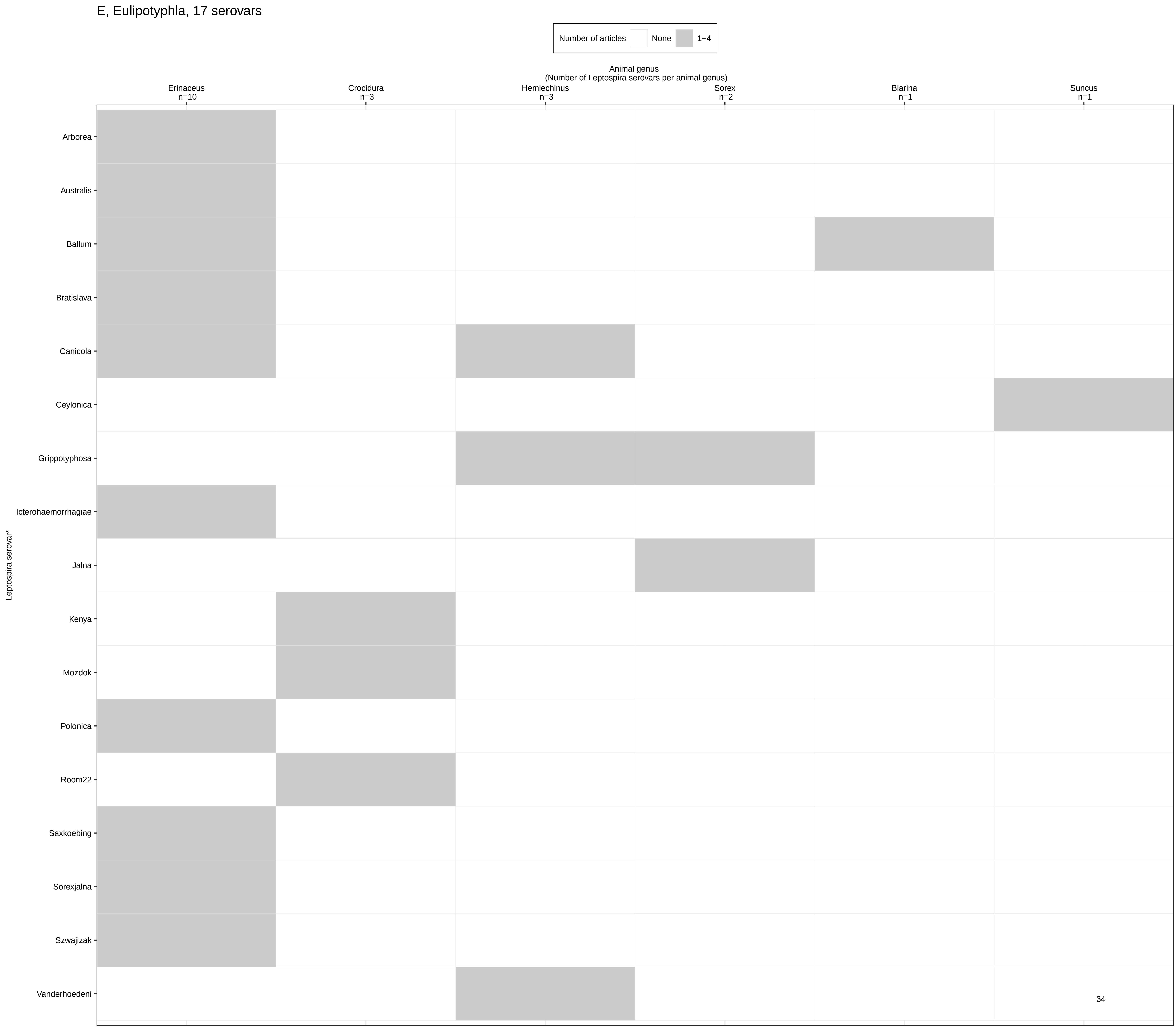
E, Eulipotyphla, 17 serovars
Number of articles
None
1−4
Animal genus
(Number of Leptospira serovars per animal genus)
Erinaceus n=10
Crocidura n=3
Hemiechinus n=3
Sorex n=2
Blarina n=1
Suncus n=1
Leptospira serovar*
Arborea
Australis
Ballum
Bratislava
Canicola
Ceylonica
Grippotyphosa
Icterohaemorrhagiae
Jalna
Kenya
Mozdok
Polonica
Room22
Saxkoebing
Sorexjalna
Szwajizak
Vanderhoedeni

F, Perissodactyla, 9 serovars

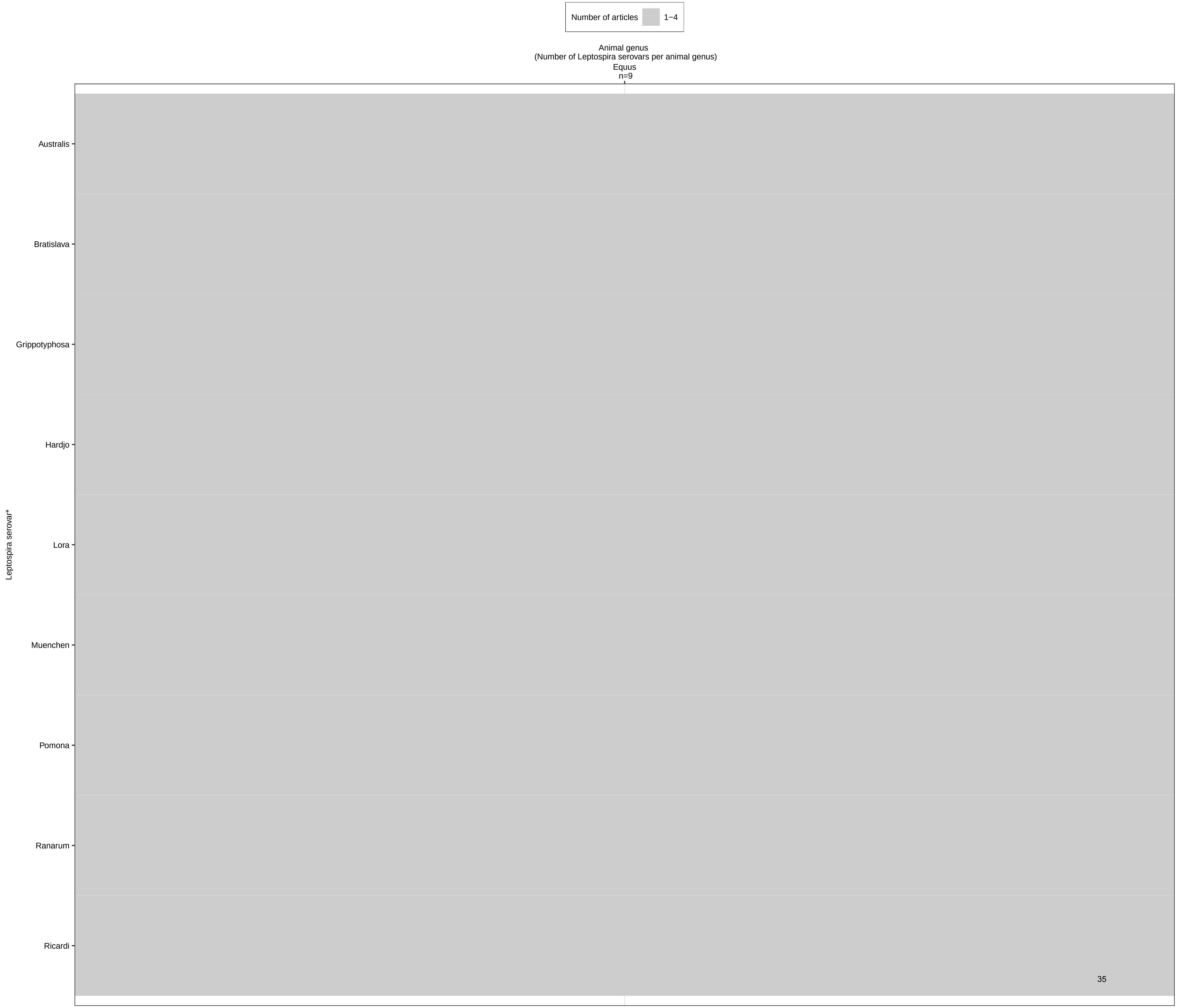

G, Anura, 8 serovars

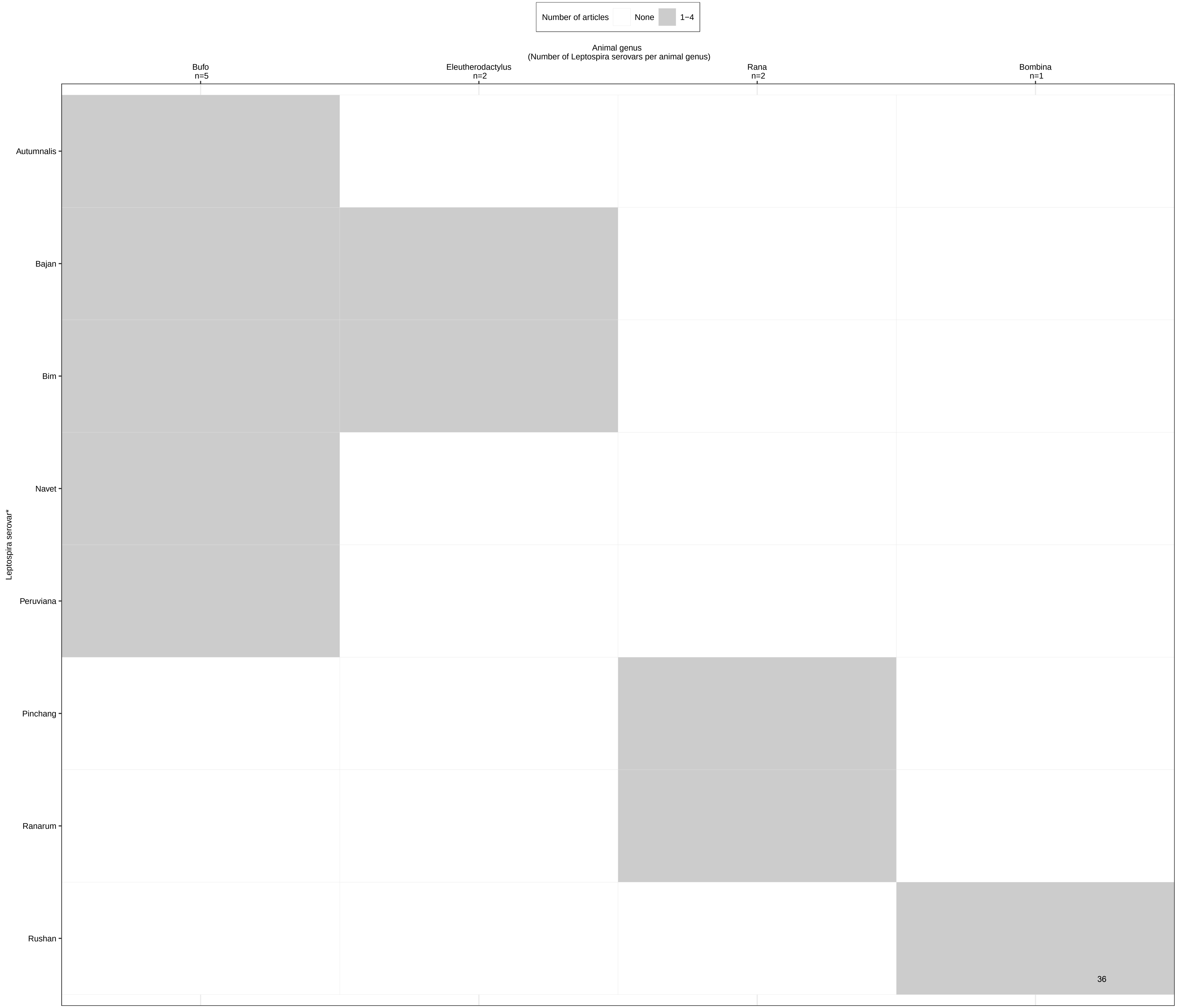

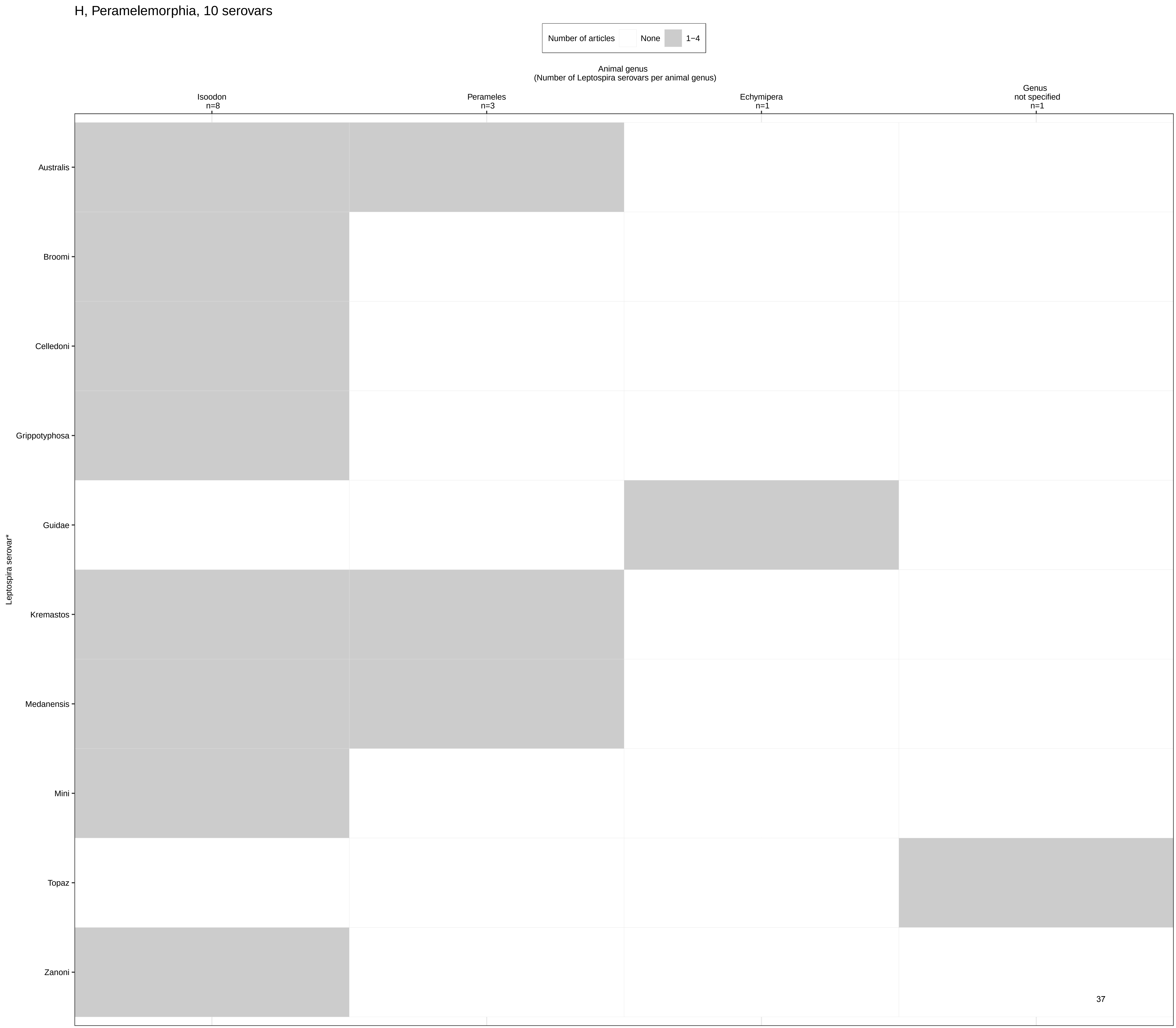
H, Peramelemorphia, 10 serovars
Number of articles
None
1–4
Animal genus
(Number of Leptospira serovars per animal genus)
Isoodon
n=8
Perameles
n=3
Echymipera
n=1
Genus
not specified
n=1
Leptospira serovar*
Australis
Broomi
Celledoni
Grippotyphosa
Guidae
Kremastos
Medanensis
Mini
Topaz
Zanoni

I, Cingulata, 6 serovars

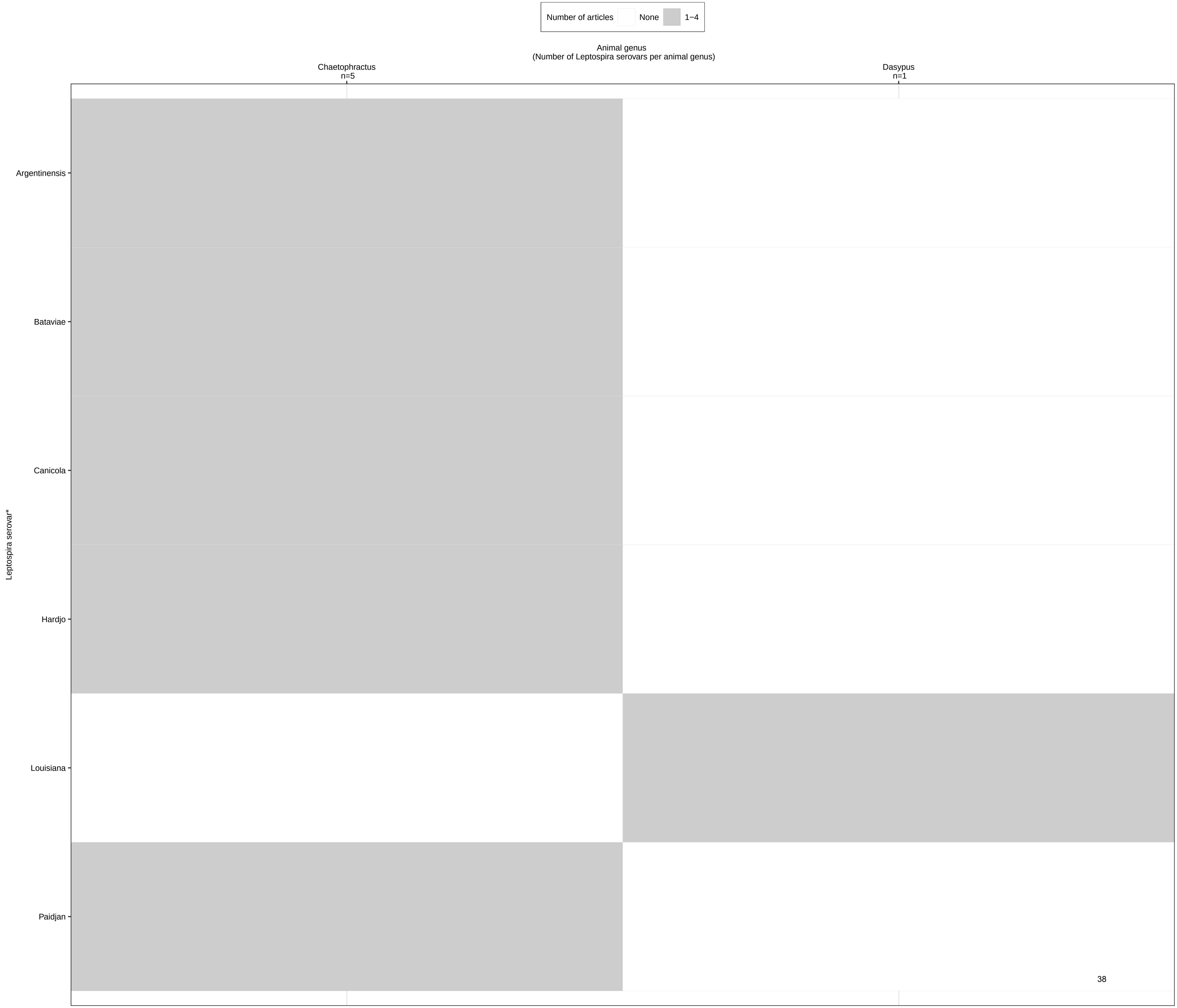

J, Squamata, 2 serovars

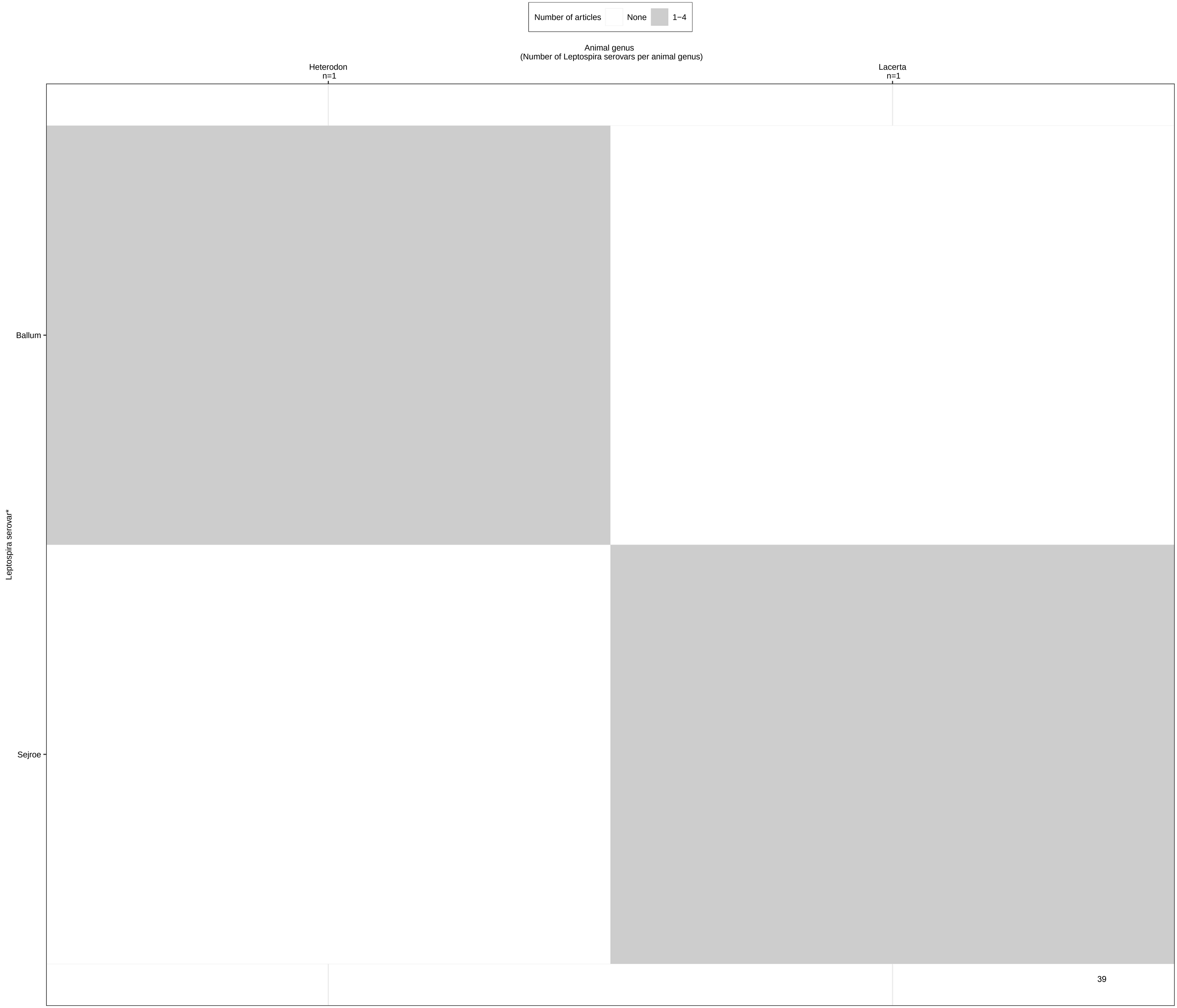

K, Chiroptera, 2 serovars

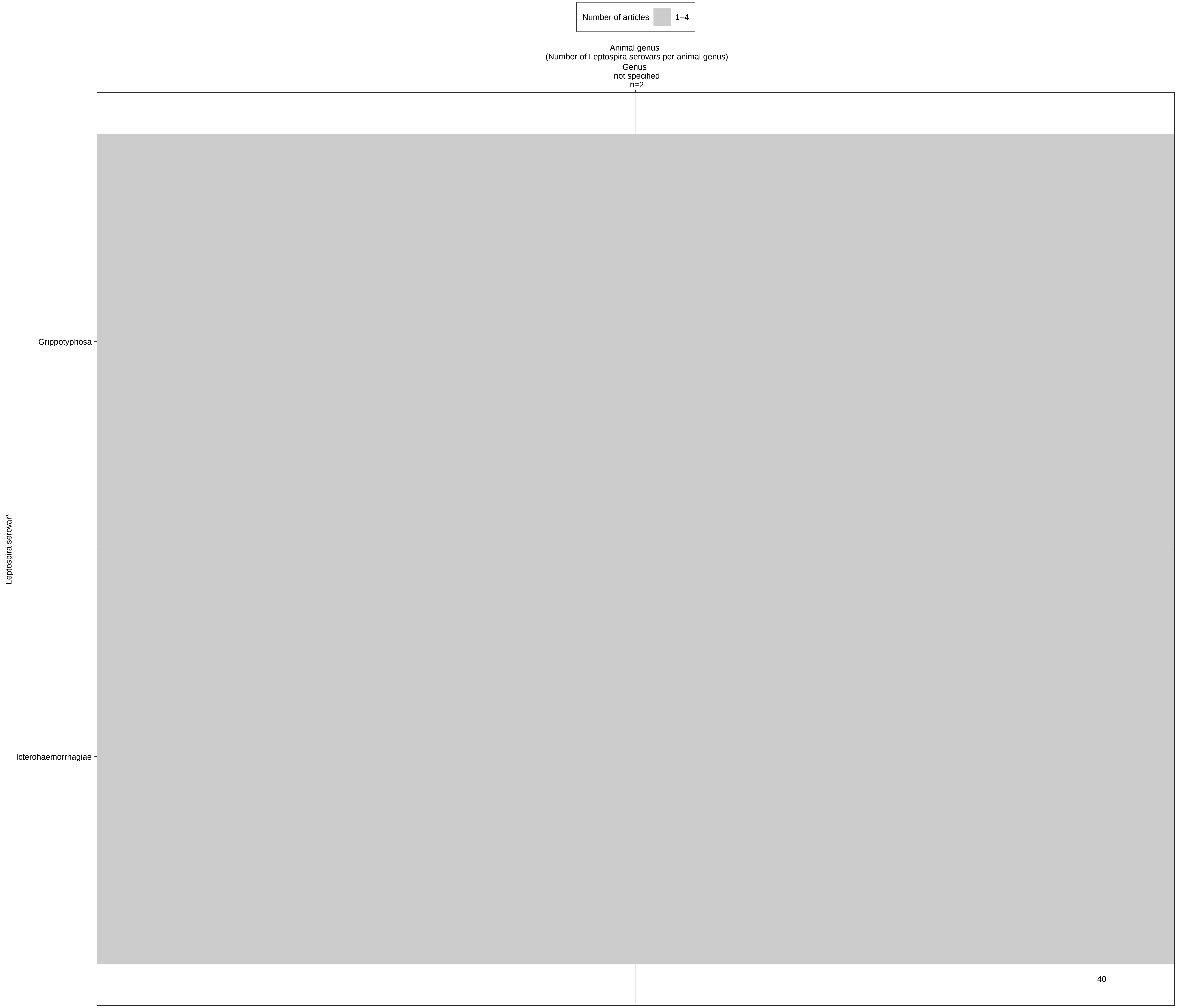

L, Diprotodontia, Ixodidae, Lagomorpha, Primates, Testudines,
all 1 serovar

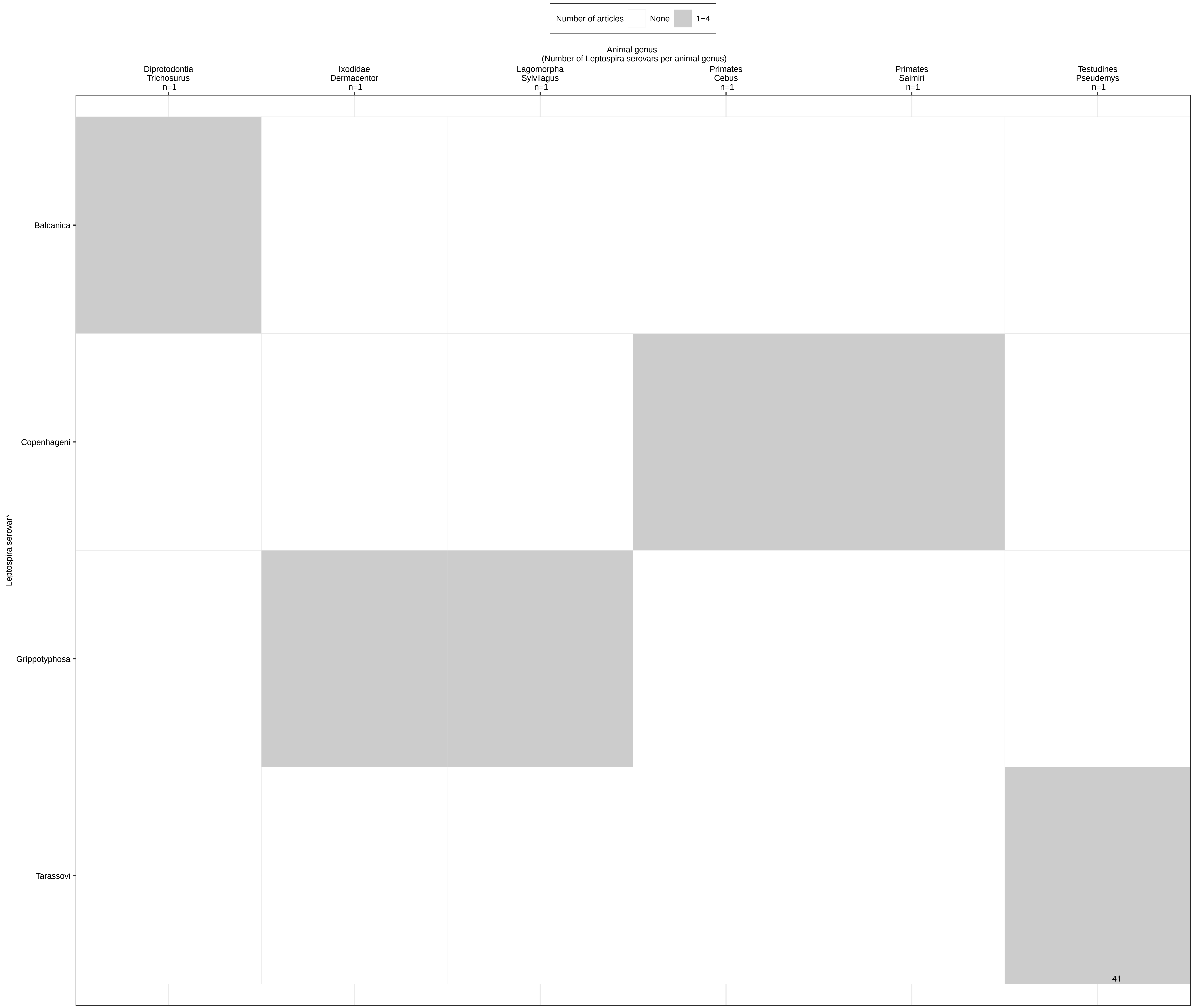

#### Table S9. Table of estimated human leptospirosis morbidity and mortality by Costa *et al.* and number of reports, number of serovars and number of animal hosts identified in the systematic review of reports on *Leptospira* serovar isolations and detections in animal host species published 1927-2022

| **Country** | **Estimated annual morbidity of human leptospirosis per 100,000 people (95% CI) (9)** | **Estimated annual mortality of human leptospirosis per 100,000 people (95% CI)(9)** | **Number of reports Identified** | **Number of unique serovars detected** | **Number of animal hosts defined to genus and species level** |
|---|---|---|---|---|---|
| Sri Lanka | 300.60 (96.54 – 604.23) | 17.98 (6.19 – 0.47) | 2 | 5 | 4 |
| Trinidad and Tobago | 300.48 (95.79 – 607.75) | 16.00 (6.07 – 32.15) | 5 | 11 | 12 |
| Maldives | 273.57 (85.47 – 536.69) | 11.04 (3.63 – 21.21) | *NA* | *NA* | *NA* |
| Solomon Islands | 262.93 (84.11 – 518.76) | 12.11 (4.37 – 22.80) | *NA* | *NA* | *NA* |
| Micronesia, Federated States of | 245.09 (77.30 – 490.19) | 12.42 (4.59 – 22.37) | *NA* | *NA* | *NA* |
| Papua New Guinea | 195.22 (61.38 – 370.88) | 12.48 (4.68 – 24.09) | 1 | 1 | 1 |
| Saint Lucia | 193.16 (63.53 – 373.98) | 8.65 (3.34 – 16.51) | *NA* | *NA* | *NA* |
| Barbados | 146.92 (51.59 – 276.84) | 8.12 (3.19 – 15.26) | 8 | 7 | 8 |
| Antigua and Barbuda | 137.27 (43.33 – 267.49) | 6.68 (2.59 – 12.23) | *NA* | *NA* | *NA* |
| Samoa | 136.69 (47.42 – 274.47) | 7.65 (3.08 – 14.22) | *NA* | *NA* | *NA* |
| Saint Kitts and Nevis | 132.71 (43.10 – 246.48) | 7.00 (2.57 – 13.05) | *NA* | *NA* | *NA* |
| Grenada | 131.53 (46.00 – 243.50) | 7.02 (2.69 – 12.83) | 1 | 6 | 4 |
| Seychelles | 126.64 (43.61 – 234.03) | 7.01 (2.28 – 13.16) | *NA* | *NA* | *NA* |
| Vanuatu | 121.15 (41.99 – 233.46) | 6.41 (2.45 – 11.79) | *NA* | *NA* | *NA* |
| Tonga | 116.07 (38.15 – 225.64) | 5.92 (2.17 – 11.28) | *NA* | *NA* | *NA* |
| Timor-Leste | 114.14 (37.57 – 214.25) | 7.56 (2.67 – 3.21) | *NA* | *NA* | *NA* |
| Saint Vincent and the Grenadines | 107.87 (34.85 – 203.93) | 5.87 (2.19 – 10.15) | *NA* | *NA* | *NA* |
| Kiribati | 106.25 (36.04 – 194.67) | 6.23 (2.28 – 10.96) | *NA* | *NA* | *NA* |
| Mauritius | 81.14 (27.42 – 154.71) | 4.37 (1.54 – 7.94) | *NA* | *NA* | *NA* |
| Comoros | 74.31 (26.97 – 138.97) | 5.43 (1.83 – 9.79) | *NA* | *NA* | *NA* |
| Tuvalu | 74.23 (24.60 – 131.78) | 5.03 (1.86 – 8.72) | *NA* | *NA* | *NA* |
| Palau | 64.06 (20.93 – 120.03) | 3.80 (1.38 – 7.30) | *NA* | *NA* | *NA* |
| Sao Tome | 58.86 (20.59 – 107.14) | 4.21 (1.33 – 7.65) | *NA* | *NA* | *NA* |
| Jamaica | 57.68 (19.64 – 100.48) | 3.18 (1.25 – 5.44) | *NA* | *NA* | *NA* |
| French Guiana | 55.33 (17.46 – 108.36) | 3.24 (1.18 – 6.54) | 2 | 2 | 3 |
| Fiji | 54.38 (18.35 – 101.12) | 3.08 (1.25 – 5.50) | *NA* | *NA* | *NA* |
| Rwanda | 50.27 (15.60 – 108.97) | 3.09 (1.14 – 5.80) | *NA* | *NA* | *NA* |
| Viet Nam | 49.69 (16.47 – 98.92) | 2.09 (0.79 – 4.15) | 3 | 3 | 2 |
| Cape Verde | 44.01 (14.62 – 79.55) | 2.18 (0.76 – 4.00) | *NA* | *NA* | *NA* |
| Dominica | 42.40 (14.75 – 79.03) | 2.45 (0.97 – 4.31) | *NA* | *NA* | *NA* |
| Kenya | 39.46 (13.04 – 79.16) | 2.89 (1.04 – 5.25) | 2 | 4 | 3 |

| | | | | | |
|---|---|---|---|---|---|
| Thailand | 39.37 (14.16 – 77.07) | 2.06 (0.72 – 0.02) | 8 | 8 | 5 |
| Indonesia | 39.20 (12.76 – 77.96) | 2.15 (0.81 – 0.01) | 3 | 4 | 3 |
| Dominican Republic | 38.60 (13.26 – 72.12) | 2.05 (0.82 – 3.72) | NA | NA | NA |
| Costa Rica | 38.06 (12.92 – 74.16) | 1.48 (0.54 – 2.76) | NA | NA | NA |
| Malaysia | 36.98 (11.36 – 73.20) | 1.68 (0.64 – 3.42) | 12 | 19 | 13 |
| Cuba | 36.43 (10.71 – 70.85) | 2.09 (0.83 – 3.75) | 1 | 1 | 1 |
| Ecuador | 35.69 (11.92 – 71.02) | 1.62 (0.50 – 3.12) | NA | NA | NA |
| Uganda | 35.67 (11.57 – 70.67) | 2.79 (1.05 – 5.13) | NA | NA | NA |
| Ethiopia | 34.99 (11.72 – 66.84) | 2.29 (0.90 – 4.12) | NA | NA | NA |
| Cambodia | 33.65 (10.68 – 63.81) | 1.83 (0.65 – 3.35) | NA | NA | NA |
| Burundi | 33.40 (11.30 – 61.92) | 2.87 (1.09 – 5.18) | NA | NA | NA |
| Haiti | 32.47 (10.79 – 59.80) | 2.30 (0.80 – 4.10) | NA | NA | NA |
| Brunei Darussalam | 32.44 (9.60 – 63.48) | 1.33 (0.45 – 2.79) | NA | NA | NA |
| Eritrea | 32.18 (10.84 – 61.14) | 1.48 (0.54 – 2.76) | NA | NA | NA |
| Lichtenstein | 31.44 (9.19 – 71.10) | 1.19 (0.33 – 2.79) | NA | NA | NA |
| Singapore | 31.03 (8.15 – 69.54) | 1.47 (0.43 – 3.72) | NA | NA | NA |
| Panama | 29.49 (9.47 – 56.95) | 1.24 (0.44 – 2.37) | 2 | 17 | 5 |
| Colombia | 27.93 (8.68 – 58.05) | 1.22 (0.40 – 2.51) | 6 | 7 | 7 |
| Suriname | 26.64 (8.68 – 54.19) | 1.37 (0.51 – 2.66) | NA | NA | NA |
| Marshall Islands | 25.71 (9.23 – 47.42) | 2.23 (0.90 – 4.15) | NA | NA | NA |
| Nicaragua | 23.45 (8.00 – 44.34) | 0.91 (0.30 – 1.64) | 2 | 6 | 5 |
| Bahamas | 23.36 (7.27 – 46.23) | 1.23 (0.44 – 2.32) | NA | NA | NA |
| Yemen | 21.78 (6.83 – 40.94) | 1.11 (0.42 – 1.95) | NA | NA | NA |
| United Republic of Tanzania | 20.89 (7.27 – 38.34) | 1.69 (0.64 – 2.90) | 4 | 7 | 5 |
| Nauru | 19.90 (5.48 – 44.02) | 1.93 (0.65 – 3.98) | NA | NA | NA |
| India | 19.69 (6.81 – 36.81) | 1.12 (0.38 – 1.95) | 16 | 9 | 8 |
| Peru | 19.63 (6.38 – 38.68) | 0.79 (0.25 – 1.53) | 10 | 23 | 11 |
| Bangladesh | 19.23 (6.58 – 35.79) | 1.00 (0.30 – 1.82) | NA | NA | NA |
| Lao People's Democratic Republic | 19.11 (6.61 – 34.07) | 1.09 (0.44 – 1.91) | NA | NA | NA |
| Belize | 19.02 (6.80 – 34.59) | 0.81 (0.30 – 1.39) | NA | NA | NA |
| Honduras | 18.61 (6.28 – 33.19) | 0.83 (0.33 – 1.45) | NA | NA | NA |
| Malawi | 18.17 (6.35 – 32.86) | 1.46 (0.58 – 2.50) | NA | NA | NA |
| Equatorial Guinea | 18.02 (6.07 – 33.84) | 1.59 (0.53 – 2.88) | NA | NA | NA |
| Togo | 17.97 (5.88 – 31.42) | 1.18 (0.39 – 2.03) | NA | NA | NA |
| Ghana | 17.73 (6.45 – 32.22) | 1.07 (0.37 – 1.94) | NA | NA | NA |
| Guatemala | 16.77 (5.96 – 29.96) | 0.76 (0.26 – 1.32) | NA | NA | NA |
| Democratic Republic of the Congo | 16.75 (5.65 – 31.65) | 1.63 (0.63 – 2.82) | NA | NA | NA |
| Nepal | 16.70 (5.63 – 32.21) | 0.91 (0.30 – 1.70) | NA | NA | NA |
| Venezuela, Bolivarian Republic of | 15.48 (4.58 – 33.41) | 0.69 (0.25 – 1.32) | NA | NA | NA |

| | | | | | |
|---|---|---|---|---|---|
| El Salvador | 15.40 (5.49 – 27.77) | 0.70 (0.26 – 1.24) | *NA* | *NA* | *NA* |
| Guinea | 15.36 (5.55 – 26.67) | 1.22 (0.42 – 2.10) | *NA* | *NA* | *NA* |
| Philippines | 14.98 (4.94 – 26.91) | 0.74 (0.26 – 1.31) | 7 | 8 | 7 |
| Brazil | 13.77 (4.72 – 27.09) | 0.65 (0.24 – 1.24) | 28 | 23 | 23 |
| Guernsey | 13.57 (4.04 – 27.43) | 0.56 (0.18 – 1.25) | *NA* | *NA* | *NA* |
| Benin | 13.26 (4.73 – 23.61) | 0.96 (0.31 – 1.68) | *NA* | *NA* | *NA* |
| Jersey | 13.18 (4.02 – 26.43) | 0.54 (0.15 – 1.10) | *NA* | *NA* | *NA* |
| Namibia | 12.65 (4.62 – 21.72) | 0.71 (0.29 – 1.19) | *NA* | *NA* | *NA* |
| Paraguay | 12.50 (4.10 – 22.03) | 0.52 (0.20 – 0.88) | *NA* | *NA* | *NA* |
| Burkina Faso | 12.47 (4.52 – 22.60) | 1.09 (0.36 – 1.86) | *NA* | *NA* | *NA* |
| Madagascar | 12.29 (4.40 – 20.80) | 0.78 (0.27 – 1.33) | 1 | 1 | 2 |
| Niger | 11.93 (3.91 – 20.85) | 1.04 (0.38 – 1.78) | *NA* | *NA* | *NA* |
| Senegal | 11.73 (4.10 – 20.59) | 0.76 (0.25 – 1.29) | *NA* | *NA* | *NA* |
| Egypt | 11.65 (4.25 – 20.62) | 0.56 (0.22 – 1.00) | 2 | 6 | 5 |
| Congo | 11.46 (4.15 – 22.36) | 0.99 (0.40 – 1.75) | *NA* | *NA* | *NA* |
| Somalia | 11.39 (4.12 – 20.25) | 1.20 (0.49 – 2.01) | *NA* | *NA* | *NA* |
| United Arab Emirates | 11.37 (3.44 – 22.33) | 0.40 (0.12 – 0.75) | *NA* | *NA* | *NA* |
| Sudan | 11.31 (4.09 – 19.92) | 0.84 (0.35 – 1.40) | *NA* | *NA* | *NA* |
| Bhutan | 11.16 (4.06 – 19.93) | 0.64 (0.21 – 1.09) | *NA* | *NA* | *NA* |
| Cote d'Ivoire | 11.13 (3.91 – 19.76) | 0.90 (0.36 – 1.55) | *NA* | *NA* | *NA* |
| Myanmar | 10.81 (3.71 – 18.72) | 0.90 (0.32 – 1.51) | *NA* | *NA* | *NA* |
| Oman | 10.65 (3.85 – 19.64) | 0.44 (0.14 – 0.79) | *NA* | *NA* | *NA* |
| China | 10.54 (3.52 – 18.39) | 0.50 (0.19 – 0.89) | 11 | 17 | 16 |
| Australia | 10.51 (3.42 – 19.20) | 0.45 (0.16 – 0.79) | 12 | 17 | 18 |
| Central African Republic | 9.87 (3.37 – 16.94) | 1.05 (0.43 – 1.75) | *NA* | *NA* | *NA* |
| Guinea-Bissau | 9.67 (3.50 – 16.71) | 0.97 (0.34 – 1.62) | *NA* | *NA* | *NA* |
| Gabon | 9.63 (3.02 – 20.20) | 0.69 (0.23 – 1.34) | *NA* | *NA* | *NA* |
| Sierra Leone | 9.57 (3.30 – 16.12) | 0.94 (0.32 – 1.56) | *NA* | *NA* | *NA* |
| Cyprus | 9.45 (3.31 – 16.19) | 0.39 (0.13 – 0.69) | *NA* | *NA* | *NA* |
| Cameroon | 9.39 (3.23 – 16.56) | 0.82 (0.30 – 1.46) | *NA* | *NA* | *NA* |
| Tajikistan | 9.22 (2.82 – 18.19) | 0.43 (0.16 – 0.80) | *NA* | *NA* | *NA* |
| Bolivia | 8.94 (3.14 – 15.93) | 0.48 (0.17 – 0.83) | *NA* | *NA* | *NA* |
| Mexico | 8.61 (3.08 – 15.50) | 0.37 (0.14 – 0.67) | 1 | 1 | 1 |
| Mauritania | 8.54 (2.91 – 14.94) | 0.59 (0.21 – 1.01) | *NA* | *NA* | *NA* |
| Portugal | 8.50 (3.13 – 15.41) | 0.38 (0.12 – 0.68) | 6 | 10 | 8 |
| Pakistan | 8.32 (2.86 – 15.14) | 0.50 (0.20 – 0.86) | *NA* | *NA* | *NA* |
| Liberia | 8.10 (2.79 – 14.64) | 0.70 (0.23 – 1.21) | *NA* | *NA* | *NA* |
| Gambia | 8.07 (2.83 – 13.85) | 0.58 (0.21 – 0.99) | *NA* | *NA* | *NA* |
| Slovenia | 7.68 (2.51 – 14.17) | 0.35 (0.11 – 0.65) | *NA* | *NA* | *NA* |

| | | | | | |
|---|---|---|---|---|---|
| Mozambique | 7.54 (2.77 – 12.58) | 0.69 (0.29 – 1.15) | NA | NA | NA |
| Bosnia and Herzegovina | 7.31 (2.41 – 13.43) | 0.36 (0.14 – 0.68) | NA | NA | NA |
| Chad | 7.25 (2.63 – 12.31) | 0.81 (0.28 – 1.31) | NA | NA | NA |
| Algeria | 7.19 (2.64 – 13.02) | 0.33 (0.12 – 0.56) | 1 | 1 | 1 |
| Morocco | 7.19 (2.55 – 12.90) | 0.33 (0.13 – 0.57) | NA | NA | NA |
| Uzbekistan | 7.05 (2.45 – 13.18) | 0.34 (0.13 – 0.60) | NA | NA | NA |
| Kosovo | 6.94 (2.24 – 13.19) | 0.32 (0.12 – 0.61) | NA | NA | NA |
| Nigeria | 6.72 (2.32 – 11.56) | 0.69 (0.24 – 1.14) | 3 | 10 | 2 |
| Republic of Moldova | 6.65 (2.18 – 12.79) | 0.32 (0.11 – 0.60) | 1 | 1 | 1 |
| Libyan Arab Jamahiriya | 6.63 (2.11 – 12.33) | 0.29 (0.10 – 0.52) | NA | NA | NA |
| Mali | 6.60 (2.25 – 11.19) | 0.64 (0.22 – 1.08) | NA | NA | NA |
| Syrian Arab Republic | 6.56 (2.19 – 12.13) | 0.26 (0.09 – 0.46) | NA | NA | NA |
| Azores | 6.55 (2.21 – 11.60) | 0.35 (0.12 – 0.64) | NA | NA | NA |
| Albania | 6.52 (2.22 – 12.20) | 0.31 (0.12 – 0.56) | NA | NA | NA |
| Qatar | 6.44 (1.96 – 13.83) | 0.26 (0.08 – 0.56) | NA | NA | NA |
| Tunisia | 6.41 (2.28 – 11.53) | 0.27 (0.09 – 0.49) | 3 | 2 | 3 |
| Kyrgyzstan | 6.28 (2.26 – 12.00) | 0.34 (0.13 – 0.62) | NA | NA | NA |
| Zambia | 6.12 (2.25 – 10.48) | 0.64 (0.26 – 1.04) | NA | NA | NA |
| Italy | 5.94 (2.01 – 10.86) | 0.26 (0.09 – 0.47) | 8 | 15 | 5 |
| Saudi Arabia | 5.68 (1.95 – 10.41) | 0.27 (0.09 – 0.48) | NA | NA | NA |
| Greece | 5.58 (1.99 – 9.79) | 0.26 (0.09 – 0.45) | NA | NA | NA |
| Botswana | 5.52 (1.89 – 9.58) | 0.35 (0.15 – 0.58) | NA | NA | NA |
| Djibouti | 5.52 (1.90 – 10.30) | 0.41 (0.16 – 0.75) | NA | NA | NA |
| Swaziland | 5.50 (1.99 – 9.93) | 0.54 (0.22 – 0.91) | NA | NA | NA |
| Iran (Islamic Republic of) | 5.47 (2.02 – 9.70) | 0.24 (0.08 – 0.43) | 4 | 2 | 4 |
| Japan | 5.46 (1.84 – 10.09) | 0.25 (0.10 – 0.47) | 10 | 9 | 8 |
| Chile | 5.37 (1.87 – 9.56) | 0.24 (0.09 – 0.44) | 1 | 1 | 1 |
| Croatia | 5.34 (1.71 – 9.73) | 0.27 (0.09 – 0.49) | 4 | 11 | 6 |
| Georgia | 5.13 (1.88 – 9.02) | 0.28 (0.11 – 0.48) | 3 | 2 | 3 |
| Spain | 5.10 (1.85 – 9.10) | 0.22 (0.07 – 0.39) | 3 | 5 | 3 |
| Republic of Korea | 5.02 (1.69 – 8.77) | 0.22 (0.08 – 0.39) | 4 | 3 | 2 |
| Isle of Man | 4.94 (1.53 – 9.62) | 0.23 (0.07 – 0.47) | NA | NA | NA |
| Kuwait | 4.86 (1.56 – 9.68) | 0.19 (0.05 – 0.37) | NA | NA | NA |
| Ireland | 4.79 (1.67 – 9.16) | 0.18 (0.06 – 0.35) | 2 | 2 | 3 |
| Israel | 4.72 (1.55 – 8.87) | 0.19 (0.06 – 0.34) | 6 | 8 | 6 |
| Serbia | 4.65 (1.55 – 8.48) | 0.25 (0.10 – 0.44) | NA | NA | NA |
| Austria | 4.53 (1.57 – 8.27) | 0.21 (0.07 – 0.38) | 1 | 5 | 1 |
| Bahrain | 4.52 (1.37 – 9.18) | 0.20 (0.06 – 0.42) | NA | NA | NA |
| Romania | 4.52 (1.49 – 8.21) | 0.24 (0.09 – 0.43) | 6 | 13 | 7 |

| | | | | | |
|---|---|---|---|---|---|
| Slovakia | 4.46 (1.57 – 8.73) | 0.22 (0.08 – 0.39) | 3 | 2 | 5 |
| The Former Yugoslav Republic of Macedonia | 4.29 (1.47 – 7.87) | 0.22 (0.08 – 0.39) | *NA* | *NA* | *NA* |
| Switzerland | 4.25 (1.44 – 7.66) | 0.19 (0.06 – 0.36) | 1 | 5 | 1 |
| Lesotho | 4.24 (1.51 – 7.70) | 0.44 (0.17 – 0.75) | *NA* | *NA* | *NA* |
| Montenegro | 4.09 (1.47 – 7.32) | 0.21 (0.08 – 0.37) | *NA* | *NA* | *NA* |
| Azerbaijan | 3.98 (1.37 – 6.95) | 0.22 (0.09 – 0.39) | *NA* | *NA* | *NA* |
| Malta | 3.82 (1.26 – 7.18) | 0.18 (0.05 – 0.34) | *NA* | *NA* | *NA* |
| United States of America | 3.81 (1.12 – 6.98) | 0.19 (0.07 – 0.34) | 74 | 24 | 35 |
| San Marino | 3.76 (1.21 – 6.88) | 0.16 (0.05 – 0.30) | *NA* | *NA* | *NA* |
| Turkmenistan | 3.73 (1.32 – 6.86) | 0.23 (0.09 – 0.39) | *NA* | *NA* | *NA* |
| France | 3.67 (1.28 – 6.85) | 0.16 (0.05 – 0.29) | 2 | 5 | 2 |
| Zimbabwe | 3.65 (1.31 – 6.13) | 0.47 (0.19 – 0.77) | 7 | 13 | 1 |
| Jordan | 3.64 (1.28 – 6.63) | 0.16 (0.06 – 0.29) | *NA* | *NA* | *NA* |
| Angola | 3.58 (1.26 – 6.15) | 0.43 (0.15 – 0.73) | *NA* | *NA* | *NA* |
| Andorra | 3.54 (1.13 – 6.51) | 0.15 (0.05 – 0.30) | *NA* | *NA* | *NA* |
| Armenia | 3.51 (1.20 – 6.28) | 0.20 (0.08 – 0.36) | *NA* | *NA* | *NA* |
| Netherlands | 3.50 (1.18 – 6.69) | 0.16 (0.05 – 0.33) | 11 | 10 | 8 |
| New Zealand | 3.48 (1.11 – 6.45) | 0.15 (0.06 – 0.28) | 10 | 6 | 10 |
| Poland | 3.48 (1.08 – 6.95) | 0.17 (0.06 – 0.31) | 4 | 9 | 6 |
| Türkiye | 3.42 (1.19 – 6.12) | 0.16 (0.06 – 0.28) | 2 | 2 | 2 |
| Democratic People’s Republic of Korea | 3.37 (1.17 – 6.01) | 0.20 (0.07 – 0.36) | *NA* | *NA* | *NA* |
| Argentina | 3.34 (1.14 – 6.23) | 0.16 (0.06 – 0.28) | 18 | 12 | 11 |
| Bulgaria | 3.18 (1.07 – 5.72) | 0.19 (0.07 – 0.33) | 3 | 6 | 8 |
| Hungary | 3.14 (0.98 – 5.73) | 0.17 (0.06 – 0.30) | 1 | 1 | 1 |
| Occupied Palestine | 3.11 (1.07 – 5.66) | 0.13 (0.05 – 0.23) | *NA* | *NA* | *NA* |
| Uruguay | 3.08 (1.03 – 5.60) | 0.16 (0.06 – 0.28) | *NA* | *NA* | *NA* |
| Germany | 3.04 (1.00 – 5.81) | 0.15 (0.05 – 0.30) | 8 | 9 | 7 |
| Lebanon | 2.93 (0.92 – 5.37) | 0.15 (0.05 – 0.26) | *NA* | *NA* | *NA* |
| South Africa | 2.83 (1.07 – 5.03) | 0.26 (0.10 – 0.44) | 5 | 4 | 4 |
| Afghanistan | 2.81 (0.95 – 4.96) | 0.37 (0.15 – 0.65) | *NA* | *NA* | *NA* |
| Czech Republic | 2.78 (0.90 – 5.31) | 0.14 (0.04 – 0.26) | *NA* | *NA* | *NA* |
| Iraq | 2.71 (0.95 – 4.79) | 0.17 (0.07 – 0.29) | *NA* | *NA* | *NA* |
| Mongolia | 2.65 (0.87 – 5.07) | 0.14 (0.05 – 0.25) | *NA* | *NA* | *NA* |
| Luxembourg | 2.51 (0.83 – 4.87) | 0.11 (0.04 – 0.22) | 1 | 5 | 1 |
| Denmark | 2.42 (0.76 – 4.83) | 0.12 (0.04 – 0.22) | 2 | 2 | 2 |
| Monaco | 2.38 (0.79 – 4.55) | 0.12 (0.04 – 0.24) | *NA* | *NA* | *NA* |
| Finland | 2.31 (0.72 – 4.75) | 0.11 (0.03 – 0.24) | *NA* | *NA* | *NA* |
| Kazakhstan | 2.29 (0.81 – 4.33) | 0.15 (0.06 – 0.27) | 1 | 1 | 1 |
| United Kingdom of Great Britain and Northern Ireland | 2.24 (0.72 – 4.27) | 0.10 (0.03 – 0.20) | 31 | 14 | 16 |

| | | | | | |
|---|---|---|---|---|---|
| Lithuania | 1.90 (0.61 – 3.80) | 0.11 (0.04 – 0.21) | NA | NA | NA |
| Ukraine | 1.90 (0.67 – 3.58) | 0.12 (0.05 – 0.22) | NA | NA | NA |
| Canada | 1.89 (0.47 – 3.78) | 0.09 (0.03 – 0.18) | 9 | 6 | 9 |
| Estonia | 1.76 (0.59 – 3.60) | 0.09 (0.03 – 0.19) | NA | NA | NA |
| Norway | 1.74 (0.54 – 3.79) | 0.08 (0.02 – 0.16) | NA | NA | NA |
| Latvia | 1.65 (0.53 – 3.30) | 0.10 (0.03 – 0.20) | NA | NA | NA |
| Belarus | 1.59 (0.52 – 3.19) | 0.10 (0.03 – 0.19) | NA | NA | NA |
| Belgium | 1.59 (0.52 – 3.49) | 0.08 (0.02 – 0.15) | 1 | 1 | 1 |
| Sweden | 1.56 (0.46 – 3.17) | 0.07 (0.02 – 0.15) | NA | NA | NA |
| Iceland | 1.17 (0.35 – 2.46) | 0.05 (0.01 – 0.10) | NA | NA | NA |
| Russian Federation | 1.08 (0.31 – 2.34) | 0.07 (0.02 – 0.15) | 12 | 12 | 11 |
| American Samoa | NA | NA | 1 | 1 | 1 |
| Guadeloupe | NA | NA | 2 | 5 | 3 |
| Puerto Rico | NA | NA | 2 | 4 | 7 |

NA, not available

#### Table S10. Summary tables of *Leptospira* serovar isolations and detections in animal host species published 1927-2022, sorted by serogroup, serovar and animal genus and species

| Serogroup | Serovar | Animal genus and species | Animal common name | Animal order | Animal class | UN country | Reference |
|---|---|---|---|---|---|---|---|
| **Australis** | **Australis** | *Apodemus agrarius* | Striped field mouse | Rodentia | Mammalia | China | (1) |
| **Australis** | **Australis** | *Arvicanthus niloticus* | African grass rat | Rodentia | Mammalia | Nigeria | (2) |
| **Australis** | **Australis** | *Bos taurus* | Cattle | Artiodactyla | Mammalia | Japan, Malaysia | (3-7) |
| **Australis** | **Australis** | *Canis lupus* | Dog | Carnivora | Mammalia | Italy | (8) |
| **Australis** | **Australis** | *Equus caballus* | Horse | Perissodactyla | Mammalia | Philippines | (9) |
| **Australis** | **Australis** | *Erinaceus europaeus* | Hedgehog | Eulipotyphla | Mammalia | Belgium, Israel, Italy | (10-12) |
| **Australis** | **Australis** | *Herpestes auropunctatus* | Lesser Indian mongoose | Carnivora | Mammalia | Guadeloupe | (13) |
| **Australis** | **Australis** | *Isoodon macrourus* | Northern brown bandicoot | Peramelemorphia | Mammalia | Australia | (14) |
| **Australis** | **Australis** | *Mus musculus* | House mouse | Rodentia | Mammalia | Australia | (14) |
| **Australis** | **Australis** | *Myocastor coypus* | Nutria | Rodentia | Mammalia | China | (15) |
| **Australis** | **Australis** | *Nectomys squamipes* | South American water rat | Rodentia | Mammalia | Brazil | (16) |
| **Australis** | **Australis** | *Perameles nasuta* | Long-nosed bandicoot | Peramelemorphia | Mammalia | Australia | (14) |
| **Australis** | **Australis** | *Procyon lotor* | Raccoon | Carnivora | Mammalia | United States of America | (17, 18) |
| **Australis** | **Australis** | *Procyon spp.* | Raccoon | Carnivora | Mammalia | United States of America | (19) |
| **Australis** | **Australis** | *Rat spp.* | Field rat | Rodentia | Mammalia | Thailand | (20, 21) |

| | | | | | | | |
|---|---|---|---|---|---|---|---|
| **Australis** | **Australis** | *Rat spp.* | Rat | Rodentia | Mammalia | Thailand | (22) |
| **Australis** | **Australis** | *Rattus fuscipes* | Bush rat | Rodentia | Mammalia | Australia | (14, 23) |
| **Australis** | **Australis** | *Rattus leucopus* | Cape York rat | Rodentia | Mammalia | Australia | (23) |
| **Australis** | **Australis** | *Rattus losea* | Lesser rice field rat | Rodentia | Mammalia | China | (1) |
| **Australis** | **Australis** | *Rattus norvegicus* | Norwegian rat | Rodentia | Mammalia | American Samoa, China, United States of America | (1, 24, 25) |
| **Australis** | **Australis** | *Rattus rattus* | Black rat | Rodentia | Mammalia | Australia | (14) |
| **Australis** | **Australis** | *Rattus sordidus* | Canefield rat | Rodentia | Mammalia | Australia | (14, 23) |
| **Australis** | **Australis** | *Rattus spp.* | Rat | Rodentia | Mammalia | Australia, China | (15, 23, 26) |
| **Australis** | **Australis** | *Sundamys muelleri* | Mueller's rat | Rodentia | Mammalia | Malaysia | (27) |
| **Australis** | **Australis** | *Uromys caudimaculatus* | Giant white-tailed rat | Rodentia | Mammalia | Australia | (28) |
| **Australis** | **Australis** | *Uromys spp.* | Giant naked-tailed rats | Rodentia | Mammalia | Australia | (23) |
| **Australis** | **Australis** | Not stated | Rodent | Rodentia | Mammalia | Australia, Poland | (29, 30) |
| **Australis** | **Bajan** | *Bufo marinus* | Giant marine toad | Anura | Amphibia | Barbados | (31) |
| **Australis** | **Bajan** | *Eleutherodactylus johnstonei* | Whistling frog | Anura | Amphibia | Barbados | (31, 32) |
| **Australis** | **Bangkok** | *Canis lupus* | Dog | Carnivora | Mammalia | Thailand | (33) |
| **Australis** | **Bratislava** | *Apodemus flavicollis* | Yellow-necked field mouse | Rodentia | Mammalia | Croatia | (34) |
| **Australis** | **Bratislava** | *Apodemus sylvaticus* | Common field mouse | Rodentia | Mammalia | Croatia | (34) |
| **Australis** | **Bratislava** | *Bos taurus* | Cattle | Artiodactyla | Mammalia | Nigeria | (35) |
| **Australis** | **Bratislava** | *Equus caballus* | Horse | Perissodactyla | Mammalia | Austria, Germany, Italy, Luxembourg, Netherlands, Portugal, Switzerland, United Kingdom of Great Britain and Northern Ireland | (36-39) |
| **Australis** | **Bratislava** | *Erinaceus europaeus* | European hedgehog | Eulipotyphla | Mammalia | Bulgaria | (40) |

| | | | | | | | |
|---|---|---|---|---|---|---|---|
| **Australis** | **Bratislava** | *Erinaceus europaeus* | Hedgehog | Eulipotyphla | Mammalia | Italy, Netherlands, Russian Federation, United Kingdom of Great Britain and Northern Ireland | (11, 39, 41, 42) |
| **Australis** | **Bratislava** | *Ovis aries* | Sheep | Artiodactyla | Mammalia | United Kingdom of Great Britain and Northern Ireland | (39, 43) |
| **Australis** | **Bratislava** | *Rattus norvegicus* | Norwegian rat | Rodentia | Mammalia | United Kingdom of Great Britain and Northern Ireland | (39) |
| **Australis** | **Bratislava** | *Sus scrofa* | Pig | Artiodactyla | Mammalia | Brazil, Germany, United Kingdom of Great Britain and Northern Ireland, United States of America | (44-49) |
| **Australis** | **Fugis** | *Bos taurus* | Cattle | Artiodactyla | Mammalia | Zimbabwe | (50) |
| **Australis** | **Jalna** | *Apodemus flavicollis* | Yellow-necked field mouse | Rodentia | Mammalia | Bulgaria, Croatia | (40, 51) |
| **Australis** | **Jalna** | *Apodemus mystacinus* | Eastern broad toothed field mouse | Rodentia | Mammalia | Bulgaria | (40) |
| **Australis** | **Jalna** | *Apodemus sylvaticus* | Common field mouse | Rodentia | Mammalia | Bulgaria, Croatia, Poland | (40, 51, 52) |
| **Australis** | **Jalna** | *Arvicola terrestris* | European water vole | Rodentia | Mammalia | Poland | (52) |
| **Australis** | **Jalna** | *Sorex araneus* | Common shrew | Eulipotyphla | Mammalia | Poland | (52) |
| **Australis** | **Jalna** | *Sorex minutus* | Eurasian pygmy shrew | Eulipotyphla | Mammalia | Poland | (52) |
| **Australis** | **Lora** | *Apodemus flavicollis* | Yellow-necked field mouse | Rodentia | Mammalia | Bulgaria, Croatia | (40, 53) |
| **Australis** | **Lora** | *Apodemus mystacinus* | Eastern broad toothed field mouse | Rodentia | Mammalia | Georgia | (54) |
| **Australis** | **Lora** | *Apodemus sylvaticus* | Common field mouse | Rodentia | Mammalia | Bulgaria, Georgia | (40, 55, 56) |
| **Australis** | **Lora** | *Clethrionomys glareolus* | Bank vole | Rodentia | Mammalia | Croatia | (57) |
| **Australis** | **Lora** | *Equus caballus* | Horse | Perissodactyla | Mammalia | Austria, Germany, Italy, Luxembourg, Netherlands, Portugal, Switzerland, United Kingdom of Great Britain and Northern Ireland | (36, 58) |
| **Australis** | **Lora** | *Mastomys spp.* | Multimammate rat | Rodentia | Mammalia | United Republic of Tanzania | (59) |
| **Australis** | **Lora** | *Microtus arvalis* | Common vole | Rodentia | Mammalia | Bulgaria | (40, 56) |
| **Australis** | **Lora** | *Mus musculus* | House mouse | Rodentia | Mammalia | Bulgaria | (56) |

| | | | | | | | |
|---|---|---|---|---|---|---|---|
| **Australis** | **Lora** | *Ondatra zibethicus* | Muskrat | Rodentia | Mammalia | Netherlands | (60) |
| **Australis** | **Lora** | *Rattus rattus* | Black rat | Rodentia | Mammalia | Bulgaria | (40) |
| **Australis** | **Lora** | *Sus scrofa* | Pig | Artiodactyla | Mammalia | Netherlands | (61) |
| **Australis** | **Muenchen** | *Apodemus flavicollis* | Yellow-necked field mouse | Rodentia | Mammalia | Croatia | (34) |
| **Australis** | **Muenchen** | *Apodemus sylvaticus* | Common field mouse | Rodentia | Mammalia | Croatia, United Kingdom of Great Britain and Northern Ireland | (34, 39) |
| **Australis** | **Muenchen** | *Clethrionomys glareolus* | Bank vole | Rodentia | Mammalia | United Kingdom of Great Britain and Northern Ireland | (39, 62) |
| **Australis** | **Muenchen** | *Equus caballus* | Horse | Perissodactyla | Mammalia | Austria, Germany, Italy, Luxembourg, Netherlands, Portugal, Switzerland, United Kingdom of Great Britain and Northern Ireland | (36) |
| **Australis** | **Muenchen** | *Microtus agrestis* | Short tailed vole | Rodentia | Mammalia | United Kingdom of Great Britain and Northern Ireland | (39, 62) |
| **Australis** | **Muenchen** | *Sciurus carolinensis* | Grey squirrel | Rodentia | Mammalia | United Kingdom of Great Britain and Northern Ireland | (39) |
| **Australis** | **Muenchen** | *Sus scrofa* | Pig | Artiodactyla | Mammalia | United Kingdom of Great Britain and Northern Ireland | (39, 44-46, 63, 64) |
| **Australis** | **Nicaragua** | *Mustela spp.* | Weasel | Carnivora | Mammalia | Nicaragua | (65) |
| **Australis** | **Peruviana** | *Bufo marinus* | Giant marine toad | Anura | Amphibia | Grenada | (66) |
| **Australis** | **Rushan** | *Bombina orientalis* | Oriental fire bellied toad | Anura | Amphibia | China | (67) |
| **Australis** | **Soteropolitana** | *Cavia aperea* | Brazilian guinea pig | Rodentia | Mammalia | Brazil | (68) |
| **Australis** | **Soteropolitana** | *Zygodontomys lasiurus* | Hairy tailed akodont | Rodentia | Mammalia | Brazil | (68) |
| **Autumnalis** | **Autumnalis** | *Apodemus speciosus* | Large japanese field mouse | Rodentia | Mammalia | Japan | (69) |
| **Autumnalis** | **Autumnalis** | *Bandicota bengalensis* | Lesser bandicoot rat | Rodentia | Mammalia | India, Thailand | (70, 71) |
| **Autumnalis** | **Autumnalis** | *Bos taurus* | Cattle | Artiodactyla | Mammalia | Japan | (6, 7) |
| **Autumnalis** | **Autumnalis** | *Bufo marinus* | Giant marine toad | Anura | Amphibia | Trinidad and Tobago | (66, 72) |
| **Autumnalis** | **Autumnalis** | *Procyon lotor* | Raccoon | Carnivora | Mammalia | United States of America | (73) |

| | | | | | | | |
|---|---|---|---|---|---|---|---|
| **Autumnalis** | **Autumnalis** | *Rat spp.* | Field rat | Rodentia | Mammalia | Thailand | (20, 21) |
| **Autumnalis** | **Autumnalis** | *Rat spp.* | Rat | Rodentia | Mammalia | Thailand | (22) |
| **Autumnalis** | **Autumnalis** | *Rattus rattus* | Black rat | Rodentia | Mammalia | Barbados, India, Thailand | (70, 74-76) |
| **Autumnalis** | **Bim** | *Bufo marinus* | Giant marine toad | Anura | Amphibia | Barbados | (31, 77) |
| **Autumnalis** | **Bim** | *Canis lupus* | Dog | Carnivora | Mammalia | Barbados | (78) |
| **Autumnalis** | **Bim** | *Eleutherodactylus johnstonei* | Whistling frog | Anura | Amphibia | Barbados | (31, 32) |
| **Autumnalis** | **Bim** | *Rattus norvegicus* | Norwegian rat | Rodentia | Mammalia | Barbados | (79) |
| **Autumnalis** | **Bulgarica** | *Rattus rattus* | Black rat | Rodentia | Mammalia | China | (80) |
| **Autumnalis** | **Fortbragg** | *Herpestes auropunctatus* | Lesser Indian mongoose | Carnivora | Mammalia | Barbados | (76, 81) |
| **Autumnalis** | **Fortbragg** | *Rattus norvegicus* | Norwegian rat | Rodentia | Mammalia | Barbados | (76) |
| **Autumnalis** | **Fortbragg** | *Rattus rattus* | Black rat | Rodentia | Mammalia | Barbados | (76) |
| **Autumnalis** | **Fortbragg** | *Rattus rattus* | Rat | Rodentia | Mammalia | Barbados | (82) |
| **Autumnalis** | **Fortbragg** | *Rattus spp.* | Rat | Rodentia | Mammalia | Barbados | (81) |
| **Autumnalis** | **Lambwe** | *Arvicanthus niloticus* | African grass rat | Rodentia | Mammalia | Kenya | (83) |
| **Autumnalis** | **Mooris** | *Rattus rattus* | Black rat | Rodentia | Mammalia | China | (80) |
| **Ballum** | **Arborea** | *Erinaceus europaeus* | Hedgehog | Eulipotyphla | Mammalia | Italy | (11) |
| **Ballum** | **Arborea** | *Mus musculus* | House mouse | Rodentia | Mammalia | Argentina, Portugal | (84-86) |
| **Ballum** | **Arborea** | *Mus spretus* | Algerian mouse | Rodentia | Mammalia | Portugal | (87) |
| **Ballum** | **Arborea** | *Rattus norvegicus* | Norwegian rat | Rodentia | Mammalia | Argentina, Barbados, Portugal | (79, 86, 87) |
| **Ballum** | **Arborea** | *Rattus rattus* | Black rat | Rodentia | Mammalia | Barbados, Israel, Portugal | (79, 85, 87, 88) |

| | | | | | | | |
|---|---|---|---|---|---|---|---|
| **Ballum** | **Arborea** | *Rattus spp.* | Rat | Rodentia | Mammalia | Australia | (89) |
| **Ballum** | **Arborea** | Not stated | Rodent | Rodentia | Mammalia | Australia | (23) |
| **Ballum** | **Ballum** | *Akodon arviculoides* | Montane grass mouse | Rodentia | Mammalia | Brazil | (90) |
| **Ballum** | **Ballum** | *Arvicanthus niloticus* | African grass rat | Rodentia | Mammalia | Nigeria | (2) |
| **Ballum** | **Ballum** | *Blarina brevicauda* | Short tailed shrew | Eulipotyphla | Mammalia | United States of America | (91) |
| **Ballum** | **Ballum** | *Bos taurus* | Cattle | Artiodactyla | Mammalia | Malaysia, New Zealand | (4, 92) |
| **Ballum** | **Ballum** | *Caluromys philander* | Bare-tailed woolly opossum | Didelphimorphia | Mammalia | Trinidad and Tobago | (66, 72) |
| **Ballum** | **Ballum** | *Canis lupus* | Dog | Carnivora | Mammalia | United States of America | (93) |
| **Ballum** | **Ballum** | *Didelphis marsupialis* | Common opossum | Didelphimorphia | Mammalia | United States of America | (18, 94-96) |
| **Ballum** | **Ballum** | *Didelphus virginiana* | Virginia opossum | Didelphimorphia | Mammalia | United States of America | (97, 98) |
| **Ballum** | **Ballum** | *Erinaceus europaeus* | Hedgehog | Eulipotyphla | Mammalia | Israel, New Zealand | (99, 100) |
| **Ballum** | **Ballum** | *Felis rufa* | Wild cat | Carnivora | Mammalia | United States of America | (18) |
| **Ballum** | **Ballum** | *Herpestes auropunctatus* | Lesser Indian mongoose | Carnivora | Mammalia | United States of America | (24) |
| **Ballum** | **Ballum** | *Heterodon platyrhinus* | Eastern hog-nosed snake | Squamata | Reptilia | United States of America | (94) |
| **Ballum** | **Ballum** | *Marmosa mitis* | Pouchless opossum | Didelphimorphia | Mammalia | Trinidad and Tobago | (66, 72) |
| **Ballum** | **Ballum** | *Mephitis mephitis* | Striped skunk | Carnivora | Mammalia | United States of America | (18, 95, 101-104) |
| **Ballum** | **Ballum** | *Microtus pennsylvanicus* | Meadow vole | Rodentia | Mammalia | United States of America | (105) |
| **Ballum** | **Ballum** | *Mus musculus* | House mouse | Rodentia | Mammalia | Brazil, Denmark, Guadeloupe, Mexico, New Zealand, Trinidad and Tobago, United States of America | (13, 16, 24, 72, 94, 98, 100, 106-113) |
| **Ballum** | **Ballum** | *Peromyscus maniculatus* | Deer mouse | Rodentia | Mammalia | United States of America | (94) |
| **Ballum** | **Ballum** | *Peromyscus polionotus* | Oldfield mouse | Rodentia | Mammalia | United States of America | (109) |

| | | | | | | | |
|---|---|---|---|---|---|---|---|
| **Ballum** | **Ballum** | *Philander opossum* | Gray four-eyed opossum | Didelphimorphia | Mammalia | Brazil | (16) |
| **Ballum** | **Ballum** | *Procyon lotor* | Raccoon | Carnivora | Mammalia | United States of America | (18) |
| **Ballum** | **Ballum** | *Rattus exulans* | Polynesian rat | Rodentia | Mammalia | United States of America | (24, 106) |
| **Ballum** | **Ballum** | *Rattus norvegicus* | Norwegian rat | Rodentia | Mammalia | Israel, Italy, New Zealand, Puerto Rico, Spain, United States of America | (24, 100, 111, 113-118) |
| **Ballum** | **Ballum** | *Rattus rattus* | Black rat | Rodentia | Mammalia | Grenada, Israel, New Zealand, Trinidad and Tobago, United States of America | (24, 66, 72, 88, 106, 111, 113, 118) |
| **Ballum** | **Ballum** | *Rattus spp.* | Rat | Rodentia | Mammalia | Canada | (119) |
| **Ballum** | **Ballum** | *Urocyon cinereoargenteus* | Gray fox | Carnivora | Mammalia | United States of America | (18) |
| **Ballum** | **Ballum** | Not stated | Opossum | Didelphimorphia | Mammalia | United States of America | (120) |
| **Ballum** | **Castellonis** | *Mus musculus* | House mouse | Rodentia | Mammalia | Barbados | (76) |
| **Ballum** | **Castellonis** | *Ovis aries* | Sheep | Artiodactyla | Mammalia | Argentina | (121) |
| **Ballum** | **Kenya** | *Cricetomys gambianus* | Giant pouched rat | Rodentia | Mammalia | United Republic of Tanzania | (122) |
| **Ballum** | **Kenya** | *Cricetomys spp.* | Giant pouched rat | Rodentia | Mammalia | United Republic of Tanzania | (59) |
| **Ballum** | **Kenya** | *Crocidura spp.* | Shrew | Eulipotyphla | Mammalia | United Republic of Tanzania | (59) |
| **Ballum** | **Kenya** | *Mastomys spp.* | Multimammate rat | Rodentia | Mammalia | United Republic of Tanzania | (59) |
| **Ballum** | **Kenya** | *Saccostomus campestris* | South African pouched mouse | Rodentia | Mammalia | Kenya | (83) |
| **Bataviae** | **Argentinensis** | *Chaetophractus villosus* | Big hairy armadillo | Cingulata | Mammalia | Argentina | (123-125) |
| **Bataviae** | **Balboa** | *Proechimys semispinosus* | Spiny rat | Rodentia | Mammalia | Panama | (65) |
| **Bataviae** | **Bataviae** | *Apodemus agrarius* | Striped field mouse | Rodentia | Mammalia | Croatia | (34) |
| **Bataviae** | **Bataviae** | *Arvicola terrestris* | European water vole | Rodentia | Mammalia | Bulgaria | (126) |
| **Bataviae** | **Bataviae** | *Chaetophractus villosus* | Big hairy armadillo | Cingulata | Mammalia | Argentina | (123, 124) |

| | | | | | | | |
|---|---|---|---|---|---|---|---|
| **Bataviae** | **Bataviae** | *Felis catus* | Cat | Carnivora | Mam malia | Indonesia, Malaysia | (127, 128) |
| **Bataviae** | **Bataviae** | *Leopoldamys sabanus* | Andaman rat | Rodentia | Mam malia | Malaysia | (27) |
| **Bataviae** | **Bataviae** | *Rat spp.* | Field rat | Rodentia | Mam malia | Thailand | (20, 21) |
| **Bataviae** | **Bataviae** | *Rat spp.* | Rat | Rodentia | Mam malia | Thailand | (22) |
| **Bataviae** | **Bataviae** | *Rattus exulans* | Polynesian rat | Rodentia | Mam malia | Malaysia | (129, 130) |
| **Bataviae** | **Bataviae** | *Rattus norvegicus* | Norwegian rat | Rodentia | Mam malia | Malaysia, Viet Nam | (129-131) |
| **Bataviae** | **Bataviae** | *Rattus rattus* | Black rat | Rodentia | Mam malia | Malaysia | (129, 130, 132) |
| **Bataviae** | **Bataviae** | *Rattus spp.* | Rat | Rodentia | Mam malia | China, Philippines, Thailand | (26, 133, 134) |
| **Bataviae** | **Bataviae** | *Sundamys muelleri* | Mueller's rat | Rodentia | Mam malia | Malaysia | (27) |
| **Bataviae** | **Bataviae** | *Sus scrofa* | Pig | Artiodact yla | Mam malia | Peru | (135) |
| **Bataviae** | **Bataviae** | Not stated | Rodent | Rodentia | Mam malia | Poland | (29) |
| **Bataviae** | **Brasiliensis** | *Didelphis marsupialis* | Common opossum | Didelphi morphia | Mam malia | Brazil | (136) |
| **Bataviae** | **Brasiliensis** | *Herpestes auropunctatus* | Lesser Indian mongoose | Carnivora | Mam malia | Grenada | (66) |
| **Bataviae** | **Brasiliensis** | *Sus scrofa* | Pig | Artiodact yla | Mam malia | United Kingdom of Great Britain and Northern Ireland | (63) |
| **Bataviae** | **Claytoni** | *Proechimys semispinosus* | Spiny rat | Rodentia | Mam malia | Panama | (65) |
| **Bataviae** | **Djatzi** | *Herpestes auropunctatus* | Lesser Indian mongoose | Carnivora | Mam malia | Puerto Rico | (115) |
| **Bataviae** | **Djatzi** | *Mus musculus* | House mouse | Rodentia | Mam malia | Puerto Rico | (115) |
| **Bataviae** | **Djatzi** | *Rattus rattus* | Black rat | Rodentia | Mam malia | Puerto Rico | (115) |
| **Bataviae** | **Kobbe** | *Proechimys semispinosus* | Spiny rat | Rodentia | Mam malia | Panama | (137) |
| **Bataviae** | **Kobbe** | *Sus scrofa* | Pig | Artiodact yla | Mam malia | Peru | (138) |

| | | | | | | | |
|---|---|---|---|---|---|---|---|
| **Bataviae** | **Losbanos** | *Rat spp.* | Rat | Rodentia | Mammalia | Philippines | (139) |
| **Bataviae** | **Losbanos** | *Rattus norvegicus* | Norwegian rat | Rodentia | Mammalia | Philippines | (140) |
| **Bataviae** | **Losbanos** | *Rattus tanezumi* | Oriental house rat | Rodentia | Mammalia | Philippines | (140) |
| **Bataviae** | **Paidjan** | *Bos taurus* | Cattle | Artiodactyla | Mammalia | Zimbabwe | (50, 141) |
| **Bataviae** | **Paidjan** | *Canis lupus* | Dog | Carnivora | Mammalia | Thailand | (142) |
| **Bataviae** | **Paidjan** | *Chaetophractus villosus* | Big hairy armadillo | Cingulata | Mammalia | Argentina | (123, 124) |
| **Bataviae** | **Paidjan** | *Didelphis albiventris* | White eared opossum | Didelphimorphia | Mammalia | Argentina | (143) |
| **Bataviae** | **Paidjan** | *Myocastor coypus* | Nutria | Rodentia | Mammalia | United States of America | (144) |
| **Bataviae** | **Rioja** | *Philander opossum* | Gray four-eyed opossum | Didelphimorphia | Mammalia | Peru | (145) |
| **Canicola** | **Benjamini** | *Rattus exulans* | Polynesian rat | Rodentia | Mammalia | Malaysia | (27) |
| **Canicola** | **Bindjei** | *Melomys cervinipes* | Fawn-footed mosaic-tailed rat | Rodentia | Mammalia | Australia | (28) |
| **Canicola** | **Bindjei** | *Melomys lutillus* | Papua grassland mosaic-tailed rat | Rodentia | Mammalia | Australia | (14) |
| **Canicola** | **Broomi** | *Isoodon macrourus* | Northern brown bandicoot | Peramelemorphia | Mammalia | Australia | (14) |
| **Canicola** | **Broomi** | *Rattus rattus* | Black rat | Rodentia | Mammalia | Australia | (14) |
| **Canicola** | **Canicola** | *Akodon azarae* | Azara's grass mouse | Rodentia | Mammalia | Argentina | (146) |
| **Canicola** | **Canicola** | *Bos taurus* | Cattle | Artiodactyla | Mammalia | Argentina, Brazil, Malaysia, Nigeria, United States of America | (4, 5, 35, 147-150) |
| **Canicola** | **Canicola** | *Canis lupus* | Dog | Carnivora | Mammalia | Argentina, Brazil, China, Georgia, India, Indonesia, Italy, Japan, Malaysia, Netherlands, Russian Federation, South Africa, Trinidad and Tobago, United States of America | (1, 72, 130, 151-173) |
| **Canicola** | **Canicola** | *Chaetophractus villosus* | Big hairy armadillo | Cingulata | Mammalia | Argentina | (123) |
| **Canicola** | **Canicola** | *Conepatus semistriatus* | Hog nosed skunk | Carnivora | Mammalia | Nicaragua | (174) |

| | | | | | | | |
|---|---|---|---|---|---|---|---|
| **Canicola** | **Canicola** | *Cricetomys spp.* | Giant pouched rat | Rodentia | Mammalia | United Republic of Tanzania | (59) |
| **Canicola** | **Canicola** | *Erinaceus europaeus* | Hedgehog | Eulipotyphla | Mammalia | Israel | (12, 99) |
| **Canicola** | **Canicola** | *Felis catus* | Cat | Carnivora | Mammalia | Trinidad and Tobago | (72, 168) |
| **Canicola** | **Canicola** | *Hemiechinus auritus* | Long eared hedgehog | Eulipotyphla | Mammalia | Israel | (99) |
| **Canicola** | **Canicola** | *Herpestes auropunctatus* | Lesser Indian mongoose | Carnivora | Mammalia | Trinidad and Tobago | (66, 72) |
| **Canicola** | **Canicola** | *Mephitis mephitis* | Striped skunk | Carnivora | Mammalia | United States of America | (102, 103, 175) |
| **Canicola** | **Canicola** | *Rattus exulans* | Polynesian rat | Rodentia | Mammalia | Malaysia | (27) |
| **Canicola** | **Canicola** | *Rattus norvegicus* | Norwegian rat | Rodentia | Mammalia | Brazil | (176, 177) |
| **Canicola** | **Canicola** | *Rattus rattus* | Black rat | Rodentia | Mammalia | Egypt | (178) |
| **Canicola** | **Canicola** | *Rattus spp.* | Rat | Rodentia | Mammalia | China | (26) |
| **Canicola** | **Canicola** | *Spilogale putorious* | Eastern spotted skunk | Carnivora | Mammalia | Nicaragua | (174) |
| **Canicola** | **Canicola** | *Sus scrofa* | Pig | Artiodactyla | Mammalia | Brazil, Malaysia, Peru, South Africa, Sri Lanka, United States of America | (162, 172, 173, 179-181) |
| **Canicola** | **Canicola** | *Urocyon cinereoargenteus* | Gray fox | Carnivora | Mammalia | Nicaragua | (174) |
| **Canicola** | **Galtoni** | *Bos taurus* | Cattle | Artiodactyla | Mammalia | Argentina | (182) |
| **Canicola** | **Kuwait** | *Rattus norvegicus* | Norwegian rat | Rodentia | Mammalia | Madagascar | (183) |
| **Canicola** | **Kuwait** | *Rattus rattus* | Black rat | Rodentia | Mammalia | Madagascar | (183) |
| **Canicola** | **Portlandvere** | *Canis lupus* | Dog | Carnivora | Mammalia | Trinidad and Tobago | (72, 168) |
| **Canicola** | **Schueffneri** | *Rattus exulans* | Polynesian rat | Rodentia | Mammalia | Malaysia | (27) |
| **Canicola** | **Schueffneri** | *Rattus rajah* | Red spiny rat | Rodentia | Mammalia | Malaysia | (71) |
| **Celledoni** | **Celledoni** | *Isoodon macrourus* | Northern brown bandicoot | Peramelemorphia | Mammalia | Australia | (14, 28) |

| | | | | | | | |
|---|---|---|---|---|---|---|---|
| **Celledoni** | **Celledoni** | *Melomys cervinipes* | Giant white-tailed rat | Rodentia | Mammalia | Australia | (14) |
| **Celledoni** | **Celledoni** | *Rattus argentiventer* | Rice-field rat | Rodentia | Mammalia | Malaysia | (27) |
| **Celledoni** | **Celledoni** | *Rattus fuscipes* | Bush rat | Rodentia | Mammalia | Australia | (14) |
| **Cynopteri** | **Tingomaria** | *Didelphis marsupialis* | Common opossum | Didelphimorphia | Mammalia | Peru | (145) |
| **Cynopteri** | **Tingomaria** | *Philander opossum* | Gray four-eyed opossum | Didelphimorphia | Mammalia | Peru | (145) |
| **Djasiman** | **Buenos Aires** | *Canis lupus* | Dog | Carnivora | Mammalia | Argentina | (184) |
| **Djasiman** | **Djasiman** | *Rattus bowersi* | Bower's white toothed rat | Rodentia | Mammalia | Malaysia | (27) |
| **Djasiman** | **Djasiman** | *Spermophilus citellus* | European ground squirrel | Rodentia | Mammalia | Türkiye | (185) |
| **Djasiman** | **Huallaga** | *Didelphis marsupialis* | Common opossum | Didelphimorphia | Mammalia | Peru | (145) |
| **Djasiman** | **Sentot** | *Paradoxurus hermaphroditus* | Asian palm civet | Carnivora | Mammalia | Malaysia | (27) |
| **Grippotyphosa** | **Canalzonae** | *Liomys adspersus* | Spiny pocket mouse | Rodentia | Mammalia | Panama | (137) |
| **Grippotyphosa** | **Canalzonae** | *Philander opossum* | Gray four-eyed opossum | Didelphimorphia | Mammalia | Panama | (137) |
| **Grippotyphosa** | **Canalzonae** | *Proechimys semispinosus* | Spiny rat | Rodentia | Mammalia | Panama | (137) |
| **Grippotyphosa** | **Dadas** | *Apodemus flavicollis* | Yellow-necked field mouse | Rodentia | Mammalia | Croatia | (34) |
| **Grippotyphosa** | **Dadas** | *Bos taurus* | Cattle | Artiodactyla | Mammalia | Türkiye | (186) |
| **Grippotyphosa** | **Dadas** | *Canis lupus* | Dog | Carnivora | Mammalia | Thailand | (142) |
| **Grippotyphosa** | **Grippotyphosa** | *Akodon arviculoides* | Montane grass mouse | Rodentia | Mammalia | Brazil | (90) |
| **Grippotyphosa** | **Grippotyphosa** | *Apodemus flavicollis* | Yellow-necked field mouse | Rodentia | Mammalia | Croatia, Slovakia | (34, 187) |
| **Grippotyphosa** | **Grippotyphosa** | *Bos taurus* | Cattle | Artiodactyla | Mammalia | Australia, Iran (Islamic Republic of), Nigeria, Tunisia, United Republic of Tanzania, United States of America | (35, 59, 188-193) |
| **Grippotyphosa** | **Grippotyphosa** | *Canis lupus* | Dog | Carnivora | Mammalia | United States of America | (93) |

| | | | | | | | |
|---|---|---|---|---|---|---|---|
| **Grippotyphosa** | **Grippotyphosa** | *Capra aegagrus* | Goat | Artiodactyla | Mammalia | Israel | (194) |
| **Grippotyphosa** | **Grippotyphosa** | *Clethrionomys glareolus* | Bank vole | Rodentia | Mammalia | Slovakia | (187) |
| **Grippotyphosa** | **Grippotyphosa** | *Dermacentor marginatus* | Tick (obtained from cattle) | Ixodidae | Arachnida | Kazakhstan | (195) |
| **Grippotyphosa** | **Grippotyphosa** | *Didelphis marsupialis* | Common opossum | Didelphimorphia | Mammalia | Brazil | (90) |
| **Grippotyphosa** | **Grippotyphosa** | *Equus caballus* | Horse | Perissodactyla | Mammalia | Austria, Germany, Italy, Luxembourg, Netherlands, Portugal, Switzerland, United Kingdom of Great Britain and Northern Ireland | (36, 196) |
| **Grippotyphosa** | **Grippotyphosa** | *Glaucomys volans* | Flying squirrel | Rodentia | Mammalia | Japan | (197) |
| **Grippotyphosa** | **Grippotyphosa** | *Hemiechinus auritus* | Long eared hedgehog | Eulipotyphla | Mammalia | Israel | (99) |
| **Grippotyphosa** | **Grippotyphosa** | *Isoodon macrourus* | Northern brown bandicoot | Peramelemorphia | Mammalia | Australia | (28) |
| **Grippotyphosa** | **Grippotyphosa** | *Mephitis mephitis* | Striped skunk | Carnivora | Mammalia | United States of America | (103) |
| **Grippotyphosa** | **Grippotyphosa** | *Microtus agrestis* | Field vole | Rodentia | Mammalia | Russian Federation | (198) |
| **Grippotyphosa** | **Grippotyphosa** | *Microtus arvalis* | Common vole | Rodentia | Mammalia | Netherlands, Poland, Slovakia | (187, 199, 200) |
| **Grippotyphosa** | **Grippotyphosa** | *Microtus oeconomus* | Tundra vole | Rodentia | Mammalia | Russian Federation | (201) |
| **Grippotyphosa** | **Grippotyphosa** | *Microtus pennsylvanicus* | Meadow vole | Rodentia | Mammalia | United States of America | (202) |
| **Grippotyphosa** | **Grippotyphosa** | *Microtus subterraneus* | Pine vole | Rodentia | Mammalia | Slovakia | (187) |
| **Grippotyphosa** | **Grippotyphosa** | *Mus musculus* | House mouse | Rodentia | Mammalia | Egypt, Iran (Islamic Republic of) | (203, 204) |
| **Grippotyphosa** | **Grippotyphosa** | *Nectomys squamipes* | South American water rat | Rodentia | Mammalia | Brazil | (90, 177) |
| **Grippotyphosa** | **Grippotyphosa** | *Oligoryzomys eliurus* | Brazilian pygmy rice rat | Rodentia | Mammalia | Brazil | (90) |
| **Grippotyphosa** | **Grippotyphosa** | *Ondatra spp.* | Muskrat | Rodentia | Mammalia | France | (205) |
| **Grippotyphosa** | **Grippotyphosa** | *Ondatra zibethicus* | Muskrat | Rodentia | Mammalia | Netherlands | (60) |
| **Grippotyphosa** | **Grippotyphosa** | *Ovis aries* | Sheep | Artiodactyla | Mammalia | Iran (Islamic Republic of), Spain | (190, 206, 207) |

| | | | | | | | |
|---|---|---|---|---|---|---|---|
| **Grippotyphosa** | **Grippotyphosa** | *Oxymycterus quaestor* | Quaestor hocicudo | Rodentia | Mammalia | Brazil | (90) |
| **Grippotyphosa** | **Grippotyphosa** | *Philander opossum* | Gray four-eyed opossum | Didelphimorphia | Mammalia | Brazil | (16) |
| **Grippotyphosa** | **Grippotyphosa** | *Procyon lotor* | Raccoon | Carnivora | Mammalia | United States of America | (18, 96, 97) |
| **Grippotyphosa** | **Grippotyphosa** | *Rattus rattus* | Black rat | Rodentia | Mammalia | Egypt, Israel | (114, 178) |
| **Grippotyphosa** | **Grippotyphosa** | *Rattus sordidus* | Canefield rat | Rodentia | Mammalia | Australia | (28) |
| **Grippotyphosa** | **Grippotyphosa** | *Reithrodontomys megalotis* | Western harvest mouse | Rodentia | Mammalia | United States of America | (96) |
| **Grippotyphosa** | **Grippotyphosa** | *Sooretamys angouya* | Paraguayan rice rat | Rodentia | Mammalia | Brazil | (90) |
| **Grippotyphosa** | **Grippotyphosa** | *Sorex spp.* | Vagrant shrew | Eulipotyphla | Mammalia | United States of America | (208) |
| **Grippotyphosa** | **Grippotyphosa** | *Sundamys muelleri* | Mueller's rat | Rodentia | Mammalia | Malaysia | (27, 71) |
| **Grippotyphosa** | **Grippotyphosa** | *Sus scrofa* | Pig | Artiodactyla | Mammalia | United States of America | (209) |
| **Grippotyphosa** | **Grippotyphosa** | *Sylvilagus aquaticus* | Swamp rabbit | Lagomorpha | Mammalia | United States of America | (210) |
| **Grippotyphosa** | **Grippotyphosa** | *Sylvilagus floridanus* | Cottontail rabbit | Lagomorpha | Mammalia | United States of America | (210) |
| **Grippotyphosa** | **Grippotyphosa** | *Tatera robusta* | Fringe-tailed gerbil | Rodentia | Mammalia | Kenya | (211) |
| **Grippotyphosa** | **Grippotyphosa** | *Thaptomys nigrita* | Blackish grass mouse | Rodentia | Mammalia | Brazil | (90) |
| **Grippotyphosa** | **Grippotyphosa** | *Urocyon cinereoargenteus* | Grey fox | Carnivora | Mammalia | United States of America | (17) |
| **Grippotyphosa** | **Grippotyphosa** | Not stated | Fruit bat | Chiroptera | Mammalia | Peru | (212) |
| **Grippotyphosa** | **Grippotyphosa** | Not stated | Opossum | Didelphimorphia | Mammalia | United States of America | (208) |
| **Grippotyphosa** | **Grippotyphosa** | Not stated | Rodent | Rodentia | Mammalia | Poland | (29) |
| **Grippotyphosa** | **Grippotyphosa** | Not stated | Rotel mice | Rodentia | Mammalia | Slovakia | (213) |
| **Grippotyphosa** | **Muelleri** | *Sundamys muelleri* | Mueller's rat | Rodentia | Mammalia | Malaysia | (71) |

| | | | | | | | |
|---|---|---|---|---|---|---|---|
| **Grippotyphosa** | **Ratnapura** | *Bos taurus* | Cattle | Artiodactyla | Mammalia | Zimbabwe | (214) |
| **Grippotyphosa** | **Ratnapura** | *Rattus rattus* | Black rat | Rodentia | Mammalia | India | (215) |
| **Grippotyphosa** | **Valbuzzi** | *Rattus rattus* | Black rat | Rodentia | Mammalia | India | (215) |
| **Grippotyphosa** | **Vanderhoedeni** | *Hemiechinus auritus* | Long eared hedgehog | Eulipotyphla | Mammalia | Israel | (216) |
| **Hebdomadis** | **Borincana** | *Ondatra spp.* | Muskrat | Rodentia | Mammalia | France | (205) |
| **Hebdomadis** | **Borincana** | *Philander spp.* | Opossum | Didelphimorphia | Mammalia | Peru | (217) |
| **Hebdomadis** | **Borincana** | *Rat spp.* | Rat | Rodentia | Mammalia | Peru | (217) |
| **Hebdomadis** | **Goiano** | *Bos taurus* | Cattle | Artiodactyla | Mammalia | Brazil | (218) |
| **Hebdomadis** | **Goiano** | *Sus scrofa* | Pig | Artiodactyla | Mammalia | Peru | (219) |
| **Hebdomadis** | **Hebdomadis** | *Apodemus flavicollis* | Yellow-necked field mouse | Rodentia | Mammalia | Romania | (220) |
| **Hebdomadis** | **Hebdomadis** | *Apodemus sylvaticus* | Common field mouse | Rodentia | Mammalia | Iran (Islamic Republic of), Romania | (221, 222) |
| **Hebdomadis** | **Hebdomadis** | *Bos taurus* | Cattle | Artiodactyla | Mammalia | Japan | (6) |
| **Hebdomadis** | **Hebdomadis** | *Microtus fortis* | Reed vole | Rodentia | Mammalia | China | (1) |
| **Hebdomadis** | **Hebdomadis** | *Mus musculus* | House mouse | Rodentia | Mammalia | Romania | (222) |
| **Hebdomadis** | **Hebdomadis** | *Mus spicilegus* | Steppe mouse | Rodentia | Mammalia | Romania | (220, 222) |
| **Hebdomadis** | **Hebdomadis** | *Procyon lotor* | Raccoon | Carnivora | Mammalia | Japan | (223) |
| **Hebdomadis** | **Hebdomadis** | *Rattus norvegicus* | Norwegian rat | Rodentia | Mammalia | China | (1) |
| **Hebdomadis** | **Hebdomadis** | *Rattus spp.* | Rat | Rodentia | Mammalia | China | (26) |
| **Hebdomadis** | **Iassy** | *Mus musculus* | House mouse | Rodentia | Mammalia | Romania | (224) |
| **Hebdomadis** | **Jules** | *Sus scrofa* | Pig | Artiodactyla | Mammalia | Peru | (138) |

| | | | | | | | |
|---|---|---|---|---|---|---|---|
| **Hebdomadis** | **Kremastos** | *Bos taurus* | Cattle | Artiodactyla | Mammalia | Japan, Peru | (135, 225) |
| **Hebdomadis** | **Kremastos** | *Isoodon macrourus* | Northern brown bandicoot | Peramelemorphia | Mammalia | Australia | (14) |
| **Hebdomadis** | **Kremastos** | *Perameles nasuta* | Long-nosed bandicoot | Peramelemorphia | Mammalia | Australia | (14) |
| **Hebdomadis** | **Marondera** | *Bos taurus* | Cattle | Artiodactyla | Mammalia | Zimbabwe | (50, 226) |
| **Hebdomadis** | **Maru** | *Herpestes urva* | Crab-eating mongoose | Carnivora | Mammalia | China | (227) |
| **Hebdomadis** | **Maru** | *Proechimys semispinosus* | Spiny rat | Rodentia | Mammalia | Panama | (137) |
| **Hebdomadis** | **Mhou** | *Bos taurus* | Cattle | Artiodactyla | Mammalia | Zimbabwe | (50, 226) |
| **Hurstbridge** | **Hurstbridge** | *Sus scrofa* | Pig | Artiodactyla | Mammalia | Australia | (228) |
| **Icterohaemorrhagiae** | **Bogvere** | *Rattus rattus* | Black rat | Rodentia | Mammalia | Guadeloupe | (229) |
| **Icterohaemorrhagiae** | **Copenhageni** | *Bos taurus* | Cattle | Artiodactyla | Mammalia | Brazil, New Zealand | (92, 150, 172, 230, 231) |
| **Icterohaemorrhagiae** | **Copenhageni** | *Canis lupus* | Dog | Carnivora | Mammalia | Barbados, Brazil, Italy, Trinidad and Tobago | (8, 72, 76, 78, 153, 168, 172, 230-234) |
| **Icterohaemorrhagiae** | **Copenhageni** | *Cebus capucinus* | Capuchin monkey | Primates | Mammalia | Colombia | (235) |
| **Icterohaemorrhagiae** | **Copenhageni** | *Herpestes auropunctatus* | Lesser Indian mongoose | Carnivora | Mammalia | Grenada | (66) |
| **Icterohaemorrhagiae** | **Copenhageni** | *Mus musculus* | House mouse | Rodentia | Mammalia | Barbados | (81) |
| **Icterohaemorrhagiae** | **Copenhageni** | *Myocastor coypus* | Nutria | Rodentia | Mammalia | Netherlands | (236) |
| **Icterohaemorrhagiae** | **Copenhageni** | *Ondatra spp.* | Muskrat | Rodentia | Mammalia | France | (205) |
| **Icterohaemorrhagiae** | **Copenhageni** | *Ondatra zibethicus* | Muskrat | Rodentia | Mammalia | Netherlands | (60) |
| **Icterohaemorrhagiae** | **Copenhageni** | *Proechimys guyannensis* | Guyenne spiny rat | Rodentia | Mammalia | Trinidad and Tobago | (66, 72) |
| **Icterohaemorrhagiae** | **Copenhageni** | *Rat spp.* | Rat | Rodentia | Mammalia | Brazil | (172) |
| **Icterohaemorrhagiae** | **Copenhageni** | *Rattus norvegicus* | Norwegian rat | Rodentia | Mammalia | Barbados, Brazil, Colombia, Germany, Grenada, Japan, New Zealand, Peru, Russian Federation, Trinidad and Tobago, United States of America | (66, 72, 76, 79, 81, 113, 219, 230, 231, 235, 237-244) |

| | | | | | | | |
|---|---|---|---|---|---|---|---|
| **Icterohaemorrhagiae** | **Copenhageni** | *Rattus rattus* | Black rat | Rodentia | Mammalia | Barbados, Brazil, French Guiana, Grenada, Trinidad and Tobago | (66, 72, 76, 79, 81, 230, 231, 245) |
| **Icterohaemorrhagiae** | **Copenhageni** | *Rattus spp.* | Rat | Rodentia | Mammalia | Romania, Trinidad and Tobago | (233, 246) |
| **Icterohaemorrhagiae** | **Copenhageni** | *Saimiri sciureus* | Squirrel monkey | Primates | Mammalia | French Guiana | (245) |
| **Icterohaemorrhagiae** | **Hongchon** | *Apodemus agrarius* | Striped field mouse | Rodentia | Mammalia | Republic of Korea | (247) |
| **Icterohaemorrhagiae** | **Icterohaemorrhagiae** | *Apodemus agrarius* | Striped field mouse | Rodentia | Mammalia | China | (1) |
| **Icterohaemorrhagiae** | **Icterohaemorrhagiae** | *Bos taurus* | Cattle | Artiodactyla | Mammalia | Brazil, India, Nigeria, United Kingdom of Great Britain and Northern Ireland, United States of America | (35, 248-251) |
| **Icterohaemorrhagiae** | **Icterohaemorrhagiae** | *Calomys lauchas* | Small vesper mouse | Rodentia | Mammalia | Argentina | (146) |
| **Icterohaemorrhagiae** | **Icterohaemorrhagiae** | *Canis lupus* | Dog | Carnivora | Mammalia | Italy, Netherlands, Puerto Rico, United States of America | (8, 115, 158, 167, 170, 252) |
| **Icterohaemorrhagiae** | **Icterohaemorrhagiae** | *Capra aegagrus* | Goat | Artiodactyla | Mammalia | Spain | (207) |
| **Icterohaemorrhagiae** | **Icterohaemorrhagiae** | *Cricetinae spp.* | Hamster | Rodentia | Mammalia | United States of America | (252) |
| **Icterohaemorrhagiae** | **Icterohaemorrhagiae** | *Cricetomys spp.* | Giant pouched rat | Rodentia | Mammalia | United Republic of Tanzania | (59) |
| **Icterohaemorrhagiae** | **Icterohaemorrhagiae** | *Didelphis marsupialis* | Common opossum | Didelphimorphia | Mammalia | Brazil | (90) |
| **Icterohaemorrhagiae** | **Icterohaemorrhagiae** | *Erinaceus europaeus* | Hedgehog | Eulipotyphla | Mammalia | Italy, Portugal | (11, 87) |
| **Icterohaemorrhagiae** | **Icterohaemorrhagiae** | *Herpestes auropunctatus* | Lesser Indian mongoose | Carnivora | Mammalia | Guadeloupe, Puerto Rico, United States of America | (13, 24, 106, 115) |
| **Icterohaemorrhagiae** | **Icterohaemorrhagiae** | *Herpestes ichneumon* | Egyption mongoose | Carnivora | Mammalia | Egypt | (204) |
| **Icterohaemorrhagiae** | **Icterohaemorrhagiae** | *Mephitis mephitis* | Skunk | Carnivora | Mammalia | United States of America | (253) |
| **Icterohaemorrhagiae** | **Icterohaemorrhagiae** | *Mephitis mephitis* | Striped skunk | Carnivora | Mammalia | United States of America | (102, 103) |
| **Icterohaemorrhagiae** | **Icterohaemorrhagiae** | *Microtus montebelli* | Japanese field vole | Rodentia | Mammalia | Japan | (69) |
| **Icterohaemorrhagiae** | **Icterohaemorrhagiae** | *Mus musculus* | House mouse | Rodentia | Mammalia | Puerto Rico, United States of America | (24, 106, 115, 253) |
| **Icterohaemorrhagiae** | **Icterohaemorrhagiae** | *Myocastor coypus* | Nutria | Rodentia | Mammalia | France | (254) |

| | | | | | | | |
|---|---|---|---|---|---|---|---|
| **Icterohaemorrhagiae** | **Icterohaemorrhagiae** | *Ondatra spp.* | Muskrat | Rodentia | Mammalia | France | (205) |
| **Icterohaemorrhagiae** | **Icterohaemorrhagiae** | *Ovis aries* | Sheep | Artiodactyla | Mammalia | Spain | (207) |
| **Icterohaemorrhagiae** | **Icterohaemorrhagiae** | *Paguma larvata* | Masked palm civet | Carnivora | Mammalia | China | (80) |
| **Icterohaemorrhagiae** | **Icterohaemorrhagiae** | *Procyon lotor* | Raccoon | Carnivora | Mammalia | United States of America | (253) |
| **Icterohaemorrhagiae** | **Icterohaemorrhagiae** | *Proechimys spp.* | Spiny rat | Rodentia | Mammalia | Peru | (255) |
| **Icterohaemorrhagiae** | **Icterohaemorrhagiae** | *Rattus exulans* | Polynesian rat | Rodentia | Mammalia | United States of America | (24, 106) |
| **Icterohaemorrhagiae** | **Icterohaemorrhagiae** | *Rattus norvegicus* | Norwegian rat | Rodentia | Mammalia | Algeria, Canada, Colombia, French Guiana, Israel, Japan, Malaysia, Philippines, Portugal, Puerto Rico, South Africa, Spain, Trinidad and Tobago, Tunisia, United Kingdom of Great Britain and Northern Ireland, United States of America | (24, 27, 72, 84, 87, 88, 106, 114, 115, 140, 237, 256-268) |
| **Icterohaemorrhagiae** | **Icterohaemorrhagiae** | *Rattus rattus* | Black rat | Rodentia | Mammalia | China, Colombia, Portugal, Puerto Rico, Trinidad and Tobago, Tunisia, United States of America | (24, 72, 80, 84, 87, 106, 115, 260, 269, 270) |
| **Icterohaemorrhagiae** | **Icterohaemorrhagiae** | *Rattus spp.* | Rat | Rodentia | Mammalia | Peru, Puerto Rico, Romania, Trinidad and Tobago, United States of America | (115, 165, 233, 246, 271, 272) |
| **Icterohaemorrhagiae** | **Icterohaemorrhagiae** | *Rattus tanezumi* | Oriental house rat | Rodentia | Mammalia | Philippines | (140) |
| **Icterohaemorrhagiae** | **Icterohaemorrhagiae** | *Sundamys muelleri* | Mueller's rat | Rodentia | Mammalia | Malaysia | (27) |
| **Icterohaemorrhagiae** | **Icterohaemorrhagiae** | *Sus scrofa* | Pig | Artiodactyla | Mammalia | Brazil, India, United Kingdom of Great Britain and Northern Ireland, United States of America | (248, 273-275) |
| **Icterohaemorrhagiae** | **Icterohaemorrhagiae** | *Urocyon cinereoargenteus* | Gray fox | Carnivora | Mammalia | United States of America | (253) |
| **Icterohaemorrhagiae** | **Icterohaemorrhagiae** | *Vulpes vulpes* | Red fox | Carnivora | Mammalia | Egypt | (204) |
| **Icterohaemorrhagiae** | **Icterohaemorrhagiae** | Not stated | Fruit bat | Chiroptera | Mammalia | Peru | (212) |
| **Icterohaemorrhagiae** | **Icterohaemorrhagiae** | Not stated | Opossum | Didelphimorphia | Mammalia | United States of America | (120) |
| **Icterohaemorrhagiae** | **Icterohaemorrhagiae** | Not stated | Rodent | Rodentia | Mammalia | Poland | (29) |
| **Icterohaemorrhagiae** | **Lai** | *Apodemus agrarius* | Striped field mouse | Rodentia | Mammalia | Republic of Korea | (276, 277) |

| | | | | | | | |
|---|---|---|---|---|---|---|---|
| **Icterohaemorrhagiae** | **Lai** | *Rat spp.* | Rat | Rodentia | Mammalia | Republic of Korea | (278) |
| **Icterohaemorrhagiae** | **Lai** | *Rattus tanezumi* | Oriental house rat | Rodentia | Mammalia | China | (279) |
| **Icterohaemorrhagiae** | **Mankarso** | *Maxomys whiteheadi* | Whitehead's spiny rat | Rodentia | Mammalia | Indonesia | (71) |
| **Icterohaemorrhagiae** | **Mankarso** | *Rattus spp.* | Rat | Rodentia | Mammalia | Trinidad and Tobago | (233) |
| **Icterohaemorrhagiae** | **Sokoine** | *Bos taurus* | Cattle | Artiodactyla | Mammalia | United Republic of Tanzania | (59, 280) |
| **Icterohaemorrhagiae** | **Sokoine** | *Cricetomys spp.* | Giant pouched rat | Rodentia | Mammalia | United Republic of Tanzania | (59) |
| **Icterohaemorrhagiae** | **Yeonchon** | *Rat spp.* | Rat | Rodentia | Mammalia | Republic of Korea | (278) |
| **Icterohaemorrhagiae** | **Zimbabwe** | *Bos taurus* | Cattle | Artiodactyla | Mammalia | Zimbabwe | (50, 281) |
| **Iquitos** | **Varillal** | *Rattus norvegicus* | Norwegian rat | Rodentia | Mammalia | Peru | (272) |
| **Iquitos** | **Varillal** | *Rattus rattus* | Black rat | Rodentia | Mammalia | Peru | (272) |
| **Javanica** | **Ceylonica** | *Suncus caeruleus* | Musk shrew | Eulipotyphla | Mammalia | Sri Lanka | (282) |
| **Javanica** | **Izatnagar** | *Rattus rattus* | Black rat | Rodentia | Mammalia | India | (283, 284) |
| **Javanica** | **Javanica** | *Bos taurus* | Cattle | Artiodactyla | Mammalia | Malaysia | (3-5) |
| **Javanica** | **Javanica** | *Canis lupus* | Dog | Carnivora | Mammalia | Sri Lanka | (180) |
| **Javanica** | **Javanica** | *Felis catus* | Cat | Carnivora | Mammalia | Indonesia | (127) |
| **Javanica** | **Javanica** | *Mus musculus* | House mouse | Rodentia | Mammalia | India | (285, 286) |
| **Javanica** | **Javanica** | *Myocastor coypus* | Nutria | Rodentia | Mammalia | China | (15) |
| **Javanica** | **Javanica** | *Ovis aries* | Sheep | Artiodactyla | Mammalia | India | (285, 287) |
| **Javanica** | **Javanica** | *Rat spp.* | Rat | Rodentia | Mammalia | Sri Lanka, Thailand | (22, 180) |
| **Javanica** | **Javanica** | *Rattus argentiventer* | Rice-field rat | Rodentia | Mammalia | Malaysia | (27) |

| | | | | | | | |
|---|---|---|---|---|---|---|---|
| **Javanica** | **Javanica** | *Rattus exulans* | Polynesian rat | Rodentia | Mammalia | Malaysia | (129, 130) |
| **Javanica** | **Javanica** | *Rattus norvegicus* | Norwegian rat | Rodentia | Mammalia | India, Malaysia, Philippines | (27, 130, 132, 140, 285, 286, 288) |
| **Javanica** | **Javanica** | *Rattus rattus* | Black rat | Rodentia | Mammalia | India, Japan, Malaysia, Thailand | (74, 75, 129, 130, 132, 289-291) |
| **Javanica** | **Javanica** | *Rattus rattus* | Malayan black rat | Rodentia | Mammalia | Malaysia | (27) |
| **Javanica** | **Javanica** | *Rattus spp.* | Rat | Rodentia | Mammalia | China, Philippines, Thailand | (15, 26, 133, 134) |
| **Javanica** | **Javanica** | *Rattus tanezumi* | Oriental house rat | Rodentia | Mammalia | Philippines | (140) |
| **Javanica** | **Menoni** | *Bandicota bengalensis* | Lesser bandicoot rat | Rodentia | Mammalia | India | (292) |
| **Javanica** | **Sorexjalna** | *Erinaceus europaeus* | Hedgehog | Eulipotyphla | Mammalia | Netherlands | (41) |
| **Javanica** | **Sorexjalna** | Not stated | Rodent | Rodentia | Mammalia | Poland | (29) |
| **Javanica** | **Zhenkang** | *Rattus flavipectus* | Yellow breasted rat | Rodentia | Mammalia | China | (293) |
| **Louisiana** | **Lanka** | *Marmosa fuscata* | Mouse opossum | Didelphimorphia | Mammalia | Trinidad and Tobago | (66, 72) |
| **Louisiana** | **Lanka** | *Marmosa mitis* | Pouchless opossum | Didelphimorphia | Mammalia | Trinidad and Tobago | (66, 72) |
| **Louisiana** | **Lanka** | *Rattus rattus* | Black rat | Rodentia | Mammalia | Trinidad and Tobago | (72) |
| **Louisiana** | **Louisiana** | *Dasypus novemcinctus* | Long nosed armadillo | Cingulata | Mammalia | United States of America | (294) |
| **Louisiana** | **Orleans** | *Myocastor coypus* | Nutria | Rodentia | Mammalia | United States of America | (294) |
| **Mini** | **Beye** | *Canis lupus* | Dog | Carnivora | Mammalia | Colombia | (235) |
| **Mini** | **Beye** | *Proechimys semispinosus* | Spiny rat | Rodentia | Mammalia | Panama | (65) |
| **Mini** | **Georgia** | *Bos taurus* | Cattle | Artiodactyla | Mammalia | Peru | (135) |
| **Mini** | **Georgia** | *Canis lupus* | Dog | Carnivora | Mammalia | Trinidad and Tobago | (72, 168) |
| **Mini** | **Georgia** | *Didelphis marsupialis* | Common opossum | Didelphimorphia | Mammalia | United States of America | (295) |

| | | | | | | | |
|---|---|---|---|---|---|---|---|
| **Mini** | **Georgia** | *Mephitis mephitis* | Striped skunk | Carnivora | Mam malia | United States of America | (103, 295) |
| **Mini** | **Georgia** | *Procyon lotor* | Raccoon | Carnivora | Mam malia | United States of America | (295) |
| **Mini** | **Mini** | *Arvicola terrestris* | European water vole | Rodentia | Mam malia | Bulgaria | (126) |
| **Mini** | **Mini** | *Isoodon macrourus* | Northern brown bandicoot | Peramele morphia | Mam malia | Australia | (14) |
| **Mini** | **Ruparupae** | *Didelphis marsupialis* | Common opossum | Didelphi morphia | Mam malia | Peru | (145) |
| **Mini** | **Ruparupae** | *Philander opossum* | Gray four-eyed opossum | Didelphi morphia | Mam malia | Peru | (145) |
| **Mini** | **Szwajizak** | *Bos taurus* | Cattle | Artiodact yla | Mam malia | United States of America | (296) |
| **Mini** | **Szwajizak** | *Didelphis marsupialis* | Common opossum | Didelphi morphia | Mam malia | Brazil | (90) |
| **Mini** | **Szwajizak** | *Erinaceus europaeus* | Hedgehog | Eulipotyp hla | Mam malia | Israel | (12, 99, 114) |
| **Mini** | **Szwajizak** | *Mus musculus* | House mouse | Rodentia | Mam malia | Israel | (12, 114) |
| **Mini** | **Szwajizak** | *Rattus rattus* | Black rat | Rodentia | Mam malia | Israel | (12, 88, 114) |
| **Panama** | **Cristobali** | *Didelphis spp.* | Didelpis opossum | Didelphi morphia | Mam malia | Panama | (65) |
| **Panama** | **Mangus** | *Didelphis albiventris* | White eared opossum | Didelphi morphia | Mam malia | Brazil | (16) |
| **Panama** | **Mangus** | *Herpestes auropunctatus* | Lesser Indian mongoose | Carnivora | Mam malia | Trinidad and Tobago | (72, 297) |
| **Panama** | **Panama** | *Didelphis marsupialis* | Common opossum | Didelphi morphia | Mam malia | Panama | (137) |
| **Pomona** | **Altodouro** | *Mus musculus* | House mouse | Rodentia | Mam malia | Portugal | (298) |
| **Pomona** | **Kunming** | *Apodemus chevrieri* | Chevrier's field mouse | Rodentia | Mam malia | China | (299) |
| **Pomona** | **Monjakov** | *Rattus norvegicus* | Norwegian rat | Rodentia | Mam malia | Russian Federation | (300) |
| **Pomona** | **Monjakov** | *Sus scrofa* | Pig | Artiodact yla | Mam malia | Republic of Moldova, Russian Federation, Sri Lanka, Viet Nam | (180, 300) |
| **Pomona** | **Mozdok** | *Apodemus agrarius* | Striped field mouse | Rodentia | Mam malia | Croatia, Germany, Russian Federation | (34, 51, 300, 301) |

| | | | | | | | |
|---|---|---|---|---|---|---|---|
| **Pomona** | **Mozdok** | *Apodemus flavicollis* | Yellow-necked field mouse | Rodentia | Mammalia | Croatia | (34) |
| **Pomona** | **Mozdok** | *Apodemus sylvaticus* | Common field mouse | Rodentia | Mammalia | Croatia, Russian Federation | (34, 300) |
| **Pomona** | **Mozdok** | *Bos taurus* | Cattle | Artiodactyla | Mammalia | Germany, Russian Federation, United Kingdom of Great Britain and Northern Ireland, Zimbabwe | (50, 300, 302, 303) |
| **Pomona** | **Mozdok** | *Canis lupus* | Dog | Carnivora | Mammalia | Italy | (8) |
| **Pomona** | **Mozdok** | *Crocidura russula* | Greater white toothed shrew | Eulipotyphla | Mammalia | Portugal | (87) |
| **Pomona** | **Mozdok** | *Herpestes urva* | Crab-eating mongoose | Carnivora | Mammalia | China | (227) |
| **Pomona** | **Mozdok** | *Microtus agrestis* | Field vole | Rodentia | Mammalia | United Kingdom of Great Britain and Northern Ireland | (303) |
| **Pomona** | **Mozdok** | *Microtus majori* | Major's pine vole | Rodentia | Mammalia | Russian Federation | (300) |
| **Pomona** | **Mozdok** | *Mus musculus* | House mouse | Rodentia | Mammalia | Germany | (301) |
| **Pomona** | **Mozdok** | *Mus spretus* | Algerian mouse | Rodentia | Mammalia | Portugal | (87) |
| **Pomona** | **Mozdok** | *Muscardinus avellanarius* | Hazel dormouse | Rodentia | Mammalia | Croatia | (51) |
| **Pomona** | **Mozdok** | *Sus scrofa* | Pig | Artiodactyla | Mammalia | Italy, United Kingdom of Great Britain and Northern Ireland | (304, 305) |
| **Pomona** | **Pomona** | *Akodon arviculoides* | Montane grass mouse | Rodentia | Mammalia | Brazil | (16) |
| **Pomona** | **Pomona** | *Apodemus agrarius* | Striped field mouse | Rodentia | Mammalia | Russian Federation | (306) |
| **Pomona** | **Pomona** | *Arvicola terrestris* | European water vole | Rodentia | Mammalia | Bulgaria | (126) |
| **Pomona** | **Pomona** | *Bos taurus* | Cattle | Artiodactyla | Mammalia | Argentina, Brazil, Canada, Malaysia, Nigeria, Romania, Russian Federation, South Africa, United States of America | (3, 4, 35, 147, 148, 189, 192, 300, 307-316) |
| **Pomona** | **Pomona** | *Canis lupus* | Dog | Carnivora | Mammalia | Brazil, Malaysia, United States of America | (153, 173, 317) |
| **Pomona** | **Pomona** | *Capra aegagrus* | Goat | Artiodactyla | Mammalia | Spain | (207) |
| **Pomona** | **Pomona** | *Cavia pamparum* | Pampas cavy | Rodentia | Mammalia | Argentina | (318) |
| **Pomona** | **Pomona** | *Cavia porcellus* | Guinea pig | Rodentia | Mammalia | Peru | (135) |

| | | | | | | | |
|---|---|---|---|---|---|---|---|
| **Pomona** | **Pomona** | *Didelphis albiventris* | White eared opossum | Didelphimorphia | Mammalia | Brazil | (16) |
| **Pomona** | **Pomona** | *Didelphis marsupialis* | Common opossum | Didelphimorphia | Mammalia | United States of America | (101) |
| **Pomona** | **Pomona** | *Equus caballus* | Horse | Perissodactyla | Mammalia | United Kingdom of Great Britain and Northern Ireland, United States of America | (38, 97, 303, 319, 320) |
| **Pomona** | **Pomona** | *Felis catus* | Cat | Carnivora | Mammalia | Malaysia, United States of America | (27, 101, 321) |
| **Pomona** | **Pomona** | *Felis rufa* | Wild cat | Carnivora | Mammalia | United States of America | (18, 95) |
| **Pomona** | **Pomona** | *Marmota monax* | Ground hog | Rodentia | Mammalia | Canada | (261, 322) |
| **Pomona** | **Pomona** | *Mephitis mephitis* | Striped skunk | Carnivora | Mammalia | United States of America | (18, 95, 97, 101, 103, 202) |
| **Pomona** | **Pomona** | *Mephitis spp.* | Skunk | Carnivora | Mammalia | Canada | (316, 322) |
| **Pomona** | **Pomona** | *Mirounga angustirostris* | Northern elephant seal | Carnivora | Mammalia | United States of America | (323) |
| **Pomona** | **Pomona** | *Mus musculus* | House mouse | Rodentia | Mammalia | Brazil | (16) |
| **Pomona** | **Pomona** | *Odocoileus virginianus* | White tailed deer | Artiodactyla | Mammalia | Canada, United States of America | (324-326) |
| **Pomona** | **Pomona** | *Oligoryzomys nigripes* | Black footed pygmy rice rat | Rodentia | Mammalia | Brazil | (16) |
| **Pomona** | **Pomona** | *Ovis aries* | Sheep | Artiodactyla | Mammalia | Canada, Spain, United Kingdom of Great Britain and Northern Ireland | (43, 207, 303, 316) |
| **Pomona** | **Pomona** | *Paradoxurus hermaphroditus* | Asian palm civet | Carnivora | Mammalia | Malaysia | (27) |
| **Pomona** | **Pomona** | *Procyon lotor* | Raccoon | Carnivora | Mammalia | United States of America | (18, 97) |
| **Pomona** | **Pomona** | *Rattus norvegicus* | Norwegian rat | Rodentia | Mammalia | Colombia | (268) |
| **Pomona** | **Pomona** | *Rattus rattus* | Black rat | Rodentia | Mammalia | Brazil | (16) |
| **Pomona** | **Pomona** | *Sus scrofa* | Pig | Artiodactyla | Mammalia | Brazil, Canada, Colombia, Hungary, Italy, Malaysia, New Zealand, Peru, Sri Lanka, United States of America | (130, 172, 173, 180, 219, 262, 270, 305, 316, 327-331) |
| **Pomona** | **Pomona** | *Vulpes spp.* | Fox | Carnivora | Mammalia | Canada | (316) |
| **Pomona** | **Pomona** | *Zalophus californianus* | California sea lion | Carnivora | Mammalia | United States of America | (332, 333) |

| | | | | | | | |
|---|---|---|---|---|---|---|---|
| **Pomona** | **Proechimys** | *Philander spp.* | Opossum | Didelphimorphia | Mammalia | Peru | (217) |
| **Pomona** | **Proechimys** | *Proechimys semispinosus* | Spiny rat | Rodentia | Mammalia | Panama | (65) |
| **Pomona** | **Proechimys** | *Rat spp.* | Rat | Rodentia | Mammalia | Peru | (217) |
| **Pomona** | **Tropica** | *Liomys adspersus* | Spiny pocket mouse | Rodentia | Mammalia | Panama | (137) |
| **Pomona** | **Tropica** | *Mus musculus* | House mouse | Rodentia | Mammalia | Brazil | (16, 107, 108) |
| **Pomona** | **Tropica** | *Proechimys semispinosus* | Spiny rat | Rodentia | Mammalia | Panama | (137) |
| **Pomona** | **Tsaratsovo** | *Apodemus agrarius* | Striped field mouse | Rodentia | Mammalia | Croatia | (53) |
| **Pomona** | **Tsaratsovo** | *Apodemus flavicollis* | Yellow-necked field mouse | Rodentia | Mammalia | Croatia | (53) |
| **Pyrogenes** | **Abramis** | *Philander opossum* | Gray four-eyed opossum | Didelphimorphia | Mammalia | Nicaragua | (174) |
| **Pyrogenes** | **Guaratuba** | *Philander opossum* | Gray four-eyed opossum | Didelphimorphia | Mammalia | Brazil | (90) |
| **Pyrogenes** | **Kwale** | *Bos taurus* | Cattle | Artiodactyla | Mammalia | Zimbabwe | (50, 334) |
| **Pyrogenes** | **Manilae** | *Rat spp.* | Rat | Rodentia | Mammalia | Philippines | (139) |
| **Pyrogenes** | **Manilae** | *Rattus norvegicus* | Norwegian rat | Rodentia | Mammalia | Philippines | (140, 335) |
| **Pyrogenes** | **Manilae** | *Rattus rattus* | Black rat | Rodentia | Mammalia | Philippines | (335) |
| **Pyrogenes** | **Manilae** | *Rattus spp.* | Rat | Rodentia | Mammalia | Philippines | (133) |
| **Pyrogenes** | **Manilae** | *Rattus tanezumi* | Oriental house rat | Rodentia | Mammalia | Philippines | (140) |
| **Pyrogenes** | **Mombe** | *Bos taurus* | Cattle | Artiodactyla | Mammalia | Zimbabwe | (50, 334) |
| **Pyrogenes** | **Myocastoris** | *Myocastor coypus* | Nutria | Rodentia | Mammalia | United States of America | (336) |
| **Pyrogenes** | **Nigeria** | *Bos taurus* | Cattle | Artiodactyla | Mammalia | Nigeria, Zimbabwe | (50, 334, 337) |
| **Pyrogenes** | **Pyrogenes** | *Bos taurus* | Cattle | Artiodactyla | Mammalia | Nigeria | (2) |

| | | | | | | | |
|---|---|---|---|---|---|---|---|
| **Pyrogenes** | **Pyrogenes** | *Nectomys squamipes* | South American water rat | Rodentia | Mammalia | Brazil | (177) |
| **Pyrogenes** | **Pyrogenes** | *Rat spp.* | Field rat | Rodentia | Mammalia | Thailand | (20, 21) |
| **Pyrogenes** | **Pyrogenes** | *Rat spp.* | Rat | Rodentia | Mammalia | Thailand | (22) |
| **Pyrogenes** | **Pyrogenes** | *Rattus rattus* | Black rat | Rodentia | Mammalia | Egypt | (178) |
| **Pyrogenes** | **Pyrogenes** | *Rattus spp.* | Rat | Rodentia | Mammalia | Philippines, Thailand | (133, 134) |
| **Pyrogenes** | **Pyrogenes** | *Sus scrofa* | Pig | Artiodactyla | Mammalia | Peru, Philippines | (135, 338, 339) |
| **Pyrogenes** | **Robinsoni** | *Rattus sordidus* | Canefield rat | Rodentia | Mammalia | Australia | (14) |
| **Pyrogenes** | **Robinsoni** | *Uromys caudimaculatus* | Giant white-tailed rat | Rodentia | Mammalia | Australia | (14) |
| **Pyrogenes** | **Varela** | Not stated | Opossum | Didelphimorphia | Mammalia | Nicaragua | (65) |
| **Pyrogenes** | **Zanoni** | *Bos taurus* | Cattle | Artiodactyla | Mammalia | Australia | (340) |
| **Pyrogenes** | **Zanoni** | *Isoodon macrourus* | Northern brown bandicoot | Peramelemorphia | Mammalia | Australia | (14) |
| **Pyrogenes** | **Zanoni** | *Melomys cervinipes* | Fawn-footed mosaic-tailed rat | Rodentia | Mammalia | Australia | (28) |
| **Pyrogenes** | **Zanoni** | *Melomys lutillus* | Papua grassland mosaic-tailed rat | Rodentia | Mammalia | Australia | (14) |
| **Pyrogenes** | **Zanoni** | *Mus musculus* | House mouse | Rodentia | Mammalia | Australia | (14) |
| **Pyrogenes** | **Zanoni** | *Rattus fuscipes* | Bush rat | Rodentia | Mammalia | Australia | (14) |
| **Pyrogenes** | **Zanoni** | *Rattus norvegicus* | Norwegian rat | Rodentia | Mammalia | Australia | (14) |
| **Pyrogenes** | **Zanoni** | *Rattus rattus* | Black rat | Rodentia | Mammalia | Australia | (14) |
| **Pyrogenes** | **Zanoni** | *Rattus sordidus* | Canefield rat | Rodentia | Mammalia | Australia | (14) |
| **Pyrogenes** | **Zanoni** | *Uromys caudimaculatus* | Giant white-tailed rat | Rodentia | Mammalia | Australia | (28) |
| **Pyrogenes** | **Zanoni** | Not stated | Rodent | Rodentia | Mammalia | Australia | (30) |

| | | | | | | | |
|---|---|---|---|---|---|---|---|
| **Ranarum** | **Pinchang** | *Rana nigromaculata* | Black spotted frog | Anura | Amphibia | China | (341) |
| **Ranarum** | **Ranarum** | *Equus caballus* | Horse | Perissodactyla | Mammalia | Brazil | (342) |
| **Ranarum** | **Ranarum** | *Rana pipiens* | Northern leopard frog | Anura | Amphibia | United States of America | (343) |
| **Sarmin** | **Machiguenga** | *Philander opossum* | Gray four-eyed opossum | Didelphimorphia | Mammalia | Peru | (145) |
| **Sarmin** | **Rio** | *Rattus rattus* | Black rat | Rodentia | Mammalia | Brazil | (344) |
| **Sarmin** | **Weaveri** | *Proechimys semispinosus* | Spiny rat | Rodentia | Mammalia | Panama | (137) |
| **Sejroe** | **Balcanica** | *Bos taurus* | Cattle | Artiodactyla | Mammalia | United States of America, Zimbabwe | (50, 314) |
| **Sejroe** | **Balcanica** | *Capra aegagrus* | Goat | Artiodactyla | Mammalia | New Zealand | (329) |
| **Sejroe** | **Balcanica** | *Trichosurus vulpecula* | Brushtailed possum | Diprotodontia | Mammalia | Australia, New Zealand | (100, 111, 329, 345, 346) |
| **Sejroe** | **Caribe** | *Rattus norvegicus* | Norwegian rat | Rodentia | Mammalia | Trinidad and Tobago | (72, 297) |
| **Sejroe** | **Gorgas** | *Proechimys semispinosus* | Spiny rat | Rodentia | Mammalia | Panama | (65) |
| **Sejroe** | **Guaricura** | *Bos taurus* | Cattle | Artiodactyla | Mammalia | Brazil | (218) |
| **Sejroe** | **Guaricura** | *Bubalus bubalis* | Domestic water buffalo | Artiodactyla | Mammalia | Brazil | (347) |
| **Sejroe** | **Hardjo Subtype Bovis** | *Bos taurus* | Cattle | Artiodactyla | Mammalia | Argentina, Australia, Canada, Netherlands, United Kingdom of Great Britain and Northern Ireland, United Republic of Tanzania, United States of America, Zimbabwe | (50, 189, 200, 340, 348-354) |
| **Sejroe** | **Hardjo Subtype Not Determined** | *Bos taurus* | Cattle | Artiodactyla | Mammalia | Argentina, Australia, Brazil, Canada, Colombia, Germany, Ireland, Malaysia, New Zealand, Nigeria, Portugal, Puerto Rico, South Africa, United Kingdom of Great Britain and Northern Ireland, United States of America | (3, 4, 35, 192, 193, 248, 250, 313-315, 329, 355-375) |
| **Sejroe** | **Hardjo Subtype Not Determined** | *Chaetophractus villosus* | Big hairy armadillo | Cingulata | Mammalia | Argentina | (123) |
| **Sejroe** | **Hardjo Subtype Not Determined** | *Equus caballus* | Horse | Perissodactyla | Mammalia | Argentina, United Kingdom of Great Britain and Northern Ireland | (37, 376) |
| **Sejroe** | **Hardjo Subtype Not Determined** | *Mus spicilegus* | Steppe mouse | Rodentia | Mammalia | Romania | (220) |

| | | | | | | | |
|---|---|---|---|---|---|---|---|
| **Sejroe** | **Hardjo Subtype Not Determined** | *Myocastor coypus* | Nutria | Rodentia | Mammalia | United Kingdom of Great Britain and Northern Ireland | (377) |
| **Sejroe** | **Hardjo Subtype Not Determined** | *Ovis aries* | Sheep | Artiodactyla | Mammalia | Australia, New Zealand, United Kingdom of Great Britain and Northern Ireland | (329, 361, 378, 379) |
| **Sejroe** | **Hardjo Subtype Not Determined** | *Procyon lotor* | Raccoon | Carnivora | Mammalia | United States of America | (97) |
| **Sejroe** | **Hardjo Subtype Not Determined** | *Sus scrofa* | Pig | Artiodactyla | Mammalia | Ireland, United Kingdom of Great Britain and Northern Ireland | (44, 329, 361, 362, 380) |
| **Sejroe** | **Hardjo Subtype Prajitno** | *Apodemus sylvaticus* | Common field mouse | Rodentia | Mammalia | Romania | (220) |
| **Sejroe** | **Hardjo Subtype Prajitno** | *Bos taurus* | Cattle | Artiodactyla | Mammalia | Canada, United Kingdom of Great Britain and Northern Ireland | (349-352, 381) |
| **Sejroe** | **Istrica** | *Mus musculus* | House mouse | Rodentia | Mammalia | Croatia | (53) |
| **Sejroe** | **Medanensis** | *Isoodon macrourus* | Northern brown bandicoot | Peramelemorphia | Mammalia | Australia | (28) |
| **Sejroe** | **Medanensis** | *Perameles nasuta* | Long-nosed bandicoot | Peramelemorphia | Mammalia | Australia | (28) |
| **Sejroe** | **Polonica** | *Erinaceus europaeus* | European hedgehog | Eulipotyphla | Mammalia | Poland | (382) |
| **Sejroe** | **Polonica** | *Mus musculus* | House mouse | Rodentia | Mammalia | Romania | (224) |
| **Sejroe** | **Polonica** | *Rattus rattus* | Black rat | Rodentia | Mammalia | Egypt | (178) |
| **Sejroe** | **Recreo** | *Philander opossum* | Gray four-eyed opossum | Didelphimorphia | Mammalia | Nicaragua | (65) |
| **Sejroe** | **Ricardi** | *Bos taurus* | Cattle | Artiodactyla | Mammalia | Peru | (138) |
| **Sejroe** | **Ricardi** | *Equus caballus* | Horse | Perissodactyla | Mammalia | Austria, Germany, Italy, Luxembourg, Netherlands, Portugal, Switzerland, United Kingdom of Great Britain and Northern Ireland | (36) |
| **Sejroe** | **Saxkoebing** | *Apodemus flavicollis* | Yellow-necked field mouse | Rodentia | Mammalia | Croatia, Croatia, Denmark, United Kingdom of Great Britain and Northern Ireland | (34, 51, 361, 383) |
| **Sejroe** | **Saxkoebing** | *Apodemus sylvaticus* | Common field mouse | Rodentia | Mammalia | Croatia, Romania, United Kingdom of Great Britain and Northern Ireland | (34, 220, 361) |
| **Sejroe** | **Saxkoebing** | *Canis lupus* | Dog | Carnivora | Mammalia | United Kingdom of Great Britain and Northern Ireland | (361) |
| **Sejroe** | **Saxkoebing** | *Clethrionomys glareolus* | Bank vole | Rodentia | Mammalia | United Kingdom of Great Britain and Northern Ireland | (361) |
| **Sejroe** | **Saxkoebing** | *Erinaceus europaeus* | Hedgehog | Eulipotyphla | Mammalia | Italy | (11) |

| | | | | | | | |
|---|---|---|---|---|---|---|---|
| **Sejroe** | **Saxkoebing** | *Meles meles* | Badger | Carnivora | Mammalia | United Kingdom of Great Britain and Northern Ireland | (361, 362) |
| **Sejroe** | **Saxkoebing** | *Microtus agrestis* | Field vole | Rodentia | Mammalia | Germany, United Kingdom of Great Britain and Northern Ireland | (361, 362, 384) |
| **Sejroe** | **Saxkoebing** | *Microtus arvalis* | Common vole | Rodentia | Mammalia | Russian Federation | (385) |
| **Sejroe** | **Saxkoebing** | *Microtus spp.* | Vole | Rodentia | Mammalia | United Kingdom of Great Britain and Northern Ireland | (362) |
| **Sejroe** | **Saxkoebing** | *Mus musculus* | House mouse | Rodentia | Mammalia | Germany, Romania | (224, 243) |
| **Sejroe** | **Saxkoebing** | *Mus spicilegus* | Steppe mouse | Rodentia | Mammalia | Romania | (220, 224) |
| **Sejroe** | **Saxkoebing** | *Vulpes vulpes* | Red fox | Carnivora | Mammalia | United Kingdom of Great Britain and Northern Ireland | (361) |
| **Sejroe** | **Saxkoebing** | Not stated | Rodent | Rodentia | Mammalia | Poland | (29) |
| **Sejroe** | **Saxkoebing** | Not stated | Vole | Rodentia | Mammalia | United Kingdom of Great Britain and Northern Ireland | (361) |
| **Sejroe** | **Sejroe** | *Bos taurus* | Cattle | Artiodactyla | Mammalia | United Kingdom of Great Britain and Northern Ireland | (386) |
| **Sejroe** | **Sejroe** | *Canis lupus* | Dog | Carnivora | Mammalia | Italy | (387) |
| **Sejroe** | **Sejroe** | *Capra aegagrus* | Goat | Artiodactyla | Mammalia | Spain | (207) |
| **Sejroe** | **Sejroe** | *Herpestes auropunctatus* | Lesser Indian mongoose | Carnivora | Mammalia | Guadeloupe, United States of America | (13, 24, 106, 388) |
| **Sejroe** | **Sejroe** | *Lacerta agilis* | Sand lizard | Squamata | Reptilia | Slovakia | (389) |
| **Sejroe** | **Sejroe** | *Microtus arvalis* | Common vole | Rodentia | Mammalia | Poland | (199) |
| **Sejroe** | **Sejroe** | *Microtus fortis* | Reed vole | Rodentia | Mammalia | China | (1) |
| **Sejroe** | **Sejroe** | *Mus musculus* | House mouse | Rodentia | Mammalia | Romania | (224) |
| **Sejroe** | **Sejroe** | *Mus spicilegus* | Steppe mouse | Rodentia | Mammalia | Romania | (220, 224) |
| **Sejroe** | **Sejroe** | *Myocastor coypus* | Nutria | Rodentia | Mammalia | France | (254) |
| **Sejroe** | **Sejroe** | *Ovis aries* | Sheep | Artiodactyla | Mammalia | Spain | (207) |

| | | | | | | | |
|---|---|---|---|---|---|---|---|
| **Sejroe** | **Sejroe** | Not stated | Rodent | Rodentia | Mammalia | Poland | (29) |
| **Sejroe** | **Unipertama** | *Bos taurus* | Cattle | Artiodactyla | Mammalia | Malaysia | (4, 390) |
| **Sejroe** | **Wolffi** | *Apodemus sylvaticus* | Common field mouse | Rodentia | Mammalia | Romania | (220) |
| **Sejroe** | **Wolffi** | *Bos taurus* | Cattle | Artiodactyla | Mammalia | Brazil | (313) |
| **Sejroe** | **Wolffi** | *Mus musculus* | House mouse | Rodentia | Mammalia | Romania | (222) |
| **Sejroe** | **Wolffi** | *Mus spicilegus* | Steppe mouse | Rodentia | Mammalia | Romania | (220, 222) |
| **Semeranga** | **Patoc** | *Bos taurus* | Cattle | Artiodactyla | Mammalia | United States of America | (391) |
| **Semeranga** | **Patoc** | *Felis catus* | Cat | Carnivora | Mammalia | Egypt | (178) |
| **Shermani** | **Babudieri** | *Canis lupus* | Dog | Carnivora | Mammalia | Colombia | (235) |
| **Shermani** | **Babudieri** | *Sus scrofa* | Pig | Artiodactyla | Mammalia | Peru | (219) |
| **Shermani** | **Carimagua** | *Caluromys philander* | Bare-tailed woolly opossum | Didelphimorphia | Mammalia | Colombia | (392) |
| **Shermani** | **Luis** | *Philander opossum* | Gray four-eyed opossum | Didelphimorphia | Mammalia | Peru | (145) |
| **Shermani** | **Shermani** | *Proechimys semispinosus* | Spiny rat | Rodentia | Mammalia | Panama | (65) |
| **Tarassovi** | **Atchafalaya** | *Didelphis marsupialis* | Common opossum | Didelphimorphia | Mammalia | United States of America | (104) |
| **Tarassovi** | **Atchafalaya** | *Herpestes auropunctatus* | Lesser Indian mongoose | Carnivora | Mammalia | Grenada | (66) |
| **Tarassovi** | **Bakeri** | *Didelphis marsupialis* | Common opossum | Didelphimorphia | Mammalia | United States of America | (104) |
| **Tarassovi** | **Bakeri** | Not stated | Opossum | Didelphimorphia | Mammalia | United States of America | (19) |
| **Tarassovi** | **Bravo** | *Liomys adspersus* | Spiny pocket mouse | Rodentia | Mammalia | Panama | (137) |
| **Tarassovi** | **Bravo** | *Proechimys semispinosus* | Spiny rat | Rodentia | Mammalia | Panama | (137) |
| **Tarassovi** | **Chagres** | *Proechimys semispinosus* | Spiny rat | Rodentia | Mammalia | Panama | (65) |

| | | | | | | | |
|---|---|---|---|---|---|---|---|
| **Tarassovi** | **Darien** | *Philander opossum* | Gray four-eyed opossum | Didelphimorphia | Mammalia | Panama | (65) |
| **Tarassovi** | **Gatuni** | *Proechimys semispinosus* | Spiny rat | Rodentia | Mammalia | Panama | (65) |
| **Tarassovi** | **Guidae** | *Echymipera kalubu* | Common echymipera | Peramelemorphia | Mammalia | Papua New Guinea | (393) |
| **Tarassovi** | **Guidae** | *Sus scrofa* | Pig | Artiodactyla | Mammalia | Romania, Russian Federation | (394, 395) |
| **Tarassovi** | **Kanana** | *Tatera robusta* | Fringe-tailed gerbil | Rodentia | Mammalia | Kenya | (83) |
| **Tarassovi** | **Navet** | *Bufo marinus* | Giant marine toad | Anura | Amphibia | Grenada | (66) |
| **Tarassovi** | **Ngavi** | *Bos taurus* | Cattle | Artiodactyla | Mammalia | Zimbabwe | (50, 396) |
| **Tarassovi** | **Osetica** | *Sus scrofa* | Pig | Artiodactyla | Mammalia | Russian Federation | (394) |
| **Tarassovi** | **Rama** | *Philander opossum* | Gray four-eyed opossum | Didelphimorphia | Mammalia | Nicaragua | (174) |
| **Tarassovi** | **Tarassovi** | *Apodemus flavicollis* | Yellow-necked field mouse | Rodentia | Mammalia | Romania | (220) |
| **Tarassovi** | **Tarassovi** | *Apodemus sylvaticus* | Common field mouse | Rodentia | Mammalia | Romania | (395) |
| **Tarassovi** | **Tarassovi** | *Bos taurus* | Cattle | Artiodactyla | Mammalia | Russian Federation, United States of America | (397, 398) |
| **Tarassovi** | **Tarassovi** | *Canis lupus* | Dog | Carnivora | Mammalia | New Zealand | (329) |
| **Tarassovi** | **Tarassovi** | *Didelphis marsupialis* | Common opossum | Didelphimorphia | Mammalia | United States of America | (104) |
| **Tarassovi** | **Tarassovi** | *Hydromys chrysogaster* | Rakali (water rat) | Rodentia | Mammalia | Australia | (28) |
| **Tarassovi** | **Tarassovi** | *Mephitis mephitis* | Striped skunk | Carnivora | Mammalia | United States of America | (102-104) |
| **Tarassovi** | **Tarassovi** | *Pseudemys scripta-elegans* | Slider turtle | Testudines | Reptilia | United States of America | (399) |
| **Tarassovi** | **Tarassovi** | *Rattus bowersi* | Bower's white toothed rat | Rodentia | Mammalia | Malaysia | (27) |
| **Tarassovi** | **Tarassovi** | *Rattus fuscipes* | Bush rat | Rodentia | Mammalia | Australia | (14) |
| **Tarassovi** | **Tarassovi** | *Rattus spp.* | Rat | Rodentia | Mammalia | Philippines | (133) |

| | | | | | | | |
|---|---|---|---|---|---|---|---|
| **Tarassovi** | **Tarassovi** | *Sundamys muelleri* | Mueller's rat | Rodentia | Mammalia | Malaysia | (27) |
| **Tarassovi** | **Tarassovi** | *Sus scrofa* | Pig | Artiodactyla | Mammalia | Chile, Cuba, India, New Zealand, Romania, Russian Federation | (275, 394, 395, 400-403) |
| **Tarassovi** | **Tarassovi** | *Uromys caudimaculatus* | Giant white-tailed rat | Rodentia | Mammalia | Australia | (14, 28) |
| **Tarassovi** | **Topaz** | *Bos taurus* | Cattle | Artiodactyla | Mammalia | Australia | (404) |
| **Tarassovi** | **Topaz** | Not stated | Bandicoot | Peramelemorphia | Mammalia | Australia | (405) |
| **Tarassovi** | **Vietnam** | *Sus scrofa* | Pig | Artiodactyla | Mammalia | Viet Nam, Russian Federation | (397) |
| **Tarassovi** | **Yunxian** | *Sus scrofa* | Pig | Artiodactyla | Mammalia | China | (293) |
| **Undesignated** | **Bananal** | *Hydrochoerus hydrochaeris* | Capybara | Rodentia | Mammalia | Brazil | (172, 406) |
| **Undesignated** | **Biflexa** | *Bos taurus* | Cattle | Artiodactyla | Mammalia | Malaysia | (4) |
| **Undesignated** | **Carioca** | *Capra aegagrus* | Goat | Artiodactyla | Mammalia | Brazil | (407) |
| **Undesignated** | **Neville** | *Bos taurus* | Cattle | Artiodactyla | Mammalia | Malaysia | (408) |
| **Undesignated** | **Room22** | *Crocidura russula* | White-toothed shrew | Eulipotyphla | Mammalia | Ireland | (409) |

#### Table S11 PRISMA checklist

| Section and Topic | Item # | Checklist item | Location where item is reported |
|---|---|---|---|
| **TITLE** | | | |
| Title | 1 | Identify the report as a systematic review. | Title |
| **ABSTRACT** | | | |
| Abstract | 2 | See the PRISMA 2020 for Abstracts checklist. | Abstract, adhered to the wordcount of the journal so databases are not listed. |
| **INTRODUCTION** | | | |
| Rationale | 3 | Describe the rationale for the review in the context of existing knowledge. | Introduction, last para |
| Objectives | 4 | Provide an explicit statement of the objective(s) or question(s) the review addresses. | Introduction, last para |
| **METHODS** | | | |
| Eligibility criteria | 5 | Specify the inclusion and exclusion criteria for the review and how studies were grouped for the syntheses. | Methods, second para, data analysis |
| Information sources | 6 | Specify all databases, registers, websites, organisations, reference lists and other sources searched or consulted to identify studies. Specify the date when each source was last searched or consulted. | Methods, first para, |
| Search strategy | 7 | Present the full search strategies for all databases, registers and websites, including any filters and limits used. | S1 search strategy |
| Selection process | 8 | Specify the methods used to decide whether a study met the inclusion criteria of the review, including how many reviewers screened each record and each report retrieved, whether they worked independently, and if applicable, details of automation tools used in the process. | Methods, third para |
| Data collection process | 9 | Specify the methods used to collect data from reports, including how many reviewers collected data from each report, whether they worked independently, any processes for obtaining or confirming data from study investigators, and if applicable, details of automation tools used in the process. | Methods, data abstraction |

| Section and Topic | Item # | Checklist item | Location where item is reported |
|---|---|---|---|
| Data items | 10a | List and define all outcomes for which data were sought. Specify whether all results that were compatible with each outcome domain in each study were sought (e.g. for all measures, time points, analyses), and if not, the methods used to decide which results to collect. | Methods, data abstraction |
| | 10b | List and define all other variables for which data were sought (e.g. participant and intervention characteristics, funding sources). Describe any assumptions made about any missing or unclear information. | Methods, data abstraction |
| Study risk of bias assessment | 11 | Specify the methods used to assess risk of bias in the included studies, including details of the tool(s) used, how many reviewers assessed each study and whether they worked independently, and if applicable, details of automation tools used in the process. | Not applicable, discussion – last para |
| Effect measures | 12 | Specify for each outcome the effect measure(s) (e.g. risk ratio, mean difference) used in the synthesis or presentation of results. | Not applicable |
| Synthesis methods | 13a | Describe the processes used to decide which studies were eligible for each synthesis (e.g. tabulating the study intervention characteristics and comparing against the planned groups for each synthesis (item #5)). | Not applicable |
| | 13b | Describe any methods required to prepare the data for presentation or synthesis, such as handling of missing summary statistics, or data conversions. | Data analysis |
| | 13c | Describe any methods used to tabulate or visually display results of individual studies and syntheses. | Data analysis |
| | 13d | Describe any methods used to synthesize results and provide a rationale for the choice(s). If meta-analysis was performed, describe the model(s), method(s) to identify the presence and extent of statistical heterogeneity, and software package(s) used. | Data analysis |
| | 13e | Describe any methods used to explore possible causes of heterogeneity among study results (e.g. subgroup analysis, meta-regression). | Not applicable |
| | 13f | Describe any sensitivity analyses conducted to assess robustness of the synthesized results. | Not applicable |
| Reporting bias assessment | 14 | Describe any methods used to assess risk of bias due to missing results in a synthesis (arising from reporting biases). | Discussion |
| Certainty assessment | 15 | Describe any methods used to assess certainty (or confidence) in the body of evidence for an outcome. | Not applicable |
| **RESULTS** | | | |
| Study selection | 16a | Describe the results of the search and selection process, from the number of records identified in the search to the number of studies included in the review, ideally using a flow diagram. | Results, first para |
| | 16b | Cite studies that might appear to meet the inclusion criteria, but which were excluded, and explain why they were excluded. | Text S3 |
| Study | 17 | Cite each included study and present its characteristics. | Table S10 |

| Section and Topic | Item # | Checklist item | Location where item is reported |
|---|---|---|---|
| characteristics | | | |
| Risk of bias in studies | 18 | Present assessments of risk of bias for each included study. | Not applicable |
| Results of individual studies | 19 | For all outcomes, present, for each study: (a) summary statistics for each group (where appropriate) and (b) an effect estimate and its precision (e.g. confidence/credible interval), ideally using structured tables or plots. | Results, Table 1, Figure 2, Figure 3, Figure 4 |
| Results of syntheses | 20a | For each synthesis, briefly summarise the characteristics and risk of bias among contributing studies. | Not applicable |
| | 20b | Present results of all statistical syntheses conducted. If meta-analysis was done, present for each the summary estimate and its precision (e.g. confidence/credible interval) and measures of statistical heterogeneity. If comparing groups, describe the direction of the effect. | Not applicable |
| | 20c | Present results of all investigations of possible causes of heterogeneity among study results. | Not applicable |
| | 20d | Present results of all sensitivity analyses conducted to assess the robustness of the synthesized results. | Not applicable |
| Reporting biases | 21 | Present assessments of risk of bias due to missing results (arising from reporting biases) for each synthesis assessed. | Not applicable |
| Certainty of evidence | 22 | Present assessments of certainty (or confidence) in the body of evidence for each outcome assessed. | Not applicable |
| **DISCUSSION** | | | |
| Discussion | 23a | Provide a general interpretation of the results in the context of other evidence. | Discussion, first and second para |
| | 23b | Discuss any limitations of the evidence included in the review. | Discussion, final paragraph |
| | 23c | Discuss any limitations of the review processes used. | Discussion, final paragraph |
| | 23d | Discuss implications of the results for practice, policy, and future research. | Discussion para 5, para 6 |
| **OTHER INFORMATION** | | | |

| Section and Topic | Item # | Checklist item | Location where item is reported |
|---|---|---|---|
| Registration and protocol | 24a | Provide registration information for the review, including register name and registration number, or state that the review was not registered. | Not applicable |
| | 24b | Indicate where the review protocol can be accessed, or state that a protocol was not prepared. | Not applicable |
| | 24c | Describe and explain any amendments to information provided at registration or in the protocol. | Not applicable |
| Support | 25 | Describe sources of financial or non-financial support for the review, and the role of the funders or sponsors in the review. | Funding statement |
| Competing interests | 26 | Declare any competing interests of review authors. | Conflict of interest statment |
| Availability of data, code and other materials | 27 | Report which of the following are publicly available and where they can be found: template data collection forms; data extracted from included studies; data used for all analyses; analytic code; any other materials used in the review. | Online database, data availability statement |
